# The relationship of COVID-19 related stress and media consumption with schizotypy, depression and anxiety

**DOI:** 10.1101/2021.11.26.21266896

**Authors:** Sarah Daimer, Lorenz Mihatsch, Sharon A.S. Neufeld, Graham K. Murray, Franziska Knolle

**Affiliations:** Department of Diagnostic and Interventional Neuroradiology, School of Medicine, Technical University of Munich, Munich, Germany; Department of Anaesthesiology and Intensive Care Medicine, Ludwig-Maximilians-Universität München, Munich, Germany; Institute for Medical Information Processing, Biometry and Epidemiology, Ludwig-Maximilians-Universität München, Munich, Germany; Department of Psychiatry, University of Cambridge, Cambridge UK; Cambridgeshire and Peterborough NHS Foundation Trust, Cambridge, United Kingdom

**Keywords:** COVID-19 pandemic, Schizotypy, mental Health, Depression, Anxiety, Structural equation model

## Abstract

Studies report a strong impact of the COVID-19 pandemic and related stressors on the mental wellbeing of general population. In this paper, we investigated whether COVID-19 related concerns and social adversity affected schizotypal traits, anxiety and depression using structural equational modelling. In mediation analyses, we furthermore explored whether these associations were mediated by healthy (sleep and physical exercise) or unhealthy behaviours (drug and alcohol consumption, excessive media use).

We assessed schizotypy, depression and anxiety as well as, healthy and unhealthy behaviours and a wide range of sociodemographic scores using online surveys from residents of Germany and the United Kingdom over one year during the COVID-19 pandemic. Four independent samples were collected (April/ May 2020: N=781, September/ October 2020: N=498, January/ February 2021: N=544, May 2021: N= 486). The results revealed that COVID-19 related life concerns were significantly associated with schizotypy in the autumn 2020 and spring 2021 surveys, and with anxiety and depressive symptoms in all surveys; and social adversity significantly affected the expression of schizotypal traits in all but the spring 2020 survey, and depressive and anxiety symptoms in all samples. Importantly, we found that excessive media consumption (>4h per day) fully mediated the relationship of COVID-19 related life concerns and schizotypal traits in the winter 2021 survey. Furthermore, several of the surveys showed that excessive media consumption was associated with increased depressive and anxiety-related symptoms in people burdened by COVID-19 related life.

The ongoing uncertainties of the pandemic and the restrictions on social life have a strong impact on mental well-being and especially the expression of schizotypal traits. The negative impact is further boosted by excessive media consumption, which is especially critical for people with high schizotypal traits.

## 1. Introduction

Starting in Wuhan, China, the novel highly infectious severe acute respiratory syndrome coronavirus 2 (SARS-CoV-2, COVID-19) spread rapidly around the world in the first months of 2020 and was declared as a pandemic by the WHO om 13^th^ March 2020. At the time of writing, more than 258 million people worldwide contracted the illness and more than 5.2 million died with or from COVID-19 (Daly & Robinson, 2021; JHU, 2021). Even after successful vaccination programmes were implemented in many countries (ECDE, 2021), the uncertainties and restrictions still impacted many areas of daily life. In many countries measures such as “lockdown” or “social distancing” introduced by governments early on (Cohen & Kupferschmidt, 2020) are now procedures regularly and flexibly applied to combat the pandemic. Many educational institutions, childcare facilities, bars and restaurants have been closed over long periods during the past 1.5 years; and cultural or sporting events still being cancelled or highly restricted. Governments in different countries differed in their approach to tackling the pandemic (Plümper & Neumayer, 2020). For example, at the start of the pandemic the United Kingdom (UK) imposed a nationwide lockdown a few days after Germany, which may have led to higher case and mortality rates. However, the UK started vaccinating earlier and faster than Germany, which allowed earlier relaxations of restrictions (Balmford et al., 2020; Pritchard et al., 2021). Due to delays in vaccine purchases, significantly less vaccines were available in EU countries, which contributed to fewer vaccinations compared to the UK (Sasse, 2021). Figure 1 shows the number of cases, deaths and vaccinations per day averaged over the week until June 2021.

**Fig. 1.**
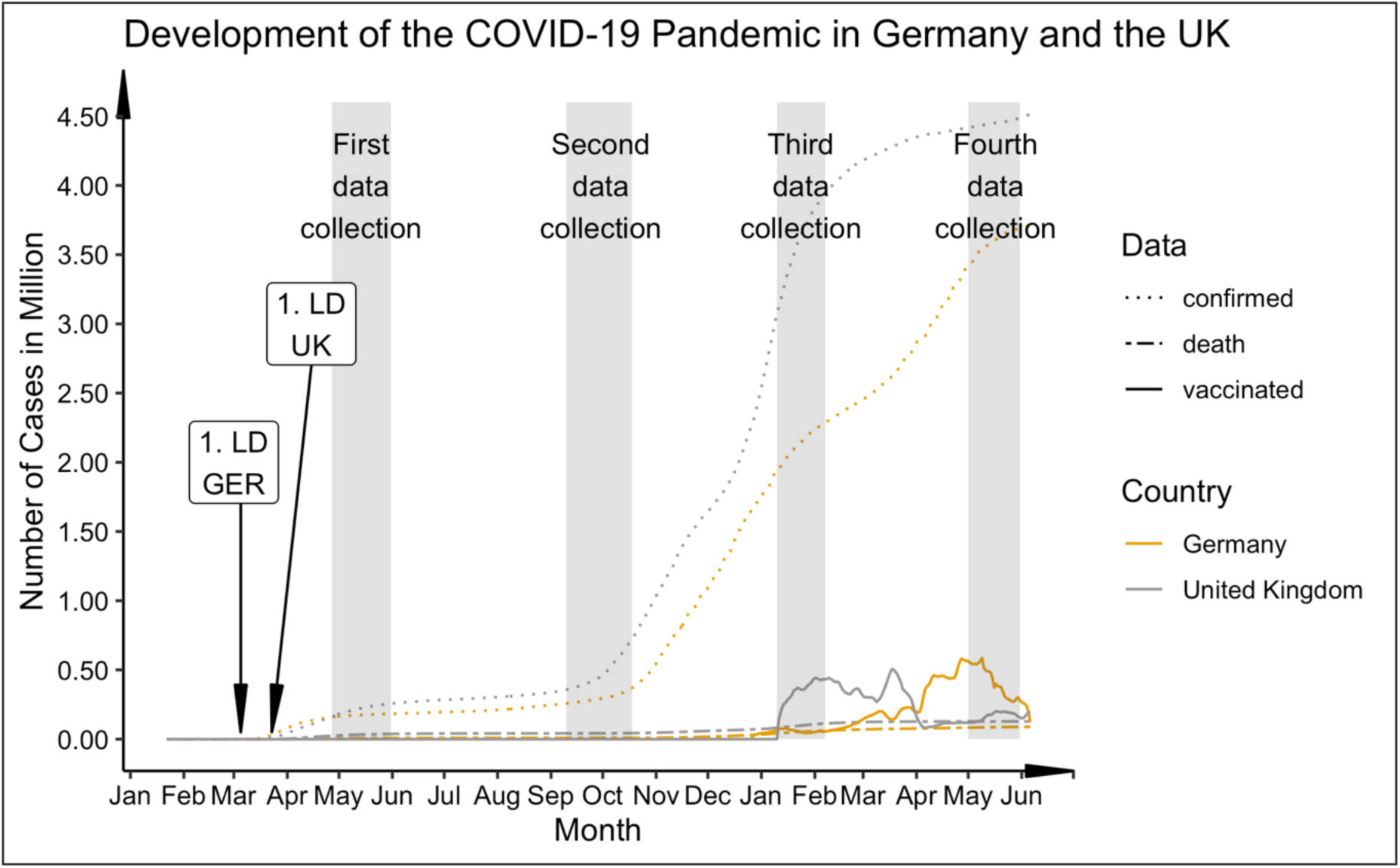
Progression of COVID-19 cases, deaths and vaccine rate (first dose) in comparison between Germany and the UK from January 2020 to June 2021. Germany: 1. Lockdown (LD): state-wise lockdown on March 16, 2020; UK: 1. Lockdown (LD): national lockdown on March 23, 2020. Numbers of vaccinations averaged over the week. Data (cases and deaths) taken from the 2019 Novel CoronaVirus CoViD (2019-nCoV) Data Repository by Johns Hopkins University Center for Systems Science and Engineering (JHU CSSE). Data for UK vaccination rate taken from GOV.UK Coronavirus in the UK (https://coronavirus.data.gov.uk/details/vaccinations) and for the German vaccination rate from Impfdashboard (https://impfdashboard.de/daten). The grey bars mark the time sections of data collection in the present study.

Early on, experts expressed concern that the fear of contracting the infection with SARS-CoV-2 as well as pandemic related stressors such as social restrictions, financial preoccupations, task overload, inadequate information, frustration and boredom posed a risk for mental health, and may be reflected in particular in depressive and anxiety-related symptoms (Amsalem et al., 2021; Brooks et al., 2020). Social distancing measures in particular represent a macro-stressor affecting the majority of people in an unprecedented manner (Ahrens et al., 2021). Different studies indeed showed that severe restrictions of social contacts as well as the fear of the virus or the impact on living conditions had a measurable impact on the mental health of general populations all over the world (Bu et al., 2020; Smith et al., 2020; C. Wang et al., 2020). During the first lockdown, increased levels of perceived stress and mental distress, COVID-19 related fear, general anxiety and depression, and a general decline in mental wellbeing were observed in many countries including, Germany and the UK (Bäuerle et al., 2020; Fancourt et al., 2020; Proto & Quintana-Domeque, 2021; Smith et al., 2020).

In this stressful context, there is the additional risk that people engage more in unhealthy behaviour and coping strategies, such as excessive consumption of alcohol or drugs, potentially due to insecurity and social restrictions (Clay & Parker, 2020; Marsden et al., 2020; McKay & Asmundson, 2020; Pfefferbaum & North, 2020). One of the first signs of this was the rise in supermarket sales of alcohol in the UK in response to pub and restaurant closures. In the week ending March 21^st^, alcohol sales rose by 67%, but total supermarket sales increased by only 43% (Finlay & Gilmore, 2020). However, reports on general alcohol consumption during the pandemic have diverse findings. Some studies reported an increase in alcohol consumption in one fifth to one third of the participants during the first weeks of the COVID-19 Pandemic in Spring 2020 (Daly & Robinson, 2021; Jacob et al., 2021; Taylor et al., 2021). A study in 21 European countries showed that alcohol consumption generally decreased in all countries, except for the UK, where alcohol consumption increased, and Ireland, where consumption remained the same (Kilian et al., 2021). However, the literature shows that the patterns of alcohol consumption change in the course of the pandemic, i.e., fewer episodes of heavy drinking (Kilian et al., 2021), but also that particular groups are at increased risk, for example individuals with children, and those with higher depressive or anxiety scores (Sallie et al., 2020). Furthermore, people who have already had an addiction problem or other psychiatric diagnosis appear particularly at risk. Marsden and colleagues showed that while the normal population did not increase their drinking behaviour, those who already showed harmful or problematic drinking patterns increased their consumption (Marsden et al., 2020). The consumption of other substances including legal, illegal and prescriptive drugs, may also have increased in response to the pandemic in general (Manthey et al., 2021), and specifically to cope with COVID-19 related stress (Czeisler et al., 2020). People with a substance use disorder may increase their consumption in reaction to the negative impact of the situation, shift to other substances if access to their primary substances become limited, or relapse, if they had already recovered from the addiction (Chiappini et al., 2020).

Besides substance use, many people may also “escape” from the current situation through excessive media consumption, or use media excessively in search for information regarding the current situation. The media plays a particular role in how individuals cope with disasters (Taylor, 2019), which also applies to the COVID-19 pandemic. On the one hand, the media provides essential information about the virus, recent developments, and protective measures; further, social media is necessary to stay in contact with others and may also be useful to distract oneself from boredom due to a lack of alternatives (Bendau et al., 2021). On the other hand, the media may produce rumours or misinformation (Tasnim et al., 2020), or perpetuate the sense of constant threat, which may lead to increased anxiety (Garfin et al., 2020; Satici et al., 2020). Interestingly, the WHO recently started using the term “infodemic” in the context of the present crisis as “a tsunami of misinformation, hate, scapegoating and scare-mongering (which) has been unleashed” (WHO, 2020). A large body of literature suggests that excessive media consumption (>3h daily) is associated with poorer mental health (e.g. (Abbas et al., 2021; Bendau et al., 2020; Gao et al., 2020; Neophytou et al., 2021; Ni et al., 2020; Su et al., 2021; Valdez et al., 2020; Y. Wang et al., 2020; Zendle, 2020)).

In the context of the COVID-19 pandemic, most general-population studies focus on the increase of symptoms from common mental disorders, i.e.: depression and anxiety. The present study extends this work by also addressing the development of schizotypal traits. Our previous studies indicated an increase in schizotypic traits during the pandemic (Daimer et al., 2021; Knolle et al., 2021). Schizotypy describes a multidimensional unifying construct that represents the underlying vulnerability for schizophrenia-spectrum psychopathology that is expressed across a broad range of personality, subclinical, and clinical psychosis phenomenology (Debbané & Barrantes-Vidal, 2015; Grant & Hennig, 2020; Raine, 1991). It is regarded as a latent multidimensional personality trait that occurs in varying degrees in every person along a continuum (Grant et al., 2018). There is no consensus on the exact content of the construct, but all definitions include positive (e.g. paranoia), negative (i.e. lack of trust) and disorganized traits, similar to schizophrenia (Lenzenweger, 2011).

The pandemic could pose an increased risk for people with pronounced schizotypal traits. For example, paranoia related to the risk of a contagious virus may result in individuals experiencing higher levels of psychological stress, depressive feelings, and anxiety (Preti et al., 2020). Fekih-Romdhane and colleagues showed that people scoring high on schizotypy were more likely to think they had COVID-19 symptoms than people with low schizotypy and they also experienced more pronounced fear of illness (Fekih-Romdhane et al., 2021). Furthermore, a comparison between individuals with low and high schizotypy during the pandemic showed that those with low schizotypy had a variety of strategies to adaptively cope with the fear of COVID-19 (e.g. positive reframing, planning, religion, self-distraction, and behavioural disengagement), yet those with high schizotypy had fewer such strategies (Fekih-Romdhane et al., 2021). Those with high schizotypy were more likely to use maladaptive strategies such as venting or self-blame, which were less effective in reducing COVID-19 related anxiety (Fekih-Romdhane et al., 2021). Due to greater levels of introversion and mistrust of others, individuals with high schizotypal traits have fewer and poorer social contacts compared to others, are less integrated into communities, and are more likely to be lonely (Kozloff et al., 2020; Le et al., 2019). When contact with others is regarded as an additional potential risk to one’s health, schizotypal individuals may further reduce their social contacts, which may take longer to be restored in the aftermath of the pandemic (Preti et al., 2020). Additionally, individuals with high schizotypal traits may be particularly likely to succumb to media misinformation, for example, due to their low agreeableness (Kwapil et al., 2018) and disposition towards believing conspiracist ideation (Barron et al., 2014). A recent study found that people with pronounced schizotypy were more likely to share false political information (Buchanan & Kempley, 2021). Buchanan and Kempley argued that such individuals are less likely to question the truth of information than those low in schizotypy. This suggests that people scoring high on schizotypy are more likely to become victims of misinformation in the media. Misinformation due to excessive media consumption following mental health deterioration has been frequently reported during this pandemic (e.g. (De Coninck et al., 2021; Su et al., 2021; Tasnim et al., 2020; Zhao & Zhou, 2020; Zhong et al., 2020)).

People with a high schizotypal personality are at increased risk of developing a schizophrenic disorder (Linscott & Morton, 2018). According to the diathesis-stress model, environmental factors, such as major psychological stressors or stressful life events, cause the onset of psychosis in the presence of a biological predisposition to schizophrenia (Debbané & Barrantes-Vidal, 2015). The psychological stress caused by the prospect of being infected by the COVID- 19 virus, as well as the social restraining measures to mitigate the virus’ spread, could potentially trigger the development of a full-blown psychosis in people with high schizotypy (Carter et al., 1995; Chapman et al., 1994; Grant & Hennig, 2020). Valdes-Florido and colleagues (2020) reported cases of psychosis triggered by the fear of infection, compulsory home-confinement or concerns about the economic consequence of the lockdown. Hu et al. (2020) reported a 25% increase in incidence of schizophrenia in January 2020 compared to previous years in China. The authors attribute this to the psychosocial stress and social distancing measures caused by COVID-19 (Hu et al., 2020). These medium and long-term effects of COVID- 19 as an adverse life event could disproportionately affect people at risk of psychosis over the next few years (Beards et al., 2013; E. Brown et al., 2020).

In this paper we therefore aim to investigate whether pandemic-related stressors, namely ‘COVID-19 related social adversity’ and ‘COVID-19 related life concerns’, have an effect on mental health, and whether these effects are mediated by healthy and unhealthy behaviours. We generated a separate latent model for each stressor across four samples, each collected at a different time point over a year from the start of the pandemic. We hypothesized that the separate COVID-19 stressors related to restrictions and financial concerns (“life concerns”), and inhibited social relationships (“social adversity”), would each be negatively associated with symptoms of common mental distress (i.e. depressive and anxiety-related symptoms), and schizotypal traits, at all timepoints during the pandemic. In addition, we hypothesised that this association will be mediated by healthy and unhealthy behaviours (respectively, exercise and sleep; alcohol, media, and drug consumption). Specifically, we expect that individuals who experience high levels of the above-mentioned stressors may damage their mental wellbeing through the unhealthy behaviours of substance use and excessive media consumption, and may improve their mental well-being through the healthy behaviours of adequate sleep and regular exercise.

## 2. Methods

### 2.1 Study design and procedure

The COVID-19 exposure and COVID-19 related mental health questionnaire was designed as an online survey using EvaSys (https://www.evasys.de, Electric Paper Evaluationssysteme GmbH, Luneburg, Germany), see Knolle et al. (2021) for a detailed description. The questionnaires were available in German and English. For participant recruitment we used a snowball sampling strategy to reach the general public in Germany and the UK. The “Spring 2020 survey” was collected from 27/04/2020 - 31/05/2020, approximately five weeks after the introduction of the nationwide lockdowns in the UK and Germany. Completion of this first survey took approximately 35 min. The three follow-up surveys took only about 15 minutes, as they did not include question on subjective change of psychological distress (“Autumn 2020 survey”: 10/09/2020 – 18/10/2020; “Winter 2021 survey”: 10/01/2021 – 07/02/2021; “Spring 2021 survey”: 01/05/2021 – 31/05/2021). Figure 1 shows the time point of data collection with respect to the development of cases and deaths in the UK and Germany. For the follow-up surveys, the respondents who provided their email addresses on any of the previous surveys were contacted again, and additional respondents were recruited via social media (i.e., Facebook, Twitter, WhatsApp) and recruitment platforms (www.callforparticipants.com). Participation was voluntary, and participants did not receive any compensation.

Ethical approval was obtained from the Ethical Commission Board of the Technical University Munich (250/20 S). All participants provided informed consent.

### 2.3. Survey measures

As described in Knolle et al. (2021) in detail, the self-reported survey assessed the following domains: demographic information (age, gender (not biological sex), education and parental education, living conditions), the Coronavirus Health Impact Survey (CRISIS, http://www.crisissurvey.org/), which assessed COVID-19 exposure (infection status, symptoms, contact), subjective mental and physical health, and healthy and unhealthy behaviour (i.e., weekly amount of sleep and exercise, consumption of alcohol, drugs, and media), and general mental health status using the Symptom Check List (SCL) (Hardt et al., 2011; Hardt & Gerbershagen, 2001; Kuhl et al., 2010). The SCL’s 27 items assess symptoms of anxiety, depression, mistrust, and somatisation. Finally, we assessed schizotypy using the of the Schizotypy Personality Questionnaire (SPQ, dichotomous version (Raine, 1991)).

### 2.4. Variables and statistical analysis

Statistical analysis and visualisations were computed using R and R Studio (R Core Team, 2016; Rstudio, 2020). We used Wilcoxon rank sum tests or Chi-square test of independence to explore differences between the countries (UK, Germany) and four surveys (April/ May 2020, September/ October 2020, January/ February 2021, May 2021) on demographics and the COVID-19 exposure variables.

To further explore the differences between the countries and time points in CRISIS variables we conducted robust ANOVAS (Mair & Wilcox, 2020) with country (UK, Germany) and survey (April/ May 2020, September/ October 2020, January/ February 2021, May 2021) as a between-subjects factor.

In order to assess the relationships between possible influences of health-damaging behaviour on depressive and anxiety-related symptoms as well as high schizotypy, we created structural equation models (SEM) using the Lavaan package in R (Rosseel, 2012). The visualisations of the models were carried out using Onyx (Brandmaier, 2021). See Figure 2 for an overview of the hypothesized models.

**Figure 2.**
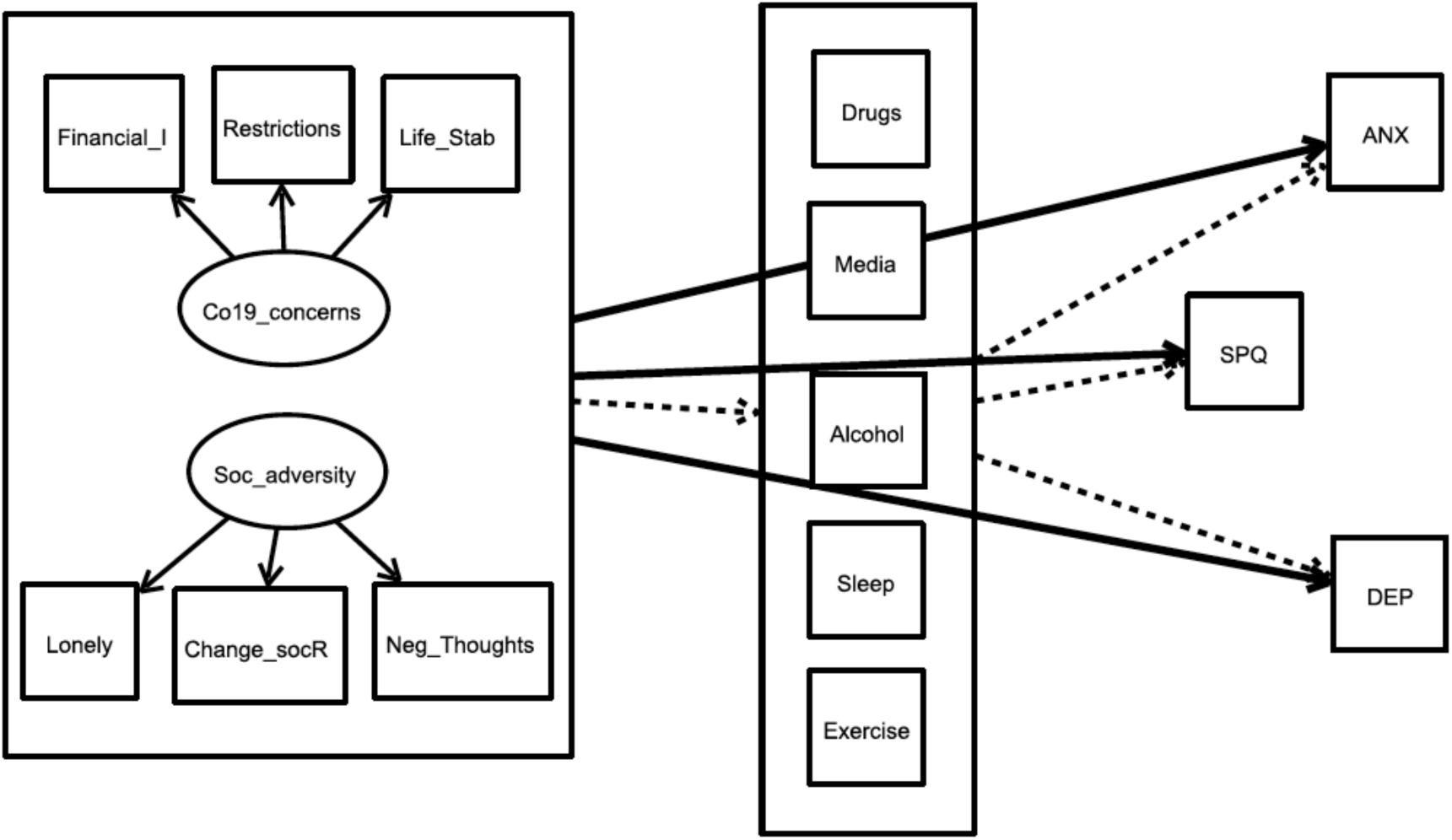
Overview of the complete theoretical model for the influence of ‘social adversity’ and ‘COVID-19 related life concerns’ directly on mental health scores and indirectly via healthy and harmful behaviour variables. The same model was calculated separately for the two exogeneous variables (“predictors”, left large box) for all four time points. The bold arrows indicate the direct pathways to the three endogenous variables (“outcomes”). The dashed arrows indicate the indirect pathways from predictor to outcomes via the five mediators (box in the middle). Co19_concerns: COVID-19 related life concerns; Soc_adversity: social adversity; Health: mental and physical health status before COVID-19; Liv_cond: living conditions; financial_I: Financial impact due to the crisis; Restrictions: Perception of the restrictions as stressful; Life_Stability: Concerns about life stability due to the crisis; Change_socR: stressful social relationship changes; ANX: anxiety symptoms with corresponding SCL-27 Items; DEP: depressive Symptoms with corresponding SCL-27 Items, SPQ: total sum scores of SPQ.

Our three endogenous manifest variables (“outcomes”) were schizotypy trait, symptoms of depression, and symptoms of anxiety. Schizotypal trait was defined as the total sum score of all SPQ items. For anxiety and depressive symptoms, specific items of the SCL-27 were summed and divided by the number of items. For depression, we used all SCL-27 items assessing dysthymic and depressive symptoms; and for anxiety we used all items describing agoraphobic and social phobic symptoms.

For the exogeneous latent variables (“predictors”), we generated two variables. The first variable, which we call ‘COVID-19 related life concerns’, reflects the direct impact of the pandemic on the individual’s life. The best fitting measurement model was formed using the CRISIS items ‘to what degree have changes related to the Coronavirus/COVID-19 crisis in your area created financial problems for you or your family’, ‘how stressful have the restrictions on your daily life been for you’ and ‘to what degree are you concerned about the stability of your living situation’. The exogeneous variables were scaled so that we fixed the variance of all factor loadings within one exogeneous latent variable to 1. This allowed comparing estimates between different models using these exogeneous latent variables quantitively. The second exogeneous latent variable, ‘social adversity’, reflects loneliness and whether change in the quality of social relationships was perceived as stressful. The variables that loaded best on the factor ‘social adversity’ were the CRISIS items ‘has the quality of the relationships between you and members of your family/social contacts changed/ how stressful have these changes in contacts been for you’, ‘how lonely were you’ and ‘to what extent did you have negative thoughts, thoughts about unpleasant experiences or things that make you feel bad’. The two measurement models for the two exogeneous latent variables (predictors) had excellent fit, see Supplementary Table 2.

The mediators drug use and excessive media use were recoded into dichotomous variables from categorical responses, due to sparse endorsement of some categories. Both were expressed with a positive value for at least one occasion of marijuana, opiate or tranquilliser use in the last four weeks, or, for at least four hours of daily consumption of any of social media, digital media, or video game (None at all, Under 1 hour, 1-3 hours, 4-6 hours, More than 6 hours), respectively. Alcohol consumption was used as a continuous variable (Not at all, Rarely, Once a month, Several times a month, Once a week, Several times a week, Once a day, More than once a day). Hours of sleep per night (<6 hours, 6-8 hours, 8-10 hours, >10 hours) during the week and frequency of 30-min exercise (sporting activities) per week were analysed with the original categories from the CRISIS measure.

For each of the four survey time points, two separate models were conducted, one to assess how ‘social adversity’ predicted mental health scores, and the second to assess how ‘COVID-19 related life concerns’ predicted mental health scores. Mental health scores were our three endogenous variables, SPQ traits, depressive symptoms and anxiety symptoms. Covariances between the outcome variables were included in the model, including covariances between mediators led to a non-converging model. As we hypothesised that these relationships are mediated by healthy and unhealthy behaviours, we included drug consumption, excessive media use, alcohol consumption, hours of sleep per night during the week, and exercise per week as mediation variables to explain the relationship between each exogeneous latent variable (‘COVID-19 related life concerns’, ‘social adversity’) and all the endogenous variables (SPQ, depression, anxiety). In order to facilitate comparison across the four surveys collected at different time points throughout the pandemic, we included as confounders in each model those variables which were associated with differences across the timepoints at p<.01 (Table 1). We did not control for suspected COVID-19 infection, as the infection rate was <1% in each of the four samples. We also included age and sex as these are known confounders of many of the paths in our proposed model. As each survey consisted of different participants, each model was based on cross-sectional data. Therefore, the direction of the associations may be ambiguous and the variables may influence each other. Nonetheless, here we examine the influence of distressing conditions, which have occurred in particular because of the COVID-19 pandemic, on mental health, and the mediation of these relationships by healthy and unhealthy behaviours. Alternative models are presented in the supplements. We fitted mediation models that included the direct effect of the exogeneous latent variable on the endogenous variable (pathway c) as well as the indirect effects via the mediation variables (pathway a+b), which included the effect of the exogeneous variable to the mediator (pathway a) and the mediator to the endogenous variable (pathway b). The total effect is calculated as the product of paths a and b plus the direct effect c. Figure 2 shows an overview of the theoretical models. Note that the models were calculated separately for the two exogeneous latent variables and the samples at the different timepoints.

**Table 1.**
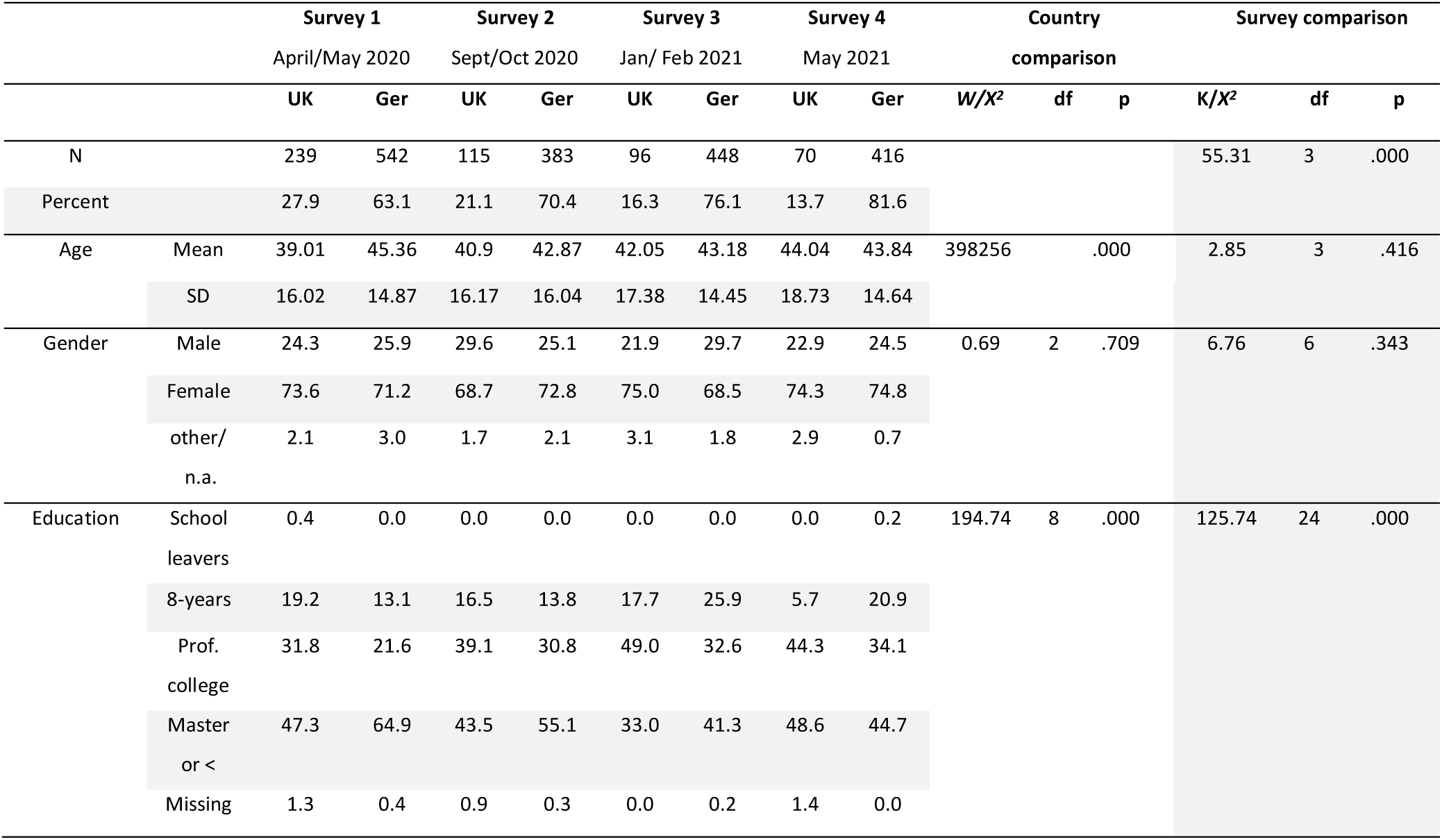
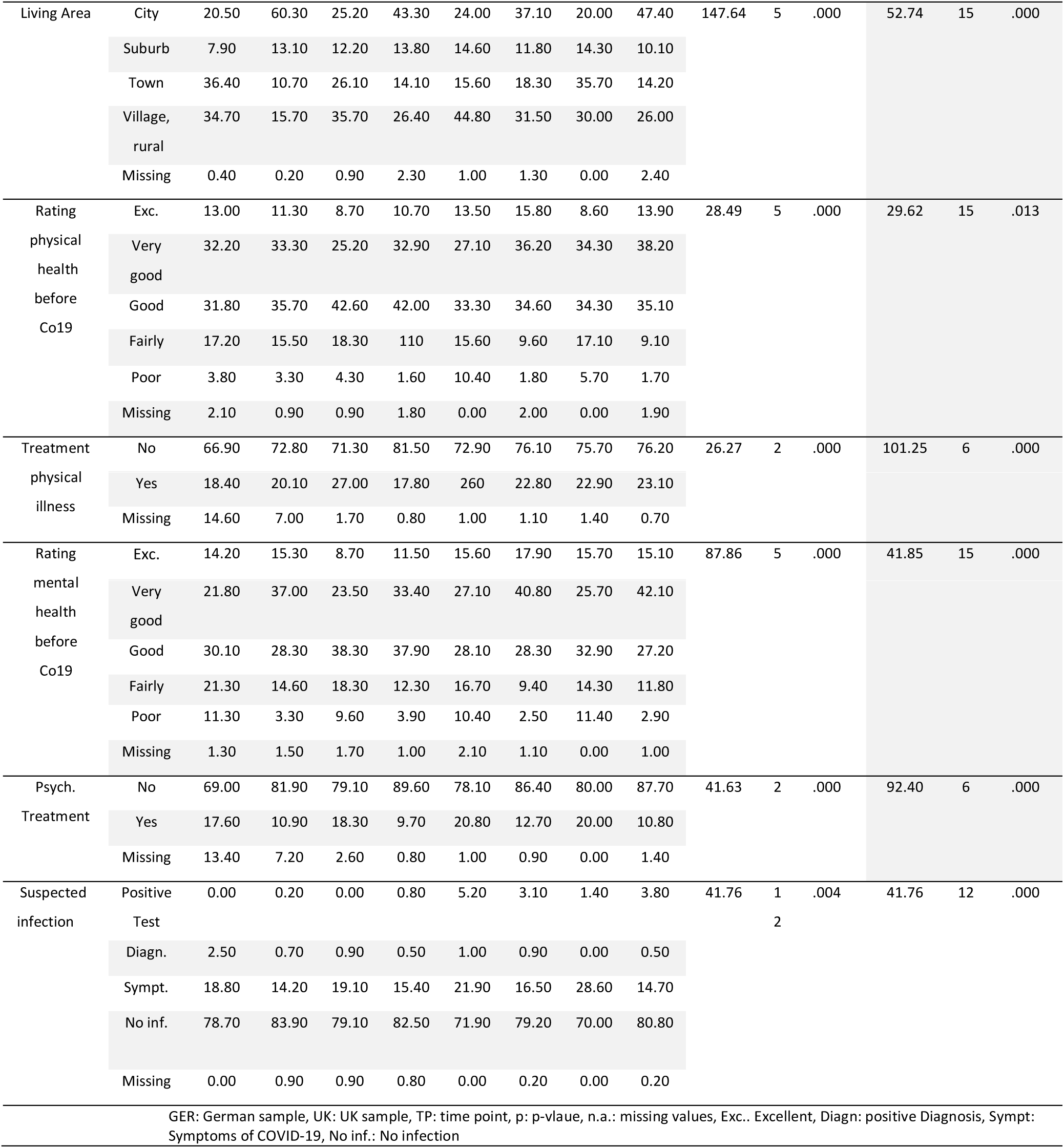
Cohort demographics and COVID-19 exposure divided by country and time point. Values in percent if not indicated otherwise.

All models were estimated using maximum likelihood estimation combined with bootstrapped errors. To examine the quality of the model fit, we consulted the Comparative-Fit-Index (CFI; (Bentler, 1990)) for the relative model fit and the root mean squared error (RMSEA; (Steiger, 1990)) for the absolute model fit. An adequate fit is assumed for RMSEA of less than .06 and a CFI of greater than .90 (Beauducel & Wittmann, 2005; Browne & Cudeck, 1992).

## 3. Results

### 3.1. Participants and sample comparisons

Sample descriptions for all four surveys are presented in Table 1. The first survey (April/May 2020) was completed by 781 participants (UK N=239; Germany N=542), after excluding three participants who did not provide consent. The second survey (September/October 2020) was completed by 498 participants (Germany, N=383; UK N=115), after excluding three responders under the age of 18, a prerequisite for taking part in the study. In the third survey (January/ February 2021), 544 participants were included after the exclusion of one person who did not give consent to the participation and four further participants under the age of 18 (Germany N=448; UK N=96). A total of 486 (Germany N=416; UK N=70) people participated in the last survey, in May 2021, after two were excluded due to lack of consent and two for being under the age of 18.

Since participation at one timepoint was not required for taking part in another one, we regard all four samples as independent. 26 participants took part in all four surveys. The samples did not differ significantly in terms of age (*X^2^*=2.85, p=.416) and gender (*X^2^* = 6.76, p =.343). However, in the second, third, and fourth survey, significantly more participants came from Germany (*X^2^*= 55.31, p <.001). In addition, the distributions of educational level (*X^2^*= 125.74, p <.001) and living area (*X^2^*= 52.74, p <.001) differed significantly between the time points (Table 1).

### 3.2 Expression of schizotypal traits, depressive and anxiety-related symptoms in the samples collected at four time points during the COVID-19 pandemic

The Kruskal-Wallis test indicated no difference in SPQ scores across the samples at the four time points, but revealed significant differences in anxiety and depressive symptoms across the samples at the four time points (Figure 3A-C; Table 2). Pearson’s correlation coefficient indicated significant high correlations between SPQ and anxiety (r=0.57- r=0.68) and between SPQ and depression (r=0.53 - r=0.58) in all four surveys (Supplementary table 1).

**Figure 3.**
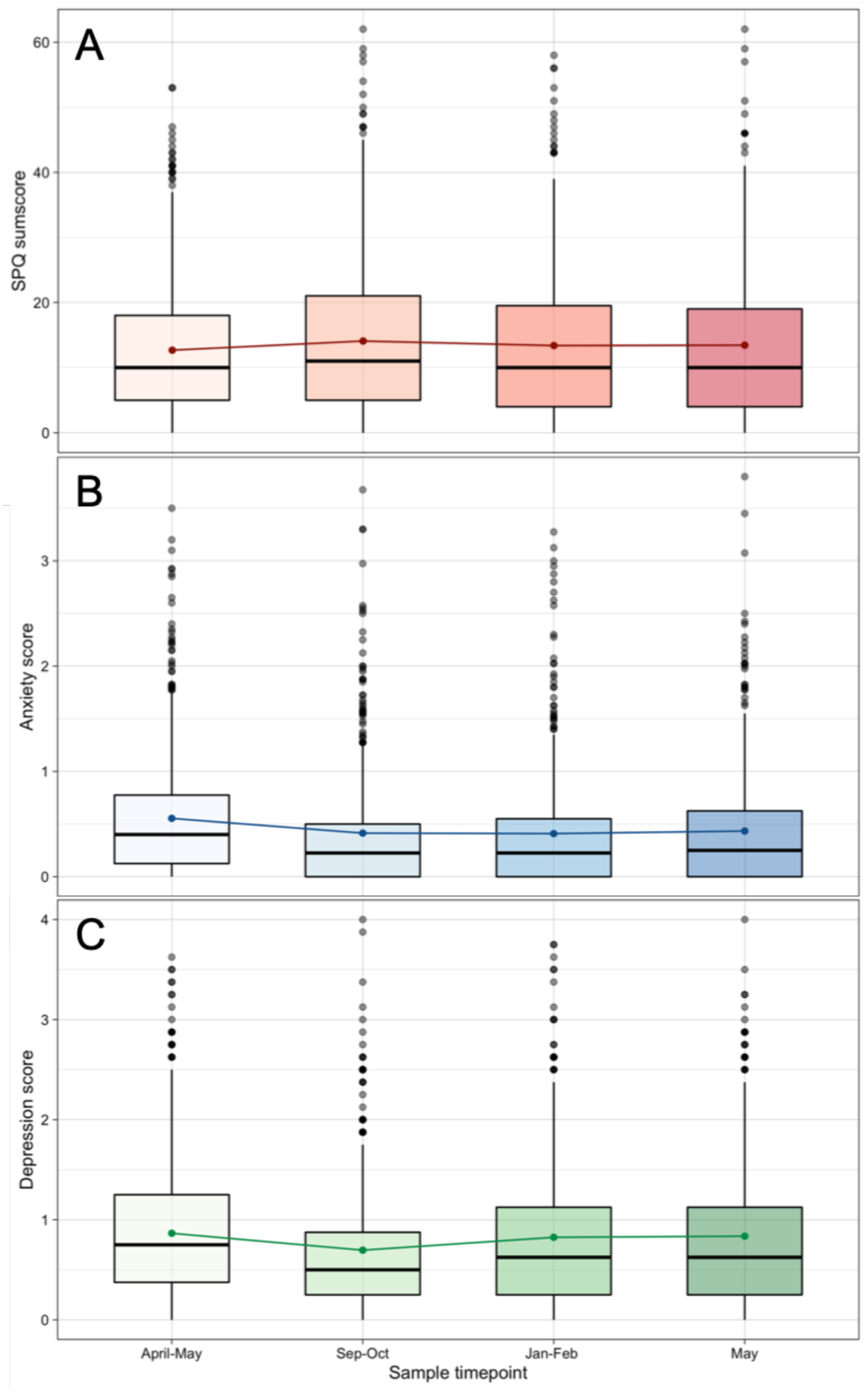
Boxplot of (A) SPQ, (B) anxiety scores and (C) depression scores separately for four time point samples. The error bars show standard deviation.

**Table 2.**
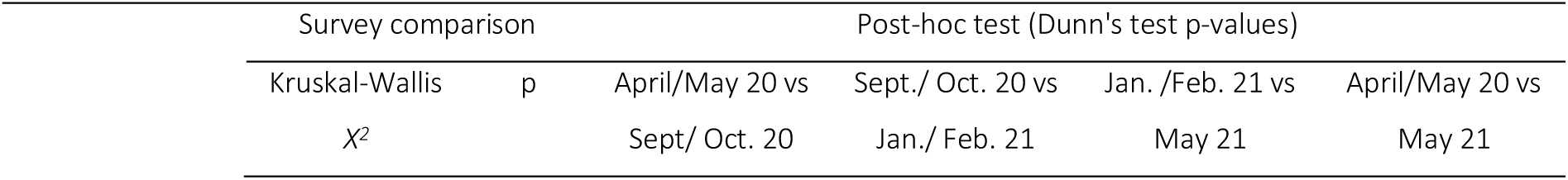
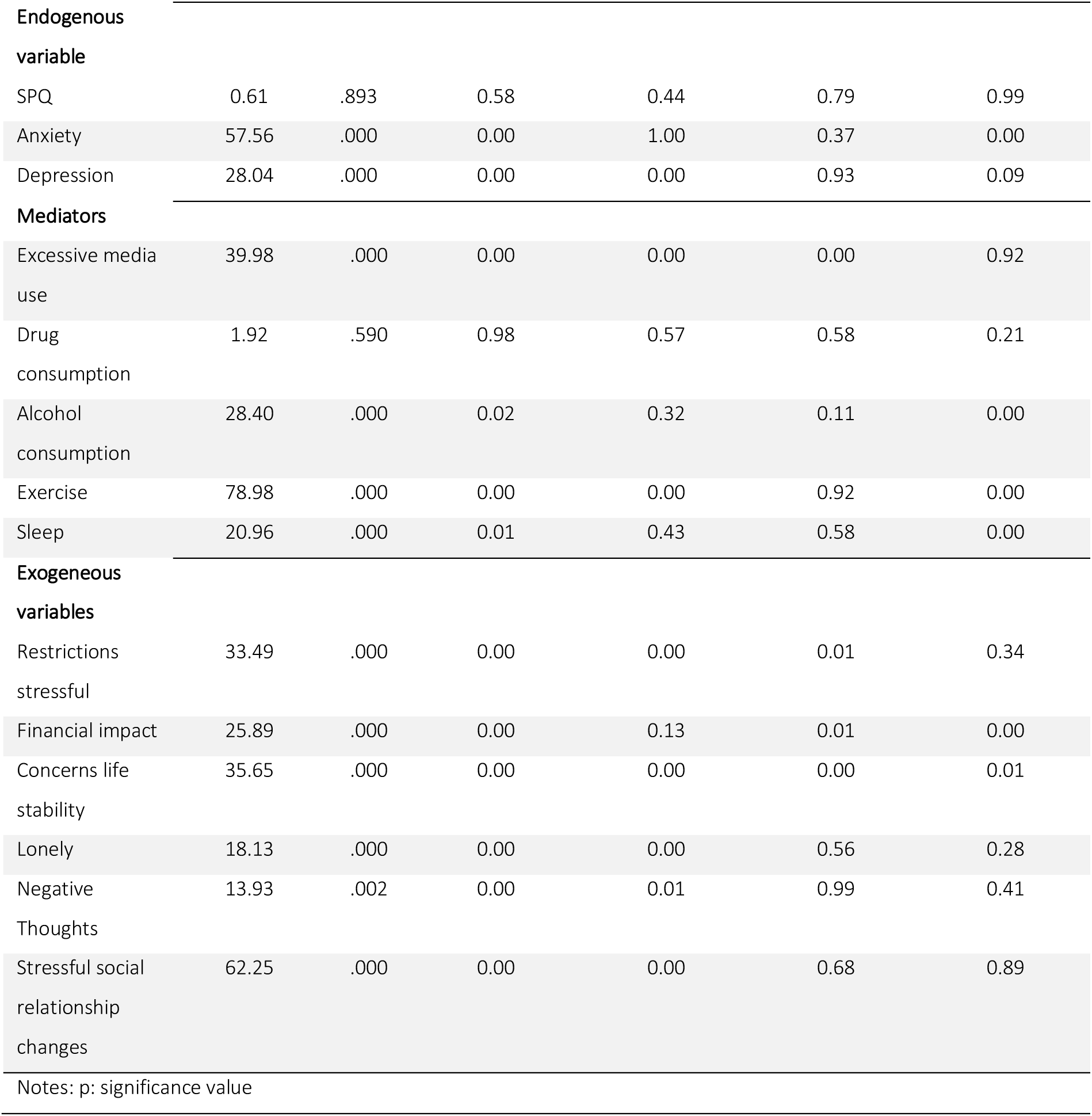
Overview of the results of the Kruskal-Wallis Chi square test and Dunn’s post-hoc test for the endogenous variables, mediators, or exogeneous latent variables in the following structural equation models. Comparisons presented are those comparing the latter survey to the prior survey, as well the last survey to the first survey.

### 3.3 Structural equation modelling

The two models showed an acceptable model fit at all four time points, with a RMSEA of ≤.08, the TLI and CFI ≥.8. Variables and items relevant for the models are described in Table 2, for different samples at different time points. An overview of all model fits is shown in Table 3. The full model statistics for the model is presented in the Supplementary materials. The two models which are presented here are highly complex, containing five mediators and three outcomes. Predictor measurement models show close to perfect fit (Supplementary Table 2). Reduced models only containing one outcome have much improved model fits, with a RMSEA of ≤.07, the TLI and CFI ≥.9; see Supplementary materials.

**Table 3.**
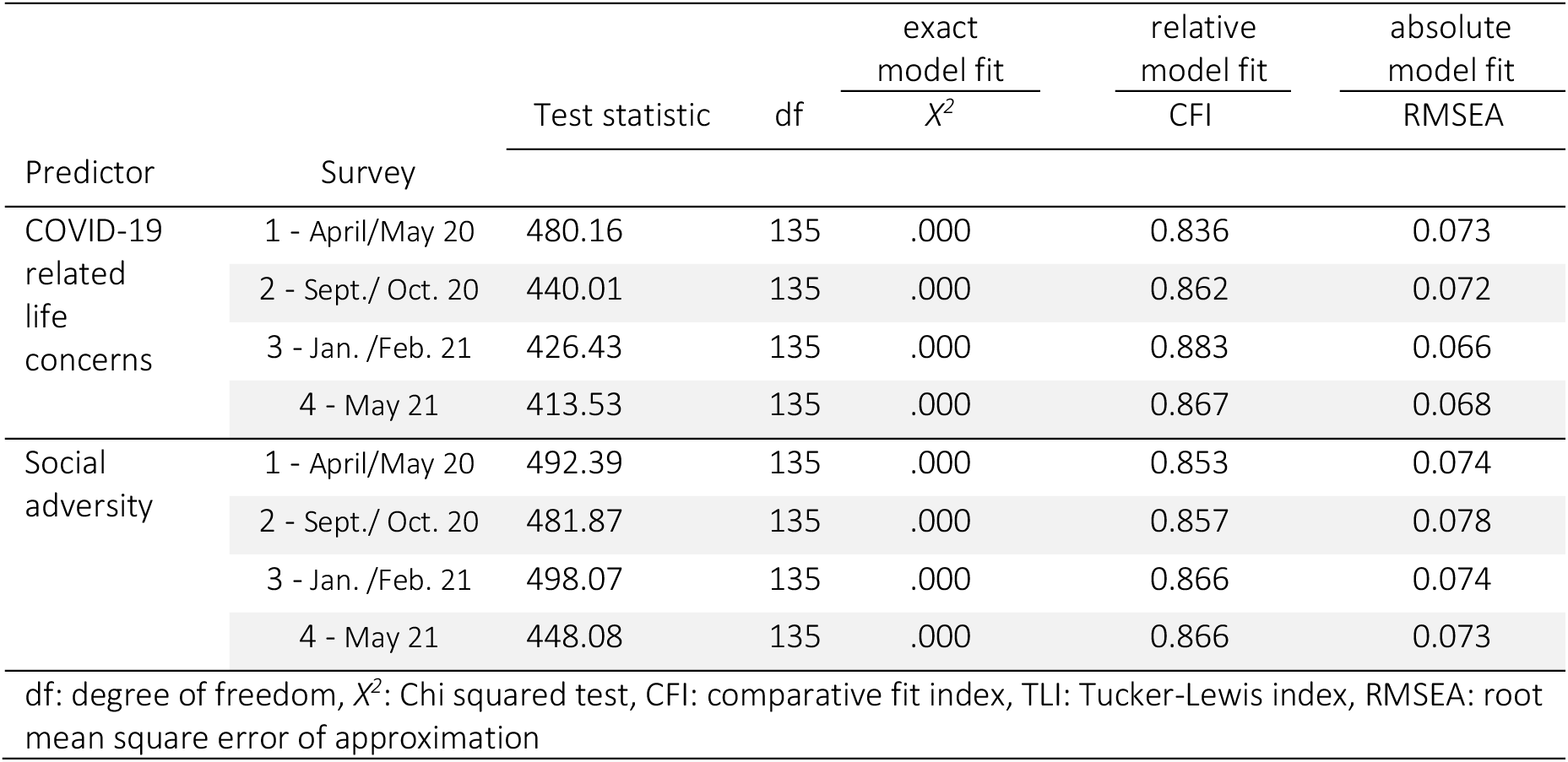
Overview of the model fit indices separated by exogeneous latent variable and survey.

| Predictor | Survey | Test statistic | df | exact<br>model fit | relative<br>model fit | absolute<br>model fit |
| --- | --- | --- | --- | --- | --- | --- |
| | | | | $\chi^2$ | CFI | RMSEA |
| COVID-19<br>related<br>life<br>concerns | 1 - April/May 20 | 480.16 | 135 | .000 | 0.836 | 0.073 |
|  | 2 - Sept./ Oct. 20 | 440.01 | 135 | .000 | 0.862 | 0.072 |
|  | 3 - Jan. /Feb. 21 | 426.43 | 135 | .000 | 0.883 | 0.066 |
|  | 4 - May 21 | 413.53 | 135 | .000 | 0.867 | 0.068 |
| Social<br>adversity | 1 - April/May 20 | 492.39 | 135 | .000 | 0.853 | 0.074 |
|  | 2 - Sept./ Oct. 20 | 481.87 | 135 | .000 | 0.857 | 0.078 |
|  | 3 - Jan. /Feb. 21 | 498.07 | 135 | .000 | 0.866 | 0.074 |
|  | 4 - May 21 | 448.08 | 135 | .000 | 0.866 | 0.073 |

df: degree of freedom, $\chi^2$ : Chi squared test, CFI: comparative fit index, TLI: Tucker-Lewis index, RMSEA: root mean square error of approximation

#### 3.2.1 ‘COVID-19 related life concerns’ model

The results for this model are presented in Table 4, and for the mediator media in Figure 4.

**Figure 4.**
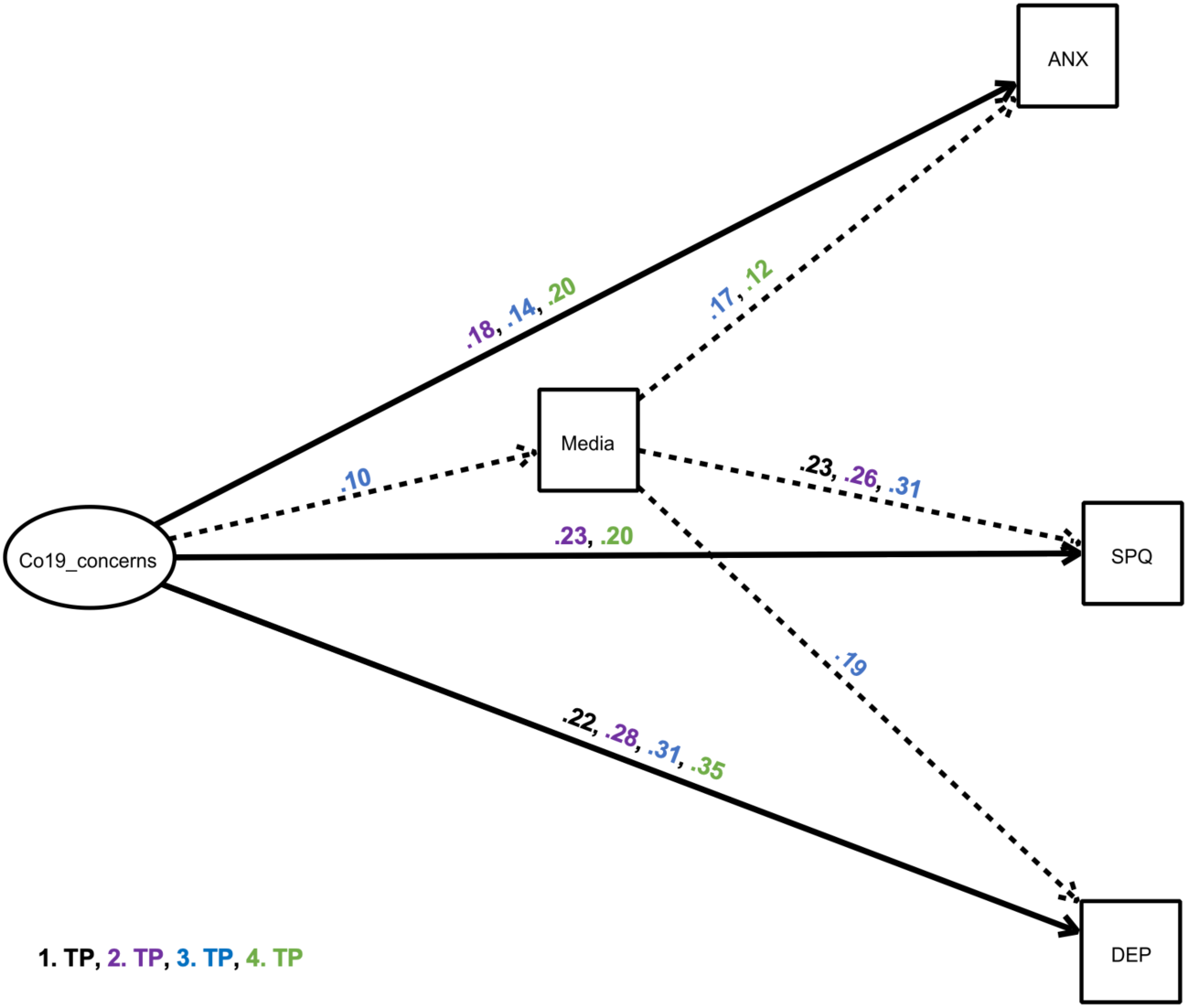
Structural equation model showing a mediator model of the impact of ‘COVID-19 related life concerns’ on depressive and anxiety-related symptoms and Schizotypal traits, via the mediator excessive media use. The exogeneous latent variable (“predictor”) had an elevating effect on the endogenous variable (“outcome”) and the mediator. Black estimates: spring 2020 survey, purple estimates: autumn 2020 survey, blue estimates: winter 2021 survey, green estimates: May 2021 survey. Solid, bold lines indicate direct effects; dashed lines indicate indirect effects. Effects are only shown where p<.05 (see Table 4). Other possible mediators were included in the model, but are not depicted for simplicity, as indirect effects were non- significant (Table 4).

**Table 4.**
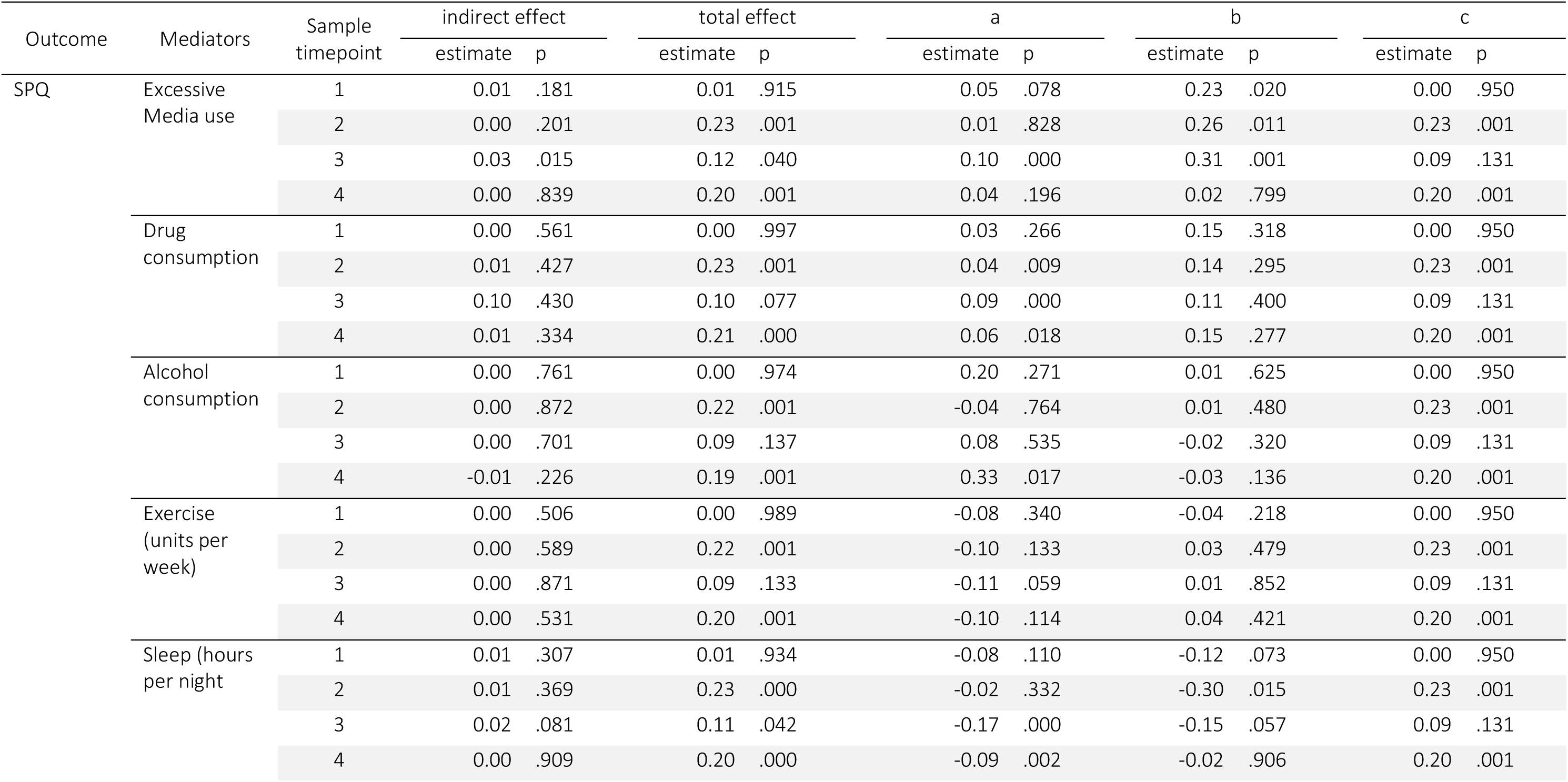
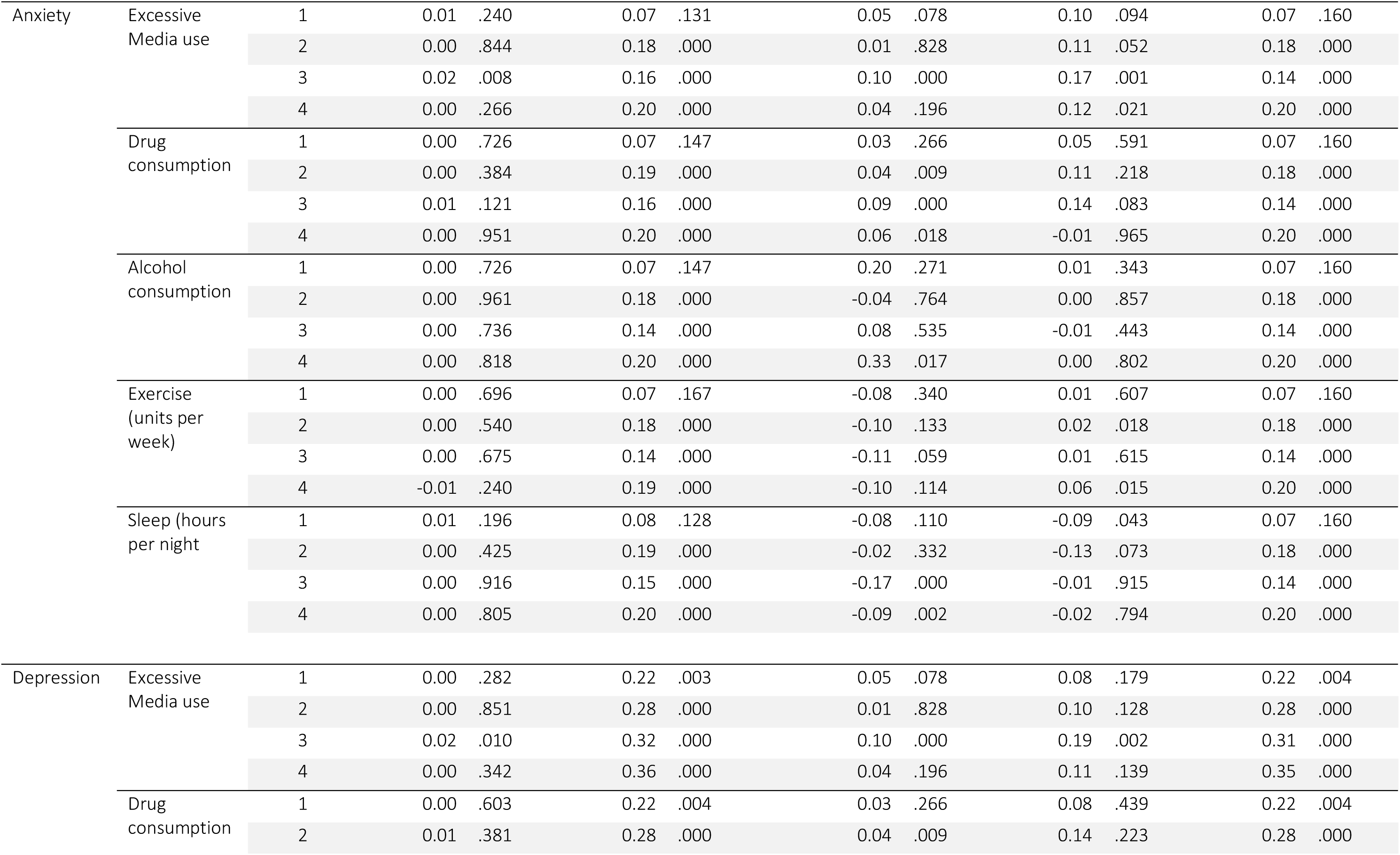
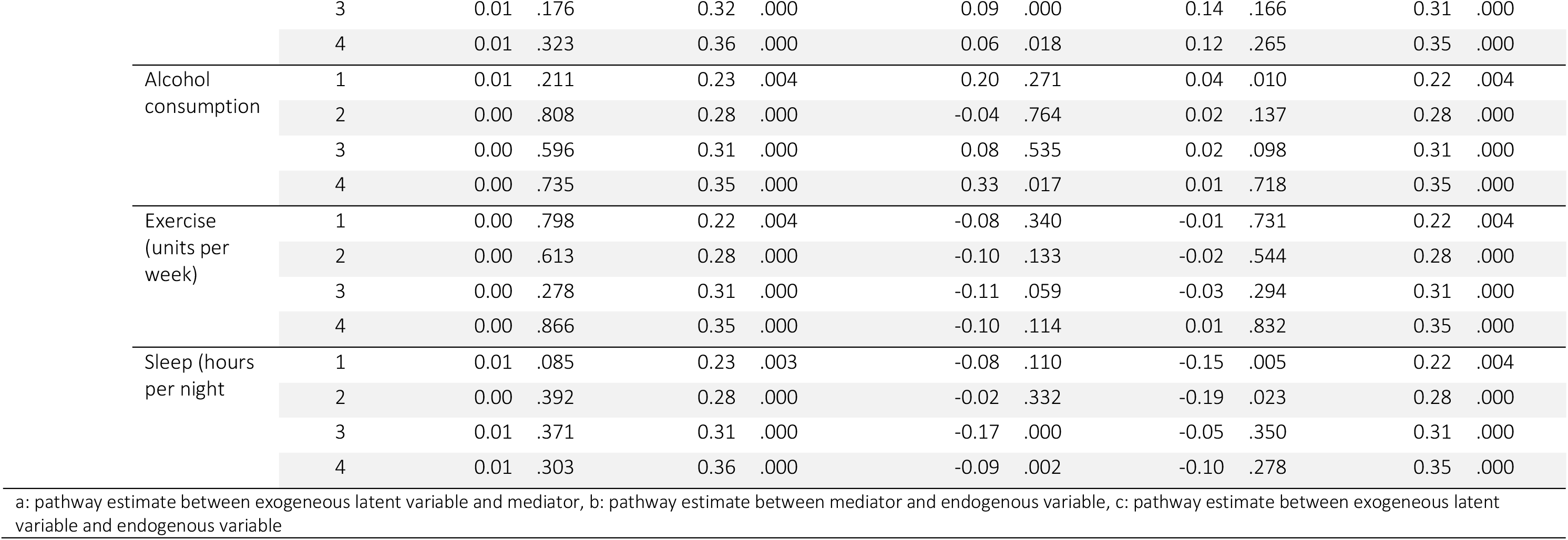
Overview of all results of the structural equation model with “COVID-19 related life concerns” as exogeneous latent variable for mental health endogenous variables mediated by harmful and healthy behaviors. The results are separated by endogenous variable (schizotypal traits, depressive symptoms, anxiety symptoms), mediators (excessive media use, drug consumption, alcohol consumption, units of exercise per week, hours of sleep per night during weeks) and samples at the four timepoints, showing indirect, direct and total effects, as well as individual pathways.

Generally, the structural equation model revealed that the exogeneous latent variable ‘COVID- 19 related life concerns’ was directly related to schizotypy (autumn 2020, spring 2021), anxiety (autumn 2020, winter 2021, spring 2021), and depression (all surveys) (pathway c), and ‘COVID- 19 related life concerns’ was positively associated with media (winter 2021), drug (autumn 2020, winter 2021, spring 2021) and alcohol consumption consumption (spring 2021), and negatively associated with sleep (winter 2021, spring 2021) (pathway a). There were significant indirect effects of ‘COVID-19 related life concerns’ on schizotypy, depression, and anxiety only through excessive media consumption (pathway a-b). The alternative models reveal a worse model fit and are presented in the supplementary materials Supplementary-table 4 for AIC/BIC in comparison.

Investigating the endogenous variable SPQ (Figure 4, Table 4), we found that concerns directly related to the COVID-19 pandemic had an increasing effect on SPQ scores in the autumn 2020 survey and May 2021 survey, with this estimate being highest at the autumn 2020 survey (c=0.23). Here, more stress due to the COVID-19 restrictions led to higher SPQ scores. At these time points as well as at the winter 2021 survey, the total effects were significant. The mediation of this effect via media consumption was significant in the winter 2021 survey. People who experienced greater COVID-19 stress were more likely to consume media excessively and have higher SPQ. Media consumption explained a quarter of the total effects of COVID-19 stress on SPQ In addition, there was a trend towards a significant mediation with sleep in the winter 2021 survey.

Investigating anxiety (Figure 4, Table 4), we found in the last three surveys that COVID-19 related concerns were associated with increased anxiety, with the strongest effect at the May 2021 survey (c=0.20) and the weakest effect in winter 2021 (c=0.14). Significant total effects were measurable at these time points. A significant indirect effect with the mediator excessive media consumption was seen in winter 2021: 12.5% of the effects of COVID-19 related concerns on anxiety symptoms occurred through excessive media consumption.

In all four surveys, increased stress from the COVID-19 pandemic was significantly related to greater depressive symptoms (Figure 4, Table 4), with this effect being lowest (c=0.22) at the spring 2020 survey and highest (c=0.35) in May 2021. Therefore, all total effects were also significant. Only in winter 2021 was there a significant indirect effect with the mediator excessive media consumption: here, excessive media consumption explained 6% of the effects of COVID-19 stress on depressive symptoms. No other statistically significant mediation effects were found.

#### 3.2.2 ‘Social adversity’ Model

The structural equation model revealed that the exogeneous latent variable ‘social adversity’ was directly related to schizotypy, anxiety, and depression in all surveys (pathway c), and social adversity was positively associated with media (winter 2021) and drug (spring 2020, winter 2021, spring 2021) consumption, and negatively associated with sleep (all surveys). Alcohol was positively correlated with social adversity at the spring 2020 survey: people who were more isolated drank more alcohol (pathway a). The alternative models revealed a worse model fit and are presented in the supplementary materials Supplementary-table 4 for AIC/BIC in comparison.

Exploring the effect of ‘social adversity’ on schizotypy (Figure 5, Table 5), we found that in all four surveys, social adversity was associated with higher levels of schizotypal traits, with this association being most pronounced in the May 2021 survey (c=0.33). Significant total effects were shown in all four surveys. The mediation with excessive media consumption revealed a statistically significant indirect effect in the winter 2021 survey: 11.5% of the effects of social adversity during the pandemic on schizotypy were mediated by excessive media consumption.

**Figure 5.**
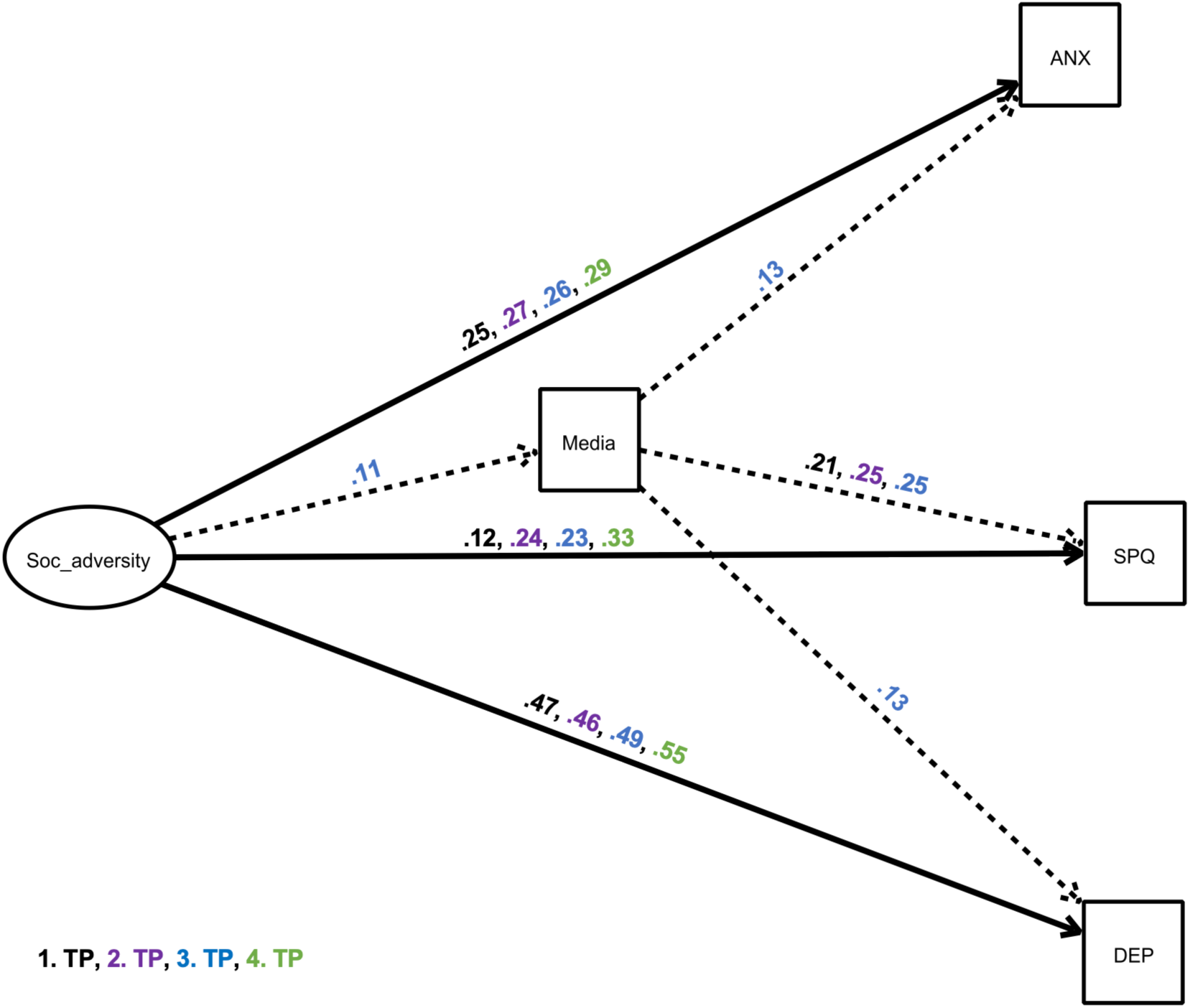
Structural equation model showing a mediator model of the impact of ‘social adversity’ on depressive and anxiety-related symptoms and schizotypal traits and via the mediator excessive media use. The exogeneous latent variable (“predictor”) had an elevating effect on the endogenous variable (“outcome”) and the mediator. We only present significant pathways (p<.05). Black estimates: spring 2020 survey, purple estimates: autumn 2020 survey, blue estimates: winter 2021, green estimates: May 2021 survey. Solid, bold lines indicate direct effects; dashed lines indicate indirect effects. Other possible mediators were included in the model, but are not depicted for simplicity, as indirect effects were non-significant (Table 5).

**Table 5.**
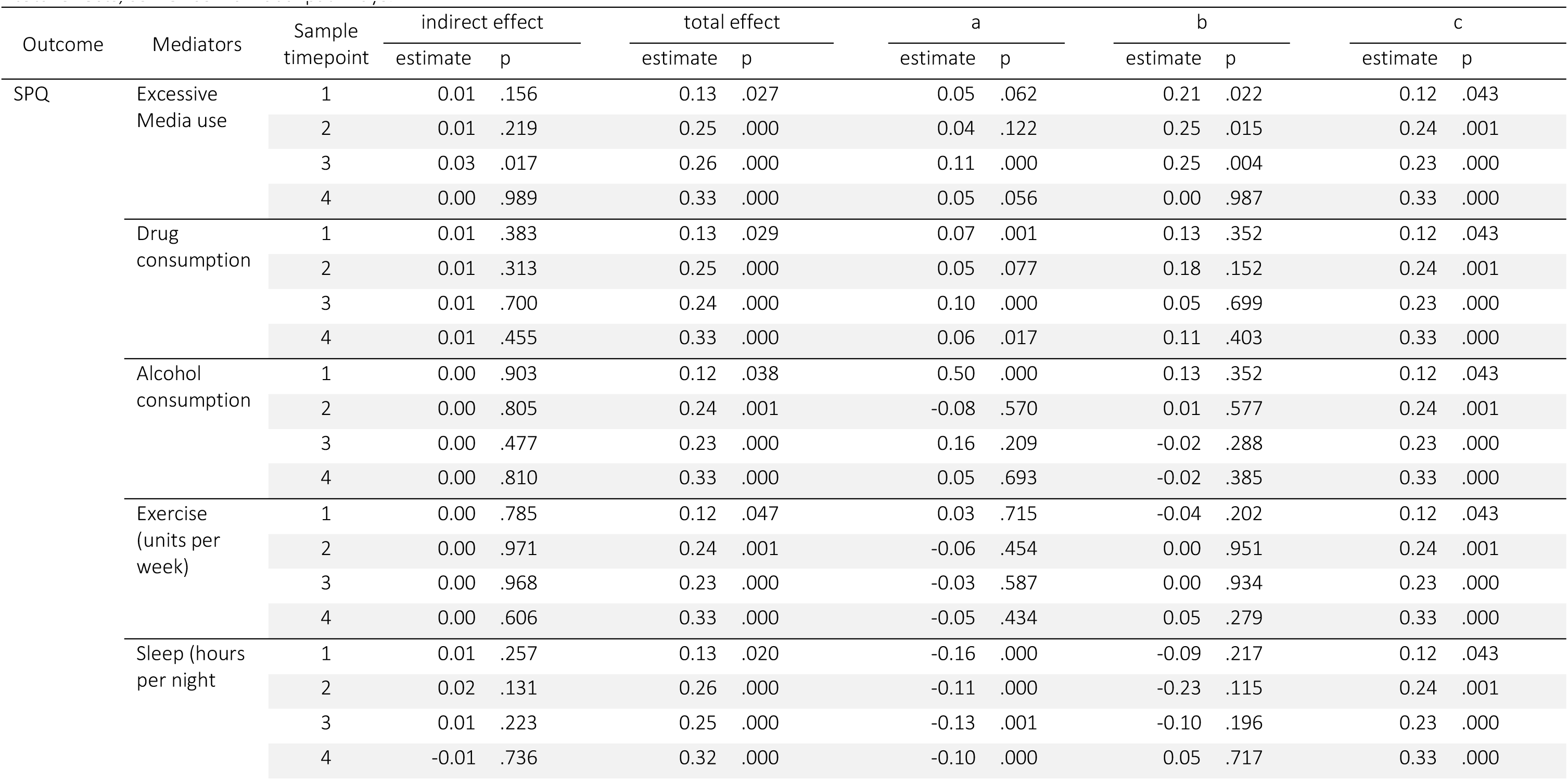
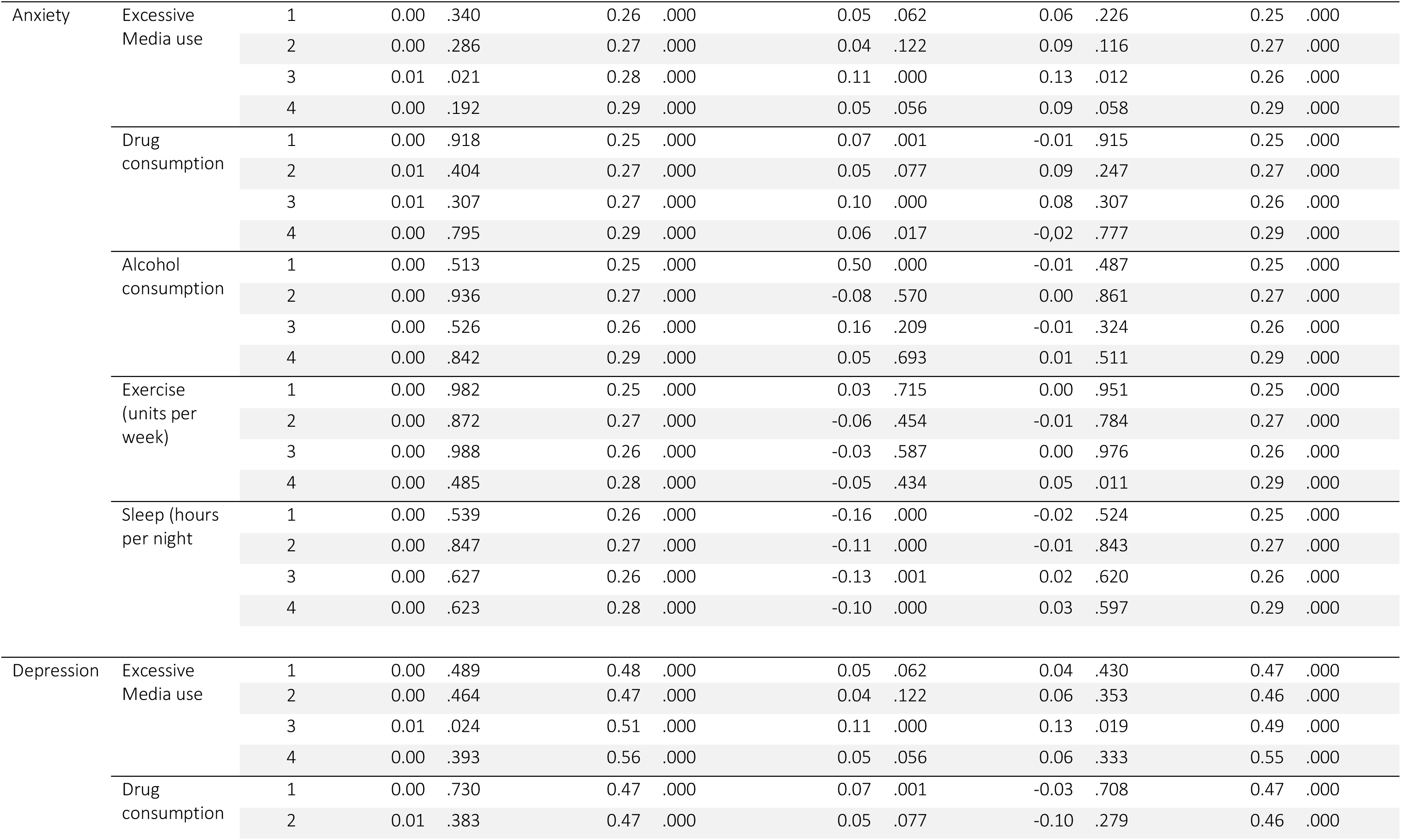
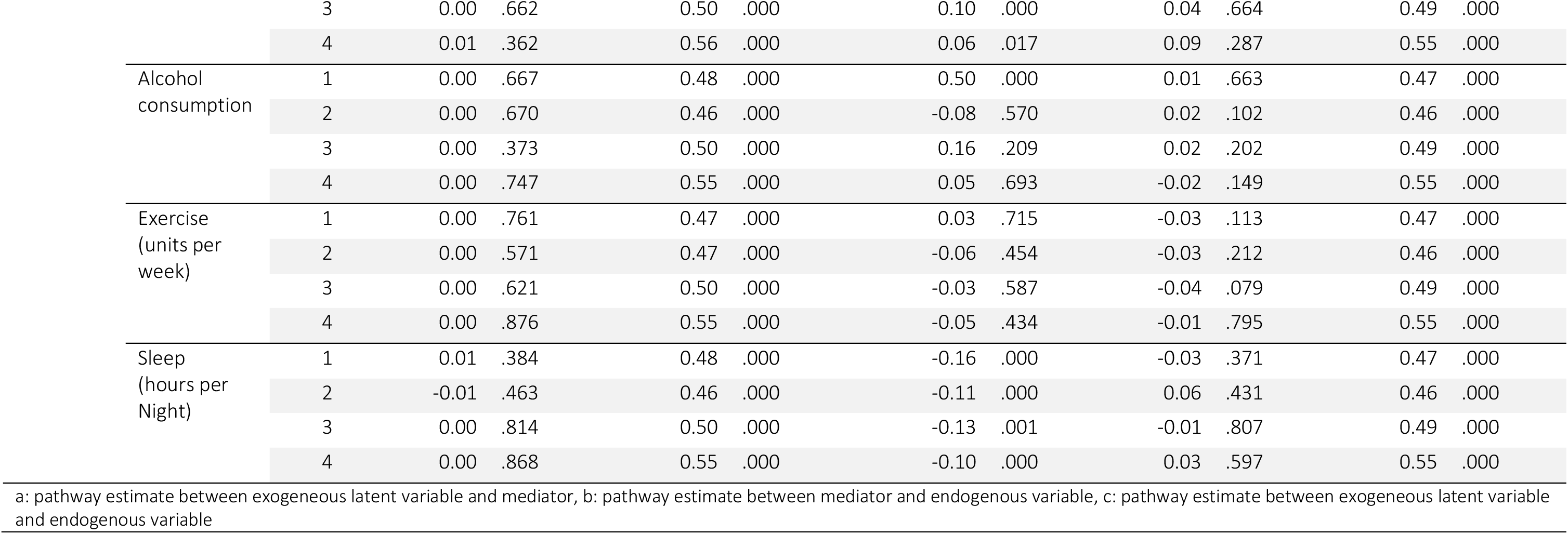
Overview of all results of the structural equation model with “social adversity” as a exogeneous latent variable for mental health endogenous variables with a mediation by harmful and healthy behaviors. The results are separated by endogenous variable (schizotypal traits, depressive symptoms, anxiety symptoms), mediators (excessive media use, drug consumption, alcohol consumption, units of exercise per week, hours of sleep per night during weeks) and samples at the four timepoints, showing indirect, direct and total effects, as well as individual pathways.

Investigating the effect of social adversity on anxiety (Figure 5, Table 5), structural equation models revealed that the exogeneous latent variable was associated with heightened anxiety (c=0.25 – c=0.29). All total effects were significant. The mediation with excessive media consumption revealed again a statistically significant indirect effect in the winter 2021 survey: 3.6% of the effects of social adversity during the pandemic on depressive symptoms were mediated by excessive media use. Again, there was no other statistically significant indirect effect, revealing no additional mediators.

Furthermore, SEM revealed that the exogeneous latent variable ‘social adversity’ had a positive association with depressive symptoms in all four surveys (Figure 5, Table 5). All total effects were significant. The mediation with excessive media consumption revealed again a statistically significant indirect effect in the winter 2021 survey, with 2% of effects explained.

#### 3.2.2. Exploratory model: Mediating effects of anxiety and depression on schizotypal traits

In an exploratory model, we investigated the mediating effect of anxiety and depression on the association between COVID-19 related life concerns and schizotypy, and social adversity and schizotypy. The model fit is presented in Supplementary Table 5. We found that the association between COVID-19 related life concerns and schizotypy was fully mediated by anxiety scores only in the winter 2021 and May 2021 surveys. Anxiety also mediated the effects of social adversity and schizotypy across all four surveys (Table 6). Depression did not act as a mediator between COVID-19 related life concerns and schizotypy, but it did mediate effects of social adversity predicting schizotypy in winter 2021.

**Table 6.**
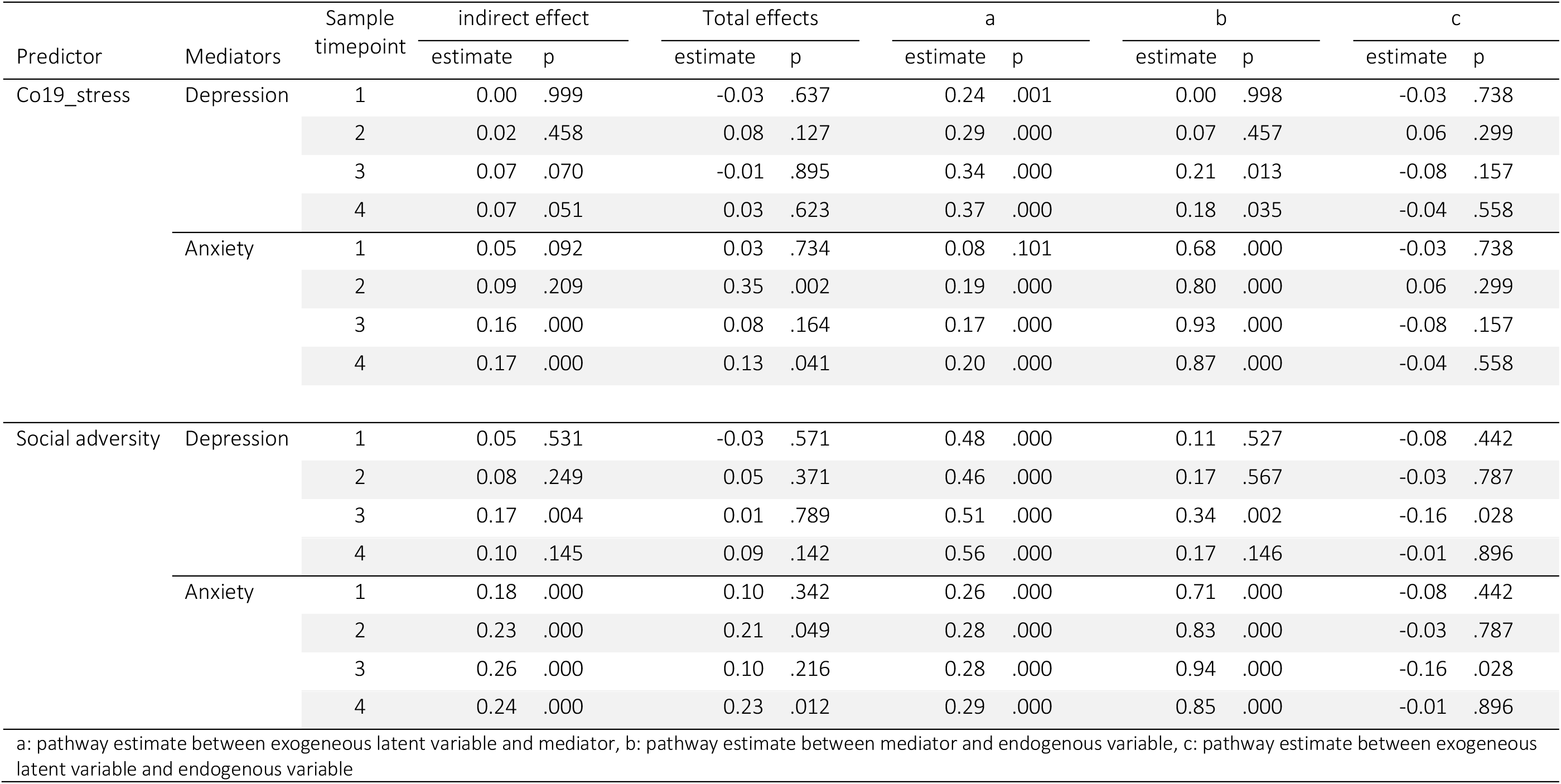
Exploratory analysis: Anxiety and Depression as Mediators and Schizotypy as Outcome.

| Predictor | Mediators | Sample<br>timepoint | indirect effect |  | Total effects |  | a |  | b |  | c |  |
| --- | --- | --- | --- | --- | --- | --- | --- | --- | --- | --- | --- | --- |
|  |  |  | estimate | p | estimate | p | estimate | p | estimate | p | estimate | p |
| Co19_stress | Depression | 1 | 0.00 | .999 | -0.03 | .637 | 0.24 | .001 | 0.00 | .998 | -0.03 | .738 |
|  |  | 2 | 0.02 | .458 | 0.08 | .127 | 0.29 | .000 | 0.07 | .457 | 0.06 | .299 |
|  |  | 3 | 0.07 | .070 | -0.01 | .895 | 0.34 | .000 | 0.21 | .013 | -0.08 | .157 |
|  |  | 4 | 0.07 | .051 | 0.03 | .623 | 0.37 | .000 | 0.18 | .035 | -0.04 | .558 |
|  | Anxiety | 1 | 0.05 | .092 | 0.03 | .734 | 0.08 | .101 | 0.68 | .000 | -0.03 | .738 |
|  |  | 2 | 0.09 | .209 | 0.35 | .002 | 0.19 | .000 | 0.80 | .000 | 0.06 | .299 |
|  |  | 3 | 0.16 | .000 | 0.08 | .164 | 0.17 | .000 | 0.93 | .000 | -0.08 | .157 |
|  |  | 4 | 0.17 | .000 | 0.13 | .041 | 0.20 | .000 | 0.87 | .000 | -0.04 | .558 |
| Social adversity | Depression | 1 | 0.05 | .531 | -0.03 | .571 | 0.48 | .000 | 0.11 | .527 | -0.08 | .442 |
|  |  | 2 | 0.08 | .249 | 0.05 | .371 | 0.46 | .000 | 0.17 | .567 | -0.03 | .787 |
|  |  | 3 | 0.17 | .004 | 0.01 | .789 | 0.51 | .000 | 0.34 | .002 | -0.16 | .028 |
|  |  | 4 | 0.10 | .145 | 0.09 | .142 | 0.56 | .000 | 0.17 | .146 | -0.01 | .896 |
|  | Anxiety | 1 | 0.18 | .000 | 0.10 | .342 | 0.26 | .000 | 0.71 | .000 | -0.08 | .442 |
|  |  | 2 | 0.23 | .000 | 0.21 | .049 | 0.28 | .000 | 0.83 | .000 | -0.03 | .787 |
|  |  | 3 | 0.26 | .000 | 0.10 | .216 | 0.28 | .000 | 0.94 | .000 | -0.16 | .028 |
|  |  | 4 | 0.24 | .000 | 0.23 | .012 | 0.29 | .000 | 0.85 | .000 | -0.01 | .896 |

a: pathway estimate between exogeneous latent variable and mediator, b: pathway estimate between mediator and endogenous variable, c: pathway estimate between exogeneous latent variable and endogenous variable

## 4. Discussion

In this paper, we investigated whether ‘social adversity’ and ‘COVID-19 related life concerns’ predicted schizotypal traits, anxiety and depression in snowball-ascertained samples of British and German participants at four time points throughout the pandemic. We furthermore explored whether these associations were mediated by healthy and unhealthy behaviours using a structural equation modelling approach.

### COVID-19 related life concerns’ model

Our results showed that COVID-19 related life concerns were associated with SPQ scores in autumn 2020 and May 2021, anxiety symptoms at all surveys except the first lockdown in March 2020, and depressive symptoms in all four surveys. Interestingly, COVID-19 related concerns were associated with schizotypal traits in those samples collected when the restrictions on daily life and the restrictive measures were less severe (autumn 2020 and May 2021 samples). This may be attributed to characteristics of a schizotypal personality. While some people may be less stressed due to lower restrictive measures and may quickly return to normality, others with more schizotypal traits may still be suspicious of the situation and unable to trust it (Preti et al., 2020), whereas they were more likely to feel secure when restrictions were in place, which is why effects were not seen during lockdowns.

COVID-19 related concerns, moreover, predicted an increase in media consumption, in the samples measured at the winter 2021 survey, confirming that individuals who experienced high stress related to COVID-19 may use excessive media consumption as a potential coping strategy (Bendau et al., 2021). Media consumption was also associated with increased schizotypy and depressive and anxiety symptoms in people experiencing high levels COVID-19 related concerns in the sample measured in winter 2021, when case numbers were highest in both countries. Here we found that media consumption significantly mediated the effect of COVID- 19 related life concerns on schizotypy and depressive and anxiety symptoms, potentially by increasing levels of fear and misinformation (Tasnim et al., 2020) and thus increasing these symptoms (Ni et al., 2020; Satici et al., 2020).

COVID-19 related life concerns also predicted drug consumption in the samples from autumn 2020, winter 2021 and May 2021, indicating the role of drugs as a coping strategy of burdened people (Czeisler et al., 2020). Furthermore, a positive correlation was found between COVID- 19 related life concerns and alcohol consumption only in the May 2021 survey. This could be due to the calming effect of alcohol on the nervous system, hence it could be expected that consumption might increase in response to stressors (Kypri & McCambridge, 2018; Eckardt et al., 1998). However, we did not find this effect at the three other surveys. Interestingly, general alcohol consumption declined in most of the European countries, with the exception of the UK (Kilian et al., 2021) during the pandemic.

Our first model also revealed that higher levels of COVID-19 related life concerns was associated with fewer hours of sleep at night in the winter 2021 and May 2021 surveys. Poor sleeping or poor sleeping patterns (e.g. <6h of sleep) are associated with worse mental health (Becker et al., 2018; João et al., 2018). Franceschini and colleagues (Franceschini et al., 2020) showed that 55% of 6519 adults reported poor sleep quality during the first lockdown, which was associated with higher levels of stress during the pandemic. Consistent with these findings, we found that more hours of sleep (>6h) were associated with lower levels of depression in the spring and autumn 2020 surveys, lower anxiety levels in the spring 2020 survey and lower SPQ scores in autumn 2020. Sleep may therefore play an important role as an emotional stabilizer (Goldstein & Walker, 2014).

We did not find a negative association between COVID related life concerns and physical activity; however, we found a positive association between physical activity and mental health scores; indicating that more physical activity is associated with higher anxiety in the autumn 2020 survey and spring 2021 survey. This is surprising and contradicts a large body of literature showing the positive effect of exercise on levels of anxiety, depression and stress (for review see (Mikkelsen et al., 2017)). As the associations, however, were found in those surveys when general restrictions were lower, more exercise potentially means an increased risk of exposure to potentially infected individuals. A potential reason for why we did not see a general positive effect might be the sensitivity of the measure (number of times of at least 30min increased heart rate activity per week). Also, individuals may have changed their exercise habits from for example team sport to individual exercise, which may dampen the positive impact.

### Social Adversity Model

In the second model, we explored investigated the association between social adversity or the lack of supportive relationships and mental health, and how those associations were mediated by healthy and unhealthy behaviour. We found that in all four surveys, social adversity predicted the expression of schizotypal traits. A study by Le et al. (2019) found that in individuals with trait schizotypy, psychosis-like symptoms were exacerbated only in those who experienced increased loneliness. Individuals high in schizotypal traits tend to have fewer social skills. Therefore an increase in loneliness during the pandemic might heighten schitzotypal traits, potentially pushing individuals with already high scores of schizotypy into a full blown psychosis (Brown et al., 2020; Chau et al., 2019; Esposito et al., 2021). Our findings in the light of the literature show the tremendous need for enabling safe social contacts during situations which require social distancing. Furthermore, we found that social stress predicted both depressive and anxiety symptoms in all four samples. This is consistent with the social exclusion theory, which identified social exclusion as the primary source of anxiety (Leary, 1990). Other studies during the COVID-19 pandemic have also shown that loneliness was the main risk factor for depression and anxiety (Okruszek et al., 2020; Palgi et al., 2020). McQuaid and colleagues (2021) found that loneliness related to anxiety and depression increased in a dose-like fashion during the first half of the COVID-19 pandemic.

In addition, social adversity was associated with increased excessive media use in the winter 2021 sample and increased drug use in sample at the surveys from spring 2020, winter 2021 and May 2021. Both have been discussed as maladaptive coping mechanisms to reduce loneliness (Rokach, 2005). Increased media consumption during the pandemic has been described as a compensation for the lack of social contacts (Cauberghe et al., 2021). However, excessive use of social media increases the risk of misinformation (Siddiqui et al., 2020; Tasnim et al., 2020), and as pointed out above is linked to lower mental health, especially to higher levels of anxiety and depression (Gao et al., 2020; Zhao & Zhou, 2020). Furthermore, higher social adversity also predicted higher alcohol consumption in the spring 2020 survey. This showed that people who felt more lonely or isolated at the beginning of the pandemic also drank more alcohol, as previously found (Horigian et al., 2021). Similar to the first model, social adversity was found to reduce sleep. Research on sleep, social adversity and mental health is inconsistent, with some studies reporting no influence of social adversity or loneliness on sleep (e.g.(Hawkley et al., 2010)), while others show a clear negative impact of social adversity and loneliness on sleep (e.g.(Bartoszek et al., 2020; Groarke et al., 2020)). Unhealthy behavioural patterns might act in a negative self-reinforcing way, as it has been reported that lack of sleep leads people to participate less in social interactions and to withdraw (Simon & Walker, 2018), increasing the risk for reduced mental health, which might again negatively impact the quality of sleep. However, our cross-sectional data does not show evidence of a direct link between reduced sleep, schizotypy, depressive or anxiety symptoms. We also found no association between social adversity and physical activity. This contradicts the results of other studies that report that loneliness decreased the likelihood of physical activity (Creese et al., 2021; Hawkley et al., 2009). However, we again found that more exercise was associated with higher levels of anxiety at the spring 2021 survey, which could be explained as.

Excessive media consumption partially mediated the effects of social adversity on SPQ traits. Thus, individuals who suffer more from loneliness or lack of social support have higher schizotypy in part because of their propensity for high levels of media consumption. However, we did not find mediation of the social adversity and schizotypy association via drugs, alcohol consumption, exercise, or sleep.

In exploratory analyses, we investigated the mediating effect of anxiety and depressive symptoms on the association between social adversity and schizotypal traits and COVID-19 related concerns and schizotypal traits. Depressive symptoms did not mediate the effects of COVID-19 concerns on schizotypy, but they did mediate effects of social adversity on schizotypy, but only during the winter 2021 survey. We found that the association between social adversity as well as COVID-19 related concerns and schizotypy was in both cases fully mediated by anxiety (all surveys for the former effects, only winter 2021 and May 2021 for the latter effects). This means that social adversity and COVID-19 related concerns increased anxiety in our samples which in turn impacted the level of schizotypal traits. Findings are consistent with other work reporting a positive association between schizotypy and social anxiety (Brown et al., 2008; Lewandowski et al., 2006).

There are several limitations to our study. First, in this analysis we investigate four different samples collected at different time points within one year from the start of the COVID-19 pandemic. Although the samples are highly comparable, and we adjust for observed differences between them in our models, we cannot adjust for unmeasured confounding. Thus, we cannot definitively say that the altered associations observed throughout the pandemic are related to genuine changes in these relationships, and thus reflect the changing impact of the pandemic. Second, an adequate fit is assumed for RMSEA of less than 0.06 and a CFI of greater than 0.90. Our models fulfil these criteria for the RMSEA, but are slightly lower for the CFI, ranging between 0.78-0.86. Our models are highly complex, with each model including five mediators and three outcomes. When reducing the complexity of the models, fit based on CFI meets threshold (≥0.9); however, for the sake of parsimony we present the full models.

In conclusion, we found that social adversity and COVID-19 related concerns predicted higher schizotypal traits, and symptoms of depression and anxiety. We furthermore found that excessive media consumption (more than 4h a day) partially mediated these relationships. We identified the ameliorating potential of regular sleep on mental health, but especially on schizotypy. Overall, our study shows that during the handling of extreme situations such as a global pandemic which require lockdowns and social distancing, sustained meaningful relationships and a healthy lifestyle are essential to maintain mental health. Furthermore, our findings highlight the risk of pandemic stressors heightening people’s levels of schizotypy, especially through excessive media consumption. Finally, our findings show that symptoms of common mental disorders (depression and anxiety) mediated the effects of social adversity and COVID-19 related concerns on schizotypy. All together our findings underscore the need for protective measures to be in place, such as social support networks, psychoeducation, media education, and treatment for common mental disorders, to support those most vulnerable to increased schizotypy.

## Data Availability

All data produced in the present study are available upon reasonable request to the authors.

## Acknowledgement

We thank the participants of the study who contributed their time and further circulated the survey.

## Data availability

All data produced in the present study are available upon reasonable request to the authors.

## Ethical statement

Ethical approval was obtained from the Ethical Commission Board of the Technical University Munich (250/20 S). All respondents included in the analyses provided informed consent.

## References

1. Abbas, J., Wang, D., Su, Z., & Ziapour, A. (2021). The role of social media in the advent of COVID-19 pandemic: crisis management, mental health challenges and implications. Risk Management and Healthcare Policy, 14, 1917.

2. Ahrens, K. F., Neumann, R. J., Kollmann, B., Plichta, M. M., Lieb, K., Tüscher, O., & Reif, A. (2021). Differential impact of COVID-related lockdown on mental health in Germany. World Psychiatry, 20(1), 140–141. https://doi.org/10.1002/wps.20830

3. Amsalem, D., Dixon, L. B., & Neria, Y. (2021). The coronavirus disease 2019 (COVID-19) outbreak and mental health: current risks and recommended actions. JAMA Psychiatry, 78(1), 9–10.

4. Balmford, B., Annan, J. D., Hargreaves, J. C., Altoè, M., & Bateman, I. J. (2020). Cross-Country Comparisons of Covid-19: Policy, Politics and the Price of Life. Environmental and Resource Economics, 1–27.

5. Barron, D., Morgan, K., Towell, T., Altemeyer, B., & Swami, V. (2014). Associations between schizotypy and belief in conspiracist ideation. Personality and Individual Differences, 70, 156–159. https://doi.org/https://doi.org/10.1016/j.paid.2014.06.040

6. Bartoszek, A., Walkowiak, D., Bartoszek, A., & Kardas, G. (2020). Mental well-being (depression, loneliness, insomnia, daily life fatigue) during COVID-19 related home-confinement—A study from Poland. International Journal of Environmental Research and Public Health, 17(20), 7417.

7. Bäuerle, A., Teufel, M., Musche, V., Weismüller, B., Kohler, H., Hetkamp, M., Dörrie, N., Schweda, A., & Skoda, E.-M. (2020). Increased generalized anxiety, depression and distress during the COVID-19 pandemic: a cross-sectional study in Germany. Journal of Public Health.

8. Beards, S., Gayer-Anderson, C., Borges, S., Dewey, M. E., Fisher, H. L., & Morgan, C. (2013). Life events and psychosis: a review and meta-analysis. Schizophrenia Bulletin, 39(4), 740–747.

9. Beauducel, A., & Wittmann, W. W. (2005). Simulation study on fit indexes in CFA based on data with slightly distorted simple structure. Structural Equation Modeling, 12(1), 41–75.

10. Becker, S. P., Jarrett, M. A., Luebbe, A. M., Garner, A. A., Burns, G. L., & Kofler, M. J. (2018). Sleep in a large, multi-university sample of college students: sleep problem prevalence, sex differences, and mental health correlates. Sleep Health, 4(2), 174–181.

11. Ben Simon, E., & Walker, M. P. (2018). Sleep loss causes social withdrawal and loneliness. Nature Communications, 9(1). https://doi.org/10.1038/s41467-018-05377-0

12. Bendau, A., Petzold, M. B., Pyrkosch, L., Maricic, L. M., Betzler, F., Rogoll, J., Große, J., Ströhle, A., & Plag, J. (2020). Associations between COVID-19 related media consumption and symptoms of anxiety, depression and COVID-19 related fear in the general population in Germany. European Archives of Psychiatry and Clinical Neuroscience, 1–9.

13. Bendau, A., Petzold, M. B., Pyrkosch, L., Mascarell Maricic, L., Betzler, F., Rogoll, J., Große, J., Ströhle, A., & Plag, J. (2021). Associations between COVID-19 related media consumption and symptoms of anxiety, depression and COVID-19 related fear in the general population in Germany. European Archives of Psychiatry and Clinical Neuroscience, 271(2), 283–291. https://doi.org/10.1007/s00406-020-01171-6

14. Bentler, P. M. (1990). Comparative fit indexes in structural models. Psychological Bulletin, 107(2), 238.

15. Brandmaier, A. (2021). Ωnyx - A graphical interface for Structural Equation Modeling.

16. Brooks, S. K., Webster, R. K., Smith, L. E., Woodland, L., Wessely, S., Greenberg, N., & Rubin, G. J. (2020). The psychological impact of quarantine and how to reduce it: rapid review of the evidence. The Lancet.

17. Brown, E., Gray, R., Lo Monaco, S., O’Donoghue, B., Nelson, B., Thompson, A., Francey, S., & McGorry, P. (2020). The potential impact of COVID-19 on psychosis: A rapid review of contemporary epidemic and pandemic research. Schizophrenia Research, 222, 79–87. https://doi.org/10.1016/j.schres.2020.05.005

18. Brown, L. H., Silvia, P. J., Myin–Germeys, I., Lewandowski, K. E., & Kwapil, T. R. (2008). The relationship of social anxiety and social anhedonia to psychometrically identified schizotypy. Journal of Social and Clinical Psychology, 27(2), 127–149.

19. Browne, M. W., & Cudeck, R. (1992). Alternative ways of assessing model fit. Sociological Methods & Research, 21(2), 230–258.

20. Bu, F., Steptoe, A., & Fancourt, D. (2020). Loneliness during a strict lockdown: Trajectories and predictors during the COVID-19 pandemic in 38,217 United Kingdom adults. Social Science and Medicine, 265(November), 113521. https://doi.org/10.1016/j.socscimed.2020.113521

21. Buchanan, T., & Kempley, J. (2021). Individual differences in sharing false political information on social media: Direct and indirect effects of cognitive-perceptual schizotypy and psychopathy. Personality and Individual Differences, 182, 111071.

22. Carter, C., Barch, D., Pearlstein, W., Baird, J., Baker, R., Cohen, J. D., & Schooler, N. (1995). A COGNITIVE NEUROPSYCHOLOGICAL STUDY OF SCHIZOPHRENIA SYMPTOMS - CORRELATES OF STROOP AND SEMANTIC PRIMING PERFORMANCE. SCHIZOPHRENIA RESEARCH, 15(1–2), 111–112.

23. Cauberghe, V., Van Wesenbeeck, I., De Jans, S., Hudders, L., & Ponnet, K. (2021). How Adolescents Use Social Media to Cope with Feelings of Loneliness and Anxiety during COVID-19 Lockdown. Cyberpsychology, Behavior, and Social Networking, 24(4), 250–257. https://doi.org/10.1089/cyber.2020.0478

24. Chapman, L. J., Chapman, J. P., Kwapil, T. R., Eckblad, M., & Zinser, M. C. (1994). Putatively psychosis-prone subjects 10 years later. Journal of Abnormal Psychology, 103(2), 171.

25. Chau, A. K. C., Zhu, C., & So, S. H.-W. (2019). Loneliness and the psychosis continuum: a meta-analysis on positive psychotic experiences and a meta-analysis on negative psychotic experiences. International Review of Psychiatry, 31(5–6), 471–490.

26. Chiappini, S., Guirguis, A., John, A., Corkery, J. M., & Schifano, F. (2020). COVID-19: The Hidden Impact on Mental Health and Drug Addiction. Frontiers in Psychiatry, 11(July), 10–13. https://doi.org/10.3389/fpsyt.2020.00767

27. Clay, J. M., & Parker, M. O. (2020). Alcohol use and misuse during the COVID-19 pandemic: a potential public health crisis? The Lancet Public Health, 5(5), e259. https://doi.org/10.1016/S2468-2667(20)30088-8

28. Cohen, J., & Kupferschmidt, K. (2020). Countries test tactics in “war” against COVID-19. Science, 367(6484), 1287–1288. https://doi.org/10.1126/science.367.6484.1287

29. Creese, B., Khan, Z., Henley, W., O’Dwyer, S., Corbett, A., Da Silva, M. V., Mills, K., Wright, N., Testad, I., & Aarsland, D. (2021). Loneliness, physical activity, and mental health during COVID-19: a longitudinal analysis of depression and anxiety in adults over the age of 50 between 2015 and 2020. International Psychogeriatrics, 33(5), 505–514.

30. Czeisler, M. É., Lane, R. I., Petrosky, E., Wiley, J. F., Christensen, A., Njai, R., Weaver, M. D., Robbins, R., Facer-Childs, E. R., Barger, L. K., Czeisler, C. A., Howard, M. E., & Rajaratnam, S. M. W. (2020). Mental Health, Substance Use, and Suicidal Ideation During the COVID- 19 Pandemic — United States, June 24–30, 2020. MMWR. Morbidity and Mortality Weekly Report, 69(32), 1049–1057. https://doi.org/10.15585/mmwr.mm6932a1

31. Daimer, S., Mihatsch, L., Ronan, L., Murray, G. K., & Knolle, F. (2021). Subjective impact of the COVID-19 pandemic on schizotypy and general mental health in Germany and the UK, for independent samples in May and in October 2020. Frontiers in Psychology, 12, 2875.

32. Daly, M., & Robinson, E. (2021). High-Risk Drinking in Midlife Before Versus During the COVID- 19 Crisis: Longitudinal Evidence From the United Kingdom. American Journal of Preventive Medicine, 60(2), 294–297. https://doi.org/10.1016/j.amepre.2020.09.004

33. De Coninck, D., Frissen, T., Matthijs, K., d’Haenens, L., Lits, G., Champagne-Poirier, O., Carignan, M.-E., David, M. D., Pignard-Cheynel, N., & Salerno, S. (2021). Beliefs in conspiracy theories and misinformation about COVID-19: Comparative perspectives on the role of anxiety, depression and exposure to and trust in information sources. Frontiers in Psychology, 12.

34. De Sousa, R. A. L., Improta-Caria, A. C., Aras-Júnior, R., de Oliveira, E. M., Soci, Ú. P. R., & Cassilhas, R. C. (2021). Physical exercise effects on the brain during COVID-19 pandemic: links between mental and cardiovascular health. Neurological Sciences, 1–10.

35. Debbané, M., & Barrantes-Vidal, N. (2015). Schizotypy from a developmental perspective. Schizophrenia Bulletin, 41(suppl_2), S386–S395.

36. ECDE. (2021). Overview of the implementation of COVID-19 vaccination strategies and deployment plans in the EU/EEA.

37. Esposito, C. M., D’Agostino, A., Osso, B. D., Fiorentini, A., Prunas, C., Callari, A., Oldani, L., Fontana, E., Gargano, G., & Viscardi, B. (2021). Impact of the first Covid-19 pandemic wave on first episode psychosis in Milan, italy. Psychiatry Research, 298, 113802.

38. Fancourt, D., Steptoe, A., & Bu, F. (2020). Trajectories of anxiety and depressive symptoms during enforced isolation due to COVID-19 in England: a longitudinal observational study. The Lancet Psychiatry.

39. Fekih-Romdhane, F., Dissem, N., & Cheour, M. (2021). How did Tunisian university students cope with fear of COVID-19? A comparison across schizotypy features. Personality and Individual Differences, 178(March), 110872. https://doi.org/10.1016/j.paid.2021.110872

40. Finlay, I., & Gilmore, I. (2020). Covid-19 and alcohol-a dangerous cocktail. The BMJ, 369(May), 2–3. https://doi.org/10.1136/bmj.m1987

41. Firth, J., Carney, R., Elliott, R., French, P., Parker, S., McIntyre, R., McPhee, J. S., & Yung, A. R. (2018). Exercise as an intervention for first-episode psychosis: a feasibility study. Early Intervention in Psychiatry, 12(3), 307–315.

42. Franceschini, C., Musetti, A., Zenesini, C., Palagini, L., Scarpelli, S., Quattropani, M. C., Lenzo, V., Freda, M. F., Lemmo, D., & Vegni, E. (2020). Poor sleep quality and its consequences on mental health during the COVID-19 lockdown in Italy. Frontiers in Psychology, 11, 3072.

43. Gao, J., Zheng, P., Jia, Y., Chen, H., Mao, Y., Chen, S., Wang, Y., Fu, H., & Dai, J. (2020). Mental health problems and social media exposure during COVID-19 outbreak. Plos One, 15(4), e0231924.

44. Garfin, D. R., Silver, R. C., & Holman, E. A. (2020). The novel coronavirus (COVID-2019) outbreak: Amplification of public health consequences by media exposure. Health Psychology.

45. Goldstein, A. N., & Walker, M. P. (2014). The role of sleep in emotional brain function. Annual Review of Clinical Psychology, 10, 679–708. https://doi.org/10.1146/annurev-clinpsy-032813-153716

46. Grant, P., Green, M. J., & Mason, O. J. (2018). Models of schizotypy: the importance of conceptual clarity. Schizophrenia Bulletin, 44(suppl_2), S556–S563.

47. Grant, P., & Hennig, J. (2020). Schizotypy, social stress and the emergence of psychotic-like states - A case for benign schizotypy? Schizophrenia Research, 216, 435–442. https://doi.org/10.1016/j.schres.2019.10.052

48. Groarke, J. M., Berry, E., Graham-Wisener, L., McKenna-Plumley, P. E., McGlinchey, E., & Armour, C. (2020). Loneliness in the UK during the COVID-19 pandemic: Cross-sectional results from the COVID-19 Psychological Wellbeing Study. PloS One, 15(9), e0239698.

49. Hardt, J., Dragan, M., & Kappis, B. (2011). A short screening instrument for mental health problems: The Symptom Checklist-27 (SCL-27) in Poland and Germany. International Journal of Psychiatry in Clinical Practice, 15(1), 42–49.

50. Hardt, J., & Gerbershagen, H. U. (2001). Cross-validation of the SCL-27: A short psychometric screening instrument for chronic pain patients. European Journal of Pain, 5(2), 187–197.

51. Hawkley, L. C., Preacher, K. J., & Cacioppo, J. T. (2010). Loneliness impairs daytime functioning but not sleep duration. Health Psychology, 29(2), 124.

52. Hawkley, L. C., Thisted, R. A., & Cacioppo, J. T. (2009). Loneliness predicts reduced physical activity: cross-sectional & longitudinal analyses. Health Psychology, 28(3), 354.

53. Horigian, V. E., Schmidt, R. D., & Feaster, D. J. (2021). Loneliness, mental health, and substance use among US young adults during COVID-19. Journal of Psychoactive Drugs, 53(1), 1–9.

54. Hu, W., Su, L., Qiao, J., Zhu, J., & Zhou, Y. (2020). COVID-19 outbreak increased risk of schizophrenia in aged adults. Chinaxiv. Org (Preprint) Web Resource Https://Scholar. Google. Com/Scholar_lookup.

55. Jacob, L., Smith, L., Armstrong, N. C., Yakkundi, A., Barnett, Y., Butler, L., McDermott, D. T., Koyanagi, A., Shin, J. Il, Meyer, J., Firth, J., Remes, O., López-Sánchez, G. F., & Tully, M. A. (2021). Alcohol use and mental health during COVID-19 lockdown: A cross-sectional study in a sample of UK adults. Drug and Alcohol Dependence, 219(November 2020). https://doi.org/10.1016/j.drugalcdep.2020.108488

56. JHU. (2021). COVID-19 Dashboard by the Center for Systems Science and Engineering (CSSE) at Johns Hopkins University (JHU). https://coronavirus.jhu.edu/map.html

57. João, K. A. D. R., de Jesus, S. N., Carmo, C., & Pinto, P. (2018). The impact of sleep quality on the mental health of a non-clinical population. Sleep Medicine, 46, 69–73.

58. Katikireddi, S. V., Whitley, E., Lewsey, J., Gray, L., & Leyland, A. H. (2017). Socioeconomic status as an effect modifier of alcohol consumption and harm: analysis of linked cohort data. The Lancet Public Health, 2(6), e267–e276. https://doi.org/10.1016/S2468-2667(17)30078-6

59. Kilian, C., Rehm, J., Allebeck, P., Braddick, F., Gual, A., Barták, M., Bloomfield, K., Gil, A., Neufeld, M., O’Donnell, A., Petruželka, B., Rogalewicz, V., Schulte, B., & Manthey, J. (2021). Alcohol consumption during the COVID-19 pandemic in Europe: a large-scale cross-sectional study in 21 countries. Addiction (Abingdon, England). https://doi.org/10.1111/add.15530

60. Knolle, F., Ronan, L., & Murray, G. K. (2021). The impact of the COVID-19 pandemic on mental health in the general population: a comparison between Germany and the UK. BMC Psychology, 9(1), 1–17.

61. Kozloff, N., Mulsant, B. H., Stergiopoulos, V., & Voineskos, A. N. (2020). The COVID-19 global pandemic: implications for people with schizophrenia and related disorders. Schizophrenia Bulletin, 46(4), 752–757.

62. Kuhl, H. C., Hartwig, I., Petitjean, S., Müller-Spahn, F., Margraf, J., & Bader, K. (2010). Validation of the Symptom Checklist SCL-27 in psychiatric patients: Psychometric testing of a multidimensional short form. International Journal of Psychiatry in Clinical Practice, 14(2), 145–149. https://doi.org/10.3109/13651501003660484

63. Kwapil, T. R., Gross, G. M., Burgin, C. J., Raulin, M. L., Silvia, P. J., & Barrantes-Vidal, N. (2018). Validity of the Multidimensional Schizotypy Scale: Associations with schizotypal traits and normal personality. Personality Disorders, 9(5), 458–466. https://doi.org/10.1037/per0000288

64. Kypri, K., & McCambridge, J. (2018). Alcohol must be recognised as a drug. BMJ, 362. https://doi.org/10.1136/bmj.k3944

65. Le, T. P., Cowan, T., Schwartz, E. K., Elvevåg, B., Holmlund, T. B., Foltz, P. W., Barkus, E., & Cohen, A. S. (2019). The importance of loneliness in psychotic-like symptoms: Data from three studies. Psychiatry Research, 282, 112625.

66. Leary, M. R. (1990). Responses to Social Exclusion: Social Anxiety, Jealousy, Loneliness, Depression, and Low Self-Esteem. Journal of Social and Clinical Psychology, 9(2), 221– 229. https://doi.org/10.1521/jscp.1990.9.2.221

67. Lenzenweger, M. F. (2011). Schizotypy and schizophrenia: The view from experimental psychopathology. Guilford Press.

68. Lewandowski, K. E., Barrantes-Vidal, N., Nelson-Gray, R. O., Clancy, C., Kepley, H. O., & Kwapil, T, R. (2006). Anxiety and depression symptoms in psychometrically identified schizotypy. Schizophrenia Research, 83(2–3), 225–235.

69. Linscott, R. J., & Morton, S. E. (2018). The Latent Taxonicity of Schizotypy in Biological Siblings of Probands With Schizophrenia. Schizophrenia Bulletin, 44(4), 922–932. https://doi.org/10.1093/schbul/sbx143

70. Lippke, S., Fischer, M. A., & Ratz, T. (2021). Physical activity, loneliness, and meaning of friendship in young individuals–a mixed-methods investigation prior to and during the COVID-19 pandemic with three cross-sectional studies. Frontiers in Psychology, 12, 146.

71. Mair, P., & Wilcox, R. (2020). Robust statistical methods in R using the WRS2 package. Behavior Research Methods, 52(2), 464–488. https://doi.org/10.3758/s13428-019-01246-w

72. Manthey, J., Kilian, C., Carr, S., Bartak, M., Bloomfield, K., Braddick, F., Gual, A., Neufeld, M., O’Donnell, A., Petruzelka, B., Rogalewicz, V., Rossow, I., Schulte, B., & Rehm, J. (2021). Use of alcohol, tobacco, cannabis, and other substances during the first wave of the SARS-CoV-2 pandemic in Europe: a survey on 36,000 European substance users. Substance Abuse Treatment, Prevention, and Policy, 16(1), 36. https://doi.org/10.1186/s13011-021-00373-y

73. Marsden, J., Darke, S., Hall, W., Hickman, M., Holmes, J., Humphreys, K., Neale, J., Tucker, J., & West, R. (2020). Mitigating and learning from the impact of COVID-19 infection on addictive disorders. Addiction, 115(6), 1007–1010. https://doi.org/10.1111/add.15080

74. McKay, D., & Asmundson, G. J. G. (2020). COVID-19 stress and substance use: Current issues and future preparations. Journal of Anxiety Disorders, 74, 102274. https://doi.org/10.1016/j.janxdis.2020.102274

75. McQuaid, R. J., Cox, S. M. L., Ogunlana, A., & Jaworska, N. (2021). The burden of loneliness: Implications of the social determinants of health during COVID-19. Psychiatry Research, 296, 113648.

76. Mikkelsen, K., Stojanovska, L., Polenakovic, M., Bosevski, M., & Apostolopoulos, V. (2017). Exercise and mental health. Maturitas, 106, 48–56.

77. Mittal, V. A., Vargas, T., Osborne, K. J., Dean, D., Gupta, T., Ristanovic, I., Hooker, C. I., & Shankman, S. A. (2017). Exercise treatments for psychosis: a review. Current Treatment Options in Psychiatry, 4(2), 152–166.

78. Neophytou, E., Manwell, L. A., & Eikelboom, R. (2021). Effects of Excessive Screen Time on Neurodevelopment, Learning, Memory, Mental Health, and Neurodegeneration: a Scoping Review. International Journal of Mental Health and Addiction, 19(3), 724–744. https://doi.org/10.1007/s11469-019-00182-2

79. Ni, M. Y., Yang, L., Leung, C. M. C., Li, N., Yao, X. I., Wang, Y., Leung, G. M., Cowling, B. J., & Liao, Q. (2020). Mental health, risk factors, and social media use during the COVID-19 epidemic and cordon sanitaire among the community and health professionals in Wuhan, China: cross-sectional survey. JMIR Mental Health, 7(5), e19009.

80. Okruszek, Ł., Aniszewska-Stańczuk, A., Piejka, A., Wiśniewska, M., & Żurek, K. (2020). Safe but lonely? Loneliness, anxiety, and depression symptoms and COVID-19. Frontiers in Psychology, 11, 3222.

81. Palgi, Y., Shrira, A., Ring, L., Bodner, E., Avidor, S., Bergmand, Y., Cohen-Fridel, S., Keisari, S., & Hoffman, Y. (2020). The loneliness pandemic: Loneliness and other concomitants of depression, anxiety and their comorbidity during the COVID-19 outbreak. Journal of Affective Disorders, 275.

82. Pfefferbaum, B., & North, C. S. (2020). Mental health and the Covid-19 pandemic. New England Journal of Medicine.

83. Plümper, T., & Neumayer, E. (2020). Lockdown policies and the dynamics of the first wave of the Sars-CoV-2 pandemic in Europe. Journal of European Public Policy. https://doi.org/10.1080/13501763.2020.1847170

84. Popov, S., Sokic, J., & Stupar, D. (2021). Activity matters: Physical exercise and stress coping during the 2020 COVID-19 state of emergency. Psihologija, 54(3), 307–322. https://doi.org/10.2298/psi200804002p

85. Preti, E., Di Pierro, R., Fanti, E., Madeddu, F., & Calati, R. (2020). Personality Disorders in Time of Pandemic. Current Psychiatry Reports, 22(12), 1–9.

86. Proto, E., & Quintana-Domeque, C. (2021). COVID-19 and mental health deterioration by ethnicity and gender in the UK. Plos One, 16(1), e0244419. https://doi.org/10.1371/journal.pone.0244419

87. Raine, A. (1991). The SPQ: A Scale for the Assessment of Schizotypal Personality Based on DSM-III-R Criteria. Schizophrenia Bulletin, 17(4), 555–564. https://doi.org/10.1093/schbul/17.4.555

88. Rokach, A. (2005). Drug withdrawal and coping with loneliness. Social Indicators Research, 73(1), 71–85. https://doi.org/10.1007/s11205-004-2008-y

89. Rosseel, Y. (2012). Lavaan: An R package for structural equation modeling and more. Version 0.5–12 (BETA). Journal of Statistical Software, 48(2), 1–36.

90. Rstudio, T. (2020). RStudio: Integrated Development for R. In Rstudio Team, PBC, Boston, MA *URL* http://www.rstudio.com/. https://doi.org/10.1145/3132847.3132886

91. Sallie, S. N., Ritou, V., Bowden-Jones, H., & Voon, V. (2020). Assessing international alcohol consumption patterns during isolation from the COVID-19 pandemic using an online survey: highlighting negative emotionality mechanisms. BMJ Open, 10(11), e044276. https://doi.org/10.1136/bmjopen-2020-044276

92. Sasse, T. (2021). EU vaccine debate highlights UK success – and need for diplomacy.

93. Satici, B., Saricali, M., Satici, S. A., & Griffiths, M. D. (2020). Intolerance of Uncertainty and Mental Wellbeing: Serial Mediation by Rumination and Fear of COVID-19. International Journal of Mental Health and Addiction. https://doi.org/10.1007/s11469-020-00305-0

94. Siddiqui, M. Y. A., Mushtaq, K., Mohamed, M. F. H., Al Soub, H., Mohamedali, M. G. H., & Yousaf, Z. (2020). “Social media misinformation”—An epidemic within the COVID-19 pandemic. The American Journal of Tropical Medicine and Hygiene, 103(2), 920.

95. Smith, L., Jacob, L., Yakkundi, A., McDermott, D., Armstrong, N. C., Barnett, Y., López-Sánchez, G. F., Martin, S., Butler, L., & Tully, M. A. (2020). Correlates of symptoms of anxiety and depression and mental wellbeing associated with COVID-19: a cross-sectional study of UK-based respondents. Psychiatry Research, 291(May), 113138. https://doi.org/10.1016/j.psychres.2020.113138

96. Steiger, J. H. (1990). Structural model evaluation and modification: An interval estimation approach. Multivariate Behavioral Research, 25(2), 173–180.

97. Su, Z., McDonnell, D., Wen, J., Kozak, M., Abbas, J., Šegalo, S., Li, X., Ahmad, J., Cheshmehzangi, A., & Cai, Y. (2021). Mental health consequences of COVID-19 media coverage: the need for effective crisis communication practices. Globalization and Health, 17(1), 1–8.

98. Tasnim, S., Hossain, M. M., & Mazumder, H. (2020). Impact of rumors and misinformation on COVID-19 in social media. Journal of Preventive Medicine and Public Health, 53(3), 171–174.

99. Taylor, S. (2019). The Psychology of Pandemics.

100. Taylor, S., Paluszek, M. M., Rachor, G. S., McKay, D., & Asmundson, G. J. G. (2021). Substance use and abuse, COVID-19-related distress, and disregard for social distancing: A network analysis. Addictive Behaviors, 114(September 2020), 106754. https://doi.org/10.1016/j.addbeh.2020.106754

101. Team, R. C. (2016). R: A Language and Environment for Statistical Computing. In R Foundation for Statistical Computing.

102. Valdés-Florido, M. J., López-Díaz, Á., Palermo-Zeballos, F. J., Martínez-Molina, I., Martín-Gil, V. E., Crespo-Facorro, B., & Ruiz-Veguilla, M. (2020). Reactive psychoses in the context of the COVID-19 pandemic: clinical perspectives from a case series. Revista de Psiquiatría y Salud Mental (English Edition), 13(2), 90–94.

103. Valdez, D., Ten Thij, M., Bathina, K., Rutter, L. A., & Bollen, J. (2020). Social media insights into US mental health during the COVID-19 pandemic: longitudinal analysis of twitter data. Journal of Medical Internet Research, 22(12), e21418.

104. Wang, C., Pan, R., Wan, X., Tan, Y., Xu, L., McIntyre, R. S., Choo, F. N., Tran, B., Ho, R., & Sharma, V. K. (2020). A longitudinal study on the mental health of general population during the COVID-19 epidemic in China. Brain, Behavior, and Immunity.

105. Wang, Y., Gao, J. L., Chen, H., Mao, Y. M., Chen, S. H., Dai, J. M., Zheng, P. P., & Fu, H. (2020). The relationship between media exposure and mental health problems during COVID-19 outbreak. Fudan University Journal of Medical Sciences, 47(2).

106. WHO. (2020). Infodemic.

107. Zendle, D. (2020). Too much of a good thing? Excessive use across behaviours and associations with mental health. https://doi.org/10.31234/osf.io/pnx2e

108. Zhang, Y., Zhang, H., Ma, X., & Di, Q. (2020). Mental health problems during the COVID-19 pandemics and the mitigation effects of exercise: A longitudinal study of college students in China. International Journal of Environmental Research and Public Health, 17(10). https://doi.org/10.3390/ijerph17103722

109. Zhao, N., & Zhou, G. (2020). Social media use and mental health during the COVID-19 pandemic: Moderator role of disaster stressor and mediator role of negative affect. Applied Psychology: Health and Well-Being, 12(4), 1019–1038.

110. Zhong, B., Jiang, Z., Xie, W., & Qin, X. (2020). Association of social media use with mental health conditions of nonpatients during the COVID-19 Outbreak: Insights from a national survey study. Journal of Medical Internet Research, 22(12), e23696.

