## Supplementary Materials for "The relationship of COVID-19 related stress and media consumption with schizotypy, depression and anxiety"

### 1 Cohort demographics and COVID-19 exposure divided by country and survey

| **Suppl. Table 1. Cohort demographics and COVID-19 exposure divided by country and timepoint.** Values in percent if not indicated otherwise | | | | | | | | | | | | | | | | | | | | | | | | | |
| --- | --- | --- | --- | --- | --- | --- | --- | --- | --- | --- | --- | --- | --- | --- | --- | --- | --- | --- | --- | --- | --- | --- | --- | --- | --- |
|  | |  | | **Survey 1**  April/May 2020 | | **Survey 2**  Sept/Oct 2020 | | **Survey 3**  Jan/ Feb 2021 | | **Survey 4**  May 2021 | |  | **Country comparison** | | | | | |  | | | **Survey comparison** | | | |
|  | |  | | **UK** | **Ger** | **UK** | **Ger** | **UK** | **Ger** | **UK** | **Ger** | ***W/X^2^*** | | **df** | | | **p** | **K/*X^2^*** | | | | | **df** | | **p** |
| N | |  | | 239 | 542 | 115 | 383 | 96 | 448 | 70 | 416 |  | |  | |  | | | | 55.31 | | | 3 | .000 | |
| Percent | |  | | 27.9 | 63.1 | 21.1 | 70.4 | 16.3 | 76.1 | 13.7 | 81.6 |  | |  | |  | | | |  | | |  |  | |
| Age | | Mean | | 39.01 | 45.36 | 40.9 | 42.87 | 42.05 | 43.18 | 44.04 | 43.84 | 398256 | | | .000 | | | | | | 2.85 | | 3 | | .416 |
|  | | SD | | 16.02 | 14.87 | 16.17 | 16.04 | 17.38 | 14.45 | 18.73 | 14.64 |  | |  | |  | | | |  | | |  |  | |
| Gender | | Male | | 24.3 | 25.9 | 29.6 | 25.1 | 21.9 | 29.7 | 22.9 | 24.5 | 0.69 | | 2 | | .709 | | | | 6.76 | | | 6 | .343 | |
|  |  | Female | | 73.6 | 71.2 | 68.7 | 72.8 | 75.0 | 68.5 | 74.3 | 74.8 |  | |  | |  | | | |  | | |  |  | |
|  | | other/  n.a. | | 2.1 | 3.0 | 1.7 | 2.1 | 3.1 | 1.8 | 2.9 | 0.7 |  | |  | |  | | | |  | | |  |  | |
| Education | | School  leavers | | 0.4 | 0.0 | 0.0 | 0.0 | 0.0 | 0.0 | 0.0 | 0.2 | 194.74 | | 8 | | .000 | | | | 125.74 | | | 24 | .000 | |
|  |  | 8-years | | 19.2 | 13.1 | 16.5 | 13.8 | 17.7 | 25.9 | 5.7 | 20.9 |  | |  | |  | | | |  | | |  |  | |
|  | | Prof. college | | 31.8 | 21.6 | 39.1 | 30.8 | 49.0 | 32.6 | 44.3 | 34.1 |  | |  | |  | | | |  | | |  |  | |
|  | | Master or < | | 47.3 | 64.9 | 43.5 | 55.1 | 33.0 | 41.3 | 48.6 | 44.7 |  | |  | |  | | | |  | | |  |  | |
|  | | Missing | | 1.3 | 0.4 | 0.9 | 0.3 | 0.0 | 0.2 | 1.4 | 0.0 |  | |  | |  | | | |  | | |  |  | |
| Living Area | | City | | 20.50 | 60.30 | 25.20 | 43.30 | 24.00 | 37.10 | 20.00 | 47.40 | 147.64 | | 5 | | .000 | | | | 52.74 | | | 15 | .000 | |
|  |  | Suburb | | 7.90 | 13.10 | 12.20 | 13.80 | 14.60 | 11.80 | 14.30 | 10.10 |  | |  | |  | | | |  | | |  |  | |
|  | | Town | | 36.40 | 10.70 | 26.10 | 14.10 | 15.60 | 18.30 | 35.70 | 14.20 |  | |  | |  | | | |  | | |  |  | |
|  | | Village,   rural | | 34.70 | 15.70 | 35.70 | 26.40 | 44.80 | 31.50 | 30.00 | 26.00 |  | |  | |  | | | |  | | |  |  | |
|  | | Missing | | 0.40 | 0.20 | 0.90 | 2.30 | 1.00 | 1.30 | 0.00 | 2.40 |  | |  | |  | | | |  | | |  |  | |
| Rating physical  health before Co19 | | Exc. | | 13.00 | 11.30 | 8.70 | 10.70 | 13.50 | 15.80 | 8.60 | 13.90 | 28.49 | | 5 | | .000 | | | | 29.62 | | | 15 | .013 | |
|  |  | Very good | | 32.20 | 33.30 | 25.20 | 32.90 | 27.10 | 36.20 | 34.30 | 38.20 |  | |  | |  | | | |  | | |  |  | |
|  |  | Good | | 31.80 | 35.70 | 42.60 | 42.00 | 33.30 | 34.60 | 34.30 | 35.10 |  | |  | |  | | | |  | | |  |  | |
|  |  | Fairly | | 17.20 | 15.50 | 18.30 | 110 | 15.60 | 9.60 | 17.10 | 9.10 |  | |  | |  | | | |  | | |  |  | |
|  |  | Poor | | 3.80 | 3.30 | 4.30 | 1.60 | 10.40 | 1.80 | 5.70 | 1.70 |  | |  | |  | | | |  | | |  |  | |
|  |  | Missing | | 2.10 | 0.90 | 0.90 | 1.80 | 0.00 | 2.00 | 0.00 | 1.90 |  | |  | |  | | | |  | | |  |  | |
| Treatment  physical illness | | No | | 66.90 | 72.80 | 71.30 | 81.50 | 72.90 | 76.10 | 75.70 | 76.20 | 26.27 | | 2 | | .000 | | | | 101.25 | | | 6 | .000 | |
|  |  | Yes | | 18.40 | 20.10 | 27.00 | 17.80 | 260 | 22.80 | 22.90 | 23.10 |  | |  | |  | | | |  | | |  |  | |
|  |  | Missing | | 14.60 | 7.00 | 1.70 | 0.80 | 1.00 | 1.10 | 1.40 | 0.70 |  | |  | |  | | | |  | | |  |  | |
| Rating mental  health before  Co19 | | Exc. | | 14.20 | 15.30 | 8.70 | 11.50 | 15.60 | 17.90 | 15.70 | 15.10 | 87.86 | | 5 | | .000 | | | | 41.85 | | | 15 | .000 | |
|  |  | Very good | | 21.80 | 37.00 | 23.50 | 33.40 | 27.10 | 40.80 | 25.70 | 42.10 |  | |  | |  | | | |  | | |  |  | |
|  |  | Good | | 30.10 | 28.30 | 38.30 | 37.90 | 28.10 | 28.30 | 32.90 | 27.20 |  | |  | |  | | | |  | | |  |  | |
|  |  | Fairly | | 21.30 | 14.60 | 18.30 | 12.30 | 16.70 | 9.40 | 14.30 | 11.80 |  | |  | |  | | | |  | | |  |  | |
|  |  | Poor | | 11.30 | 3.30 | 9.60 | 3.90 | 10.40 | 2.50 | 11.40 | 2.90 |  | |  | |  | | | |  | | |  |  | |
|  |  | Missing | | 1.30 | 1.50 | 1.70 | 1.00 | 2.10 | 1.10 | 0.00 | 1.00 |  | |  | |  | | | |  | | |  |  | |
| Psych. Treatment | | No | | 69.00 | 81.90 | 79.10 | 89.60 | 78.10 | 86.40 | 80.00 | 87.70 | 41.63 | | 2 | | .000 | | | | 92.40 | | | 6 | .000 | |
|  |  | Yes | | 17.60 | 10.90 | 18.30 | 9.70 | 20.80 | 12.70 | 20.00 | 10.80 |  | |  | |  | | | |  | | |  |  | |
|  |  | Missing | | 13.40 | 7.20 | 2.60 | 0.80 | 1.00 | 0.90 | 0.00 | 1.40 |  | |  | |  | | | |  | | |  |  | |
| Suspected  infection | | Positive Test | | 0.00 | 0.20 | 0.00 | 0.80 | 5.20 | 3.10 | 1.40 | 3.80 | 41.76 | | 12 | | .004 | | | | 41.76 | | | 12 | .000 | |
|  |  | Diagn. | | 2.50 | 0.70 | 0.90 | 0.50 | 1.00 | 0.90 | 0.00 | 0.50 |  | |  | |  | | | |  | | |  |  | |
|  |  | Sympt. | | 18.80 | 14.20 | 19.10 | 15.40 | 21.90 | 16.50 | 28.60 | 14.70 |  | |  | |  | | | |  | | |  |  | |
|  |  | No inf. | | 78.70 | 83.90 | 79.10 | 82.50 | 71.90 | 79.20 | 70.00 | 80.80 |  | |  | |  | | | |  | | |  |  | |
|  |  | Missing | | 0.00 | 0.90 | 0.90 | 0.80 | 0.00 | 0.20 | 0.00 | 0.20 |  | |  | |  | | | |  | | |  |  | |
|  |  | | GER: German sample, UK: UK sample, TP: time point, p: p-vlaue, n.a.: missing values, Exc.. Excellent, Diagn: positive Diagnosis, Sympt: Symptoms of COVID-19, No inf.: No infection | | | | | | | | | | | | | | | | | | | | | | |

### 2 Pearson’s correlation of anxiety and SPQ scores and depression and SPQ scores

| **Suppl. Table 2. Pearson’s correlation coefficient between SPQ sumscore and anxiety score and SPQ sumscore (SCL-27) and depression score (SCL-27)** | | |
| --- | --- | --- |
| Survey | SPQ &  Anxiety | SPQ &  Depression |
| 1 - April/May 20 | 0.57 *** | 0.53 *** |
| 2 - Sept./ Oct. 20 | 0.68 *** | 0.58 *** |
| 3 - Jan. /Feb. 21 | 0.67 *** | 0.56 *** |
| 4 - May 21 | 0.66 *** | 0.56 *** |
| *** = p <.001 | | |

### 3 Model fit predictor measurement models

**Suppl. Table 3. Overview of the model fit indices for predictor models separate by timepoint**

|  |  |  |  | exact modelfit |  | relativ modelfit | | absolute modelfit | |
| --- | --- | --- | --- | --- | --- | --- | --- | --- | --- |
|  |  | Teststatistic | DF | *X^2^* |  | CFI |  | | RMSEA |
| Predictor | Timepoint |  |  |  |  |  |  | |  |
| COVID-19 related life concerns | 1 | 163.73 | 3 | .000 |  | 1.0 |  | | 0.0 |
|  | 2 | 201.93 | 3 | .000 |  | 1.0 |  | | 0.0 |
|  | 3 | 326.03 | 3 | .000 |  | 1.0 |  | | 0.0 |
|  | 4 | 242.49 | 3 | .000 |  | 1.0 |  | | 0.0 |
| Social adversity | 1 | 269.48 | 3 | .000 |  | 1.0 |  | | 0.0 |
|  | 2 | 258.3 | 3 | .000 |  | 1.0 |  | | 0.0 |
|  | 3 | 364.4 | 3 | .000 |  | 1.0 |  | | 0.0 |
|  | 4 | 335.31 | 3 | .000 |  | 1.0 |  | | 0.0 |
| DF: degree of freedom, *X^2^*: Chi squared test, CFI: comparative fit index, TLI: Tucker-Lewis index, RMSEA: root mean square error of approximation | | | | | | | | | |

### 4 Overview of all results of the structural equation model with “COVID-19 related life concerns”

| **Suppl. Table 4. Overview of all results of the structural equation model with “COVID-19 related life concerns” as exogeneous latent variable for mental health endogenous variables mediated by harmful and healthy behaviors**. The results are separated by endogenous variable (schizotypal traits, depressive symptoms, anxiety symptoms), mediators (excessive media use, drug consumption, alcohol consumption, units of exercise per week, hours of sleep per night during weeks) and samples at the four timepoints, showing indirect, direct and total effects, as well as individual pathways. | | | | | | | | | | | | | | | | |
| --- | --- | --- | --- | --- | --- | --- | --- | --- | --- | --- | --- | --- | --- | --- | --- | --- |
| Outcome | Mediators | Sample timepoint | indirect effect | |  | total effect | |  | a | |  | b | |  | c | |
|  |  |  | estimate | p |  | estimate | p |  | estimate | p |  | estimate | p |  | estimate | p |
| SPQ | Excessive Media use | 1 | 0.01 | .181 |  | 0.01 | .915 |  | 0.05 | .078 |  | 0.23 | .020 |  | 0.00 | .950 |
|  |  | 2 | 0.00 | .201 |  | 0.23 | .001 |  | 0.01 | .828 |  | 0.26 | .011 |  | 0.23 | .001 |
|  |  | 3 | 0.03 | .015 |  | 0.12 | .040 |  | 0.10 | .000 |  | 0.31 | .001 |  | 0.09 | .131 |
|  |  | 4 | 0.00 | .839 |  | 0.20 | .001 |  | 0.04 | .196 |  | 0.02 | .799 |  | 0.20 | .001 |
|  | Drug consumption | 1 | 0.00 | .561 |  | 0.00 | .997 |  | 0.03 | .266 |  | 0.15 | .318 |  | 0.00 | .950 |
|  |  | 2 | 0.01 | .427 |  | 0.23 | .001 |  | 0.04 | .009 |  | 0.14 | .295 |  | 0.23 | .001 |
|  |  | 3 | 0.10 | .430 |  | 0.10 | .077 |  | 0.09 | .000 |  | 0.11 | .400 |  | 0.09 | .131 |
|  |  | 4 | 0.01 | .334 |  | 0.21 | .000 |  | 0.06 | .018 |  | 0.15 | .277 |  | 0.20 | .001 |
|  | Alcohol consumption | 1 | 0.00 | .761 |  | 0.00 | .974 |  | 0.20 | .271 |  | 0.01 | .625 |  | 0.00 | .950 |
|  |  | 2 | 0.00 | .872 |  | 0.22 | .001 |  | -0.04 | .764 |  | 0.01 | .480 |  | 0.23 | .001 |
|  |  | 3 | 0.00 | .701 |  | 0.09 | .137 |  | 0.08 | .535 |  | -0.02 | .320 |  | 0.09 | .131 |
|  |  | 4 | -0.01 | .226 |  | 0.19 | .001 |  | 0.33 | .017 |  | -0.03 | .136 |  | 0.20 | .001 |
|  | Exercise (units per week) | 1 | 0.00 | .506 |  | 0.00 | .989 |  | -0.08 | .340 |  | -0.04 | .218 |  | 0.00 | .950 |
|  |  | 2 | 0.00 | .589 |  | 0.22 | .001 |  | -0.10 | .133 |  | 0.03 | .479 |  | 0.23 | .001 |
|  |  | 3 | 0.00 | .871 |  | 0.09 | .133 |  | -0.11 | .059 |  | 0.01 | .852 |  | 0.09 | .131 |
|  |  | 4 | 0.00 | .531 |  | 0.20 | .001 |  | -0.10 | .114 |  | 0.04 | .421 |  | 0.20 | .001 |
|  | Sleep (hours per night | 1 | 0.01 | .307 |  | 0.01 | .934 |  | -0.08 | .110 |  | -0.12 | .073 |  | 0.00 | .950 |
|  |  | 2 | 0.01 | .369 |  | 0.23 | .000 |  | -0.02 | .332 |  | -0.30 | .015 |  | 0.23 | .001 |
|  |  | 3 | 0.02 | .081 |  | 0.11 | .042 |  | -0.17 | .000 |  | -0.15 | .057 |  | 0.09 | .131 |
|  |  | 4 | 0.00 | .909 |  | 0.20 | .000 |  | -0.09 | .002 |  | -0.02 | .906 |  | 0.20 | .001 |
| Anxiety | Excessive Media use | 1 | 0.01 | .240 |  | 0.07 | .131 |  | 0.05 | .078 |  | 0.10 | .094 |  | 0.07 | .160 |
|  |  | 2 | 0.00 | .844 |  | 0.18 | .000 |  | 0.01 | .828 |  | 0.11 | .052 |  | 0.18 | .000 |
|  |  | 3 | 0.02 | .008 |  | 0.16 | .000 |  | 0.10 | .000 |  | 0.17 | .001 |  | 0.14 | .000 |
|  |  | 4 | 0.00 | .266 |  | 0.20 | .000 |  | 0.04 | .196 |  | 0.12 | .021 |  | 0.20 | .000 |
|  | Drug consumption | 1 | 0.00 | .726 |  | 0.07 | .147 |  | 0.03 | .266 |  | 0.05 | .591 |  | 0.07 | .160 |
|  |  | 2 | 0.00 | .384 |  | 0.19 | .000 |  | 0.04 | .009 |  | 0.11 | .218 |  | 0.18 | .000 |
|  |  | 3 | 0.01 | .121 |  | 0.16 | .000 |  | 0.09 | .000 |  | 0.14 | .083 |  | 0.14 | .000 |
|  |  | 4 | 0.00 | .951 |  | 0.20 | .000 |  | 0.06 | .018 |  | -0.01 | .965 |  | 0.20 | .000 |
|  | Alcohol consumption | 1 | 0.00 | .726 |  | 0.07 | .147 |  | 0.20 | .271 |  | 0.01 | .343 |  | 0.07 | .160 |
|  |  | 2 | 0.00 | .961 |  | 0.18 | .000 |  | -0.04 | .764 |  | 0.00 | .857 |  | 0.18 | .000 |
|  |  | 3 | 0.00 | .736 |  | 0.14 | .000 |  | 0.08 | .535 |  | -0.01 | .443 |  | 0.14 | .000 |
|  |  | 4 | 0.00 | .818 |  | 0.20 | .000 |  | 0.33 | .017 |  | 0.00 | .802 |  | 0.20 | .000 |
|  | Exercise (units per week) | 1 | 0.00 | .696 |  | 0.07 | .167 |  | -0.08 | .340 |  | 0.01 | .607 |  | 0.07 | .160 |
|  |  | 2 | 0.00 | .540 |  | 0.18 | .000 |  | -0.10 | .133 |  | 0.02 | .018 |  | 0.18 | .000 |
|  |  | 3 | 0.00 | .675 |  | 0.14 | .000 |  | -0.11 | .059 |  | 0.01 | .615 |  | 0.14 | .000 |
|  |  | 4 | -0.01 | .240 |  | 0.19 | .000 |  | -0.10 | .114 |  | 0.06 | .015 |  | 0.20 | .000 |
|  | Sleep (hours per night | 1 | 0.01 | .196 |  | 0.08 | .128 |  | -0.08 | .110 |  | -0.09 | .043 |  | 0.07 | .160 |
|  |  | 2 | 0.00 | .425 |  | 0.19 | .000 |  | -0.02 | .332 |  | -0.13 | .073 |  | 0.18 | .000 |
|  |  | 3 | 0.00 | .916 |  | 0.15 | .000 |  | -0.17 | .000 |  | -0.01 | .915 |  | 0.14 | .000 |
|  |  | 4 | 0.00 | .805 |  | 0.20 | .000 |  | -0.09 | .002 |  | -0.02 | .794 |  | 0.20 | .000 |
| Depression | Excessive Media use | 1 | 0.00 | .282 |  | 0.22 | .003 |  | 0.05 | .078 |  | 0.08 | .179 |  | 0.22 | .004 |
|  |  | 2 | 0.00 | .851 |  | 0.28 | .000 |  | 0.01 | .828 |  | 0.10 | .128 |  | 0.28 | .000 |
|  |  | 3 | 0.02 | .010 |  | 0.32 | .000 |  | 0.10 | .000 |  | 0.19 | .002 |  | 0.31 | .000 |
|  |  | 4 | 0.00 | .342 |  | 0.36 | .000 |  | 0.04 | .196 |  | 0.11 | .139 |  | 0.35 | .000 |
|  | Drug consumption | 1 | 0.00 | .603 |  | 0.22 | .004 |  | 0.03 | .266 |  | 0.08 | .439 |  | 0.22 | .004 |
|  |  | 2 | 0.01 | .381 |  | 0.28 | .000 |  | 0.04 | .009 |  | 0.14 | .223 |  | 0.28 | .000 |
|  |  | 3 | 0.01 | .176 |  | 0.32 | .000 |  | 0.09 | .000 |  | 0.14 | .166 |  | 0.31 | .000 |
|  |  | 4 | 0.01 | .323 |  | 0.36 | .000 |  | 0.06 | .018 |  | 0.12 | .265 |  | 0.35 | .000 |
|  | Alcohol consumption | 1 | 0.01 | .211 |  | 0.23 | .004 |  | 0.20 | .271 |  | 0.04 | .010 |  | 0.22 | .004 |
|  |  | 2 | 0.00 | .808 |  | 0.28 | .000 |  | -0.04 | .764 |  | 0.02 | .137 |  | 0.28 | .000 |
|  |  | 3 | 0.00 | .596 |  | 0.31 | .000 |  | 0.08 | .535 |  | 0.02 | .098 |  | 0.31 | .000 |
|  |  | 4 | 0.00 | .735 |  | 0.35 | .000 |  | 0.33 | .017 |  | 0.01 | .718 |  | 0.35 | .000 |
|  | Exercise (units per week) | 1 | 0.00 | .798 |  | 0.22 | .004 |  | -0.08 | .340 |  | -0.01 | .731 |  | 0.22 | .004 |
|  |  | 2 | 0.00 | .613 |  | 0.28 | .000 |  | -0.10 | .133 |  | -0.02 | .544 |  | 0.28 | .000 |
|  |  | 3 | 0.00 | .278 |  | 0.31 | .000 |  | -0.11 | .059 |  | -0.03 | .294 |  | 0.31 | .000 |
|  |  | 4 | 0.00 | .866 |  | 0.35 | .000 |  | -0.10 | .114 |  | 0.01 | .832 |  | 0.35 | .000 |
|  | Sleep (hours per night | 1 | 0.01 | .085 |  | 0.23 | .003 |  | -0.08 | .110 |  | -0.15 | .005 |  | 0.22 | .004 |
|  |  | 2 | 0.00 | .392 |  | 0.28 | .000 |  | -0.02 | .332 |  | -0.19 | .023 |  | 0.28 | .000 |
|  |  | 3 | 0.01 | .371 |  | 0.31 | .000 |  | -0.17 | .000 |  | -0.05 | .350 |  | 0.31 | .000 |
|  |  | 4 | 0.01 | .303 |  | 0.36 | .000 |  | -0.09 | .002 |  | -0.10 | .278 |  | 0.35 | .000 |
| a: pathway estimate between exogeneous latent variable and mediator, b: pathway estimate between mediator and endogenous variable, c: pathway estimate between exogeneous latent variable and endogenous variable | | | | | | | | | | | | | | | | |

### 5 Overview of all results of the structural equation model with “social adversity”

| **Suppl. Table 5. Overview of all results of the structural equation model with “social adversity” as a exogeneous latent variable for mental health endogenous latent variables with a mediation by harmful and healthy behaviors**. The results are separated by endogenous latent variable (schizotypal traits, depressive symptoms, anxiety symptoms), mediators (excessive media use, drug consumption, alcohol consumption, units of exercise per week, hours of sleep per night during weeks) and samples at the four timepoints, showing indirect, direct and total effects, as well as individual pathways. | | | | | | | | | | | | | | | | |
| --- | --- | --- | --- | --- | --- | --- | --- | --- | --- | --- | --- | --- | --- | --- | --- | --- |
| Outcome | Mediators | Sample timepoint | indirect effect | |  | total effect | |  | a | |  | b | |  | c | |
|  |  |  | estimate | p |  | estimate | p |  | estimate | p |  | estimate | p |  | estimate | p |
| SPQ | Excessive Media use | 1 | 0.01 | .156 |  | 0.13 | .027 |  | 0.05 | .062 |  | 0.21 | .022 |  | 0.12 | .043 |
|  |  | 2 | 0.01 | .219 |  | 0.25 | .000 |  | 0.04 | .122 |  | 0.25 | .015 |  | 0.24 | .001 |
|  |  | 3 | 0.03 | .017 |  | 0.26 | .000 |  | 0.11 | .000 |  | 0.25 | .004 |  | 0.23 | .000 |
|  |  | 4 | 0.00 | .989 |  | 0.33 | .000 |  | 0.05 | .056 |  | 0.00 | .987 |  | 0.33 | .000 |
|  | Drug consumption | 1 | 0.01 | .383 |  | 0.13 | .029 |  | 0.07 | .001 |  | 0.13 | .352 |  | 0.12 | .043 |
|  |  | 2 | 0.01 | .313 |  | 0.25 | .000 |  | 0.05 | .077 |  | 0.18 | .152 |  | 0.24 | .001 |
|  |  | 3 | 0.01 | .700 |  | 0.24 | .000 |  | 0.10 | .000 |  | 0.05 | .699 |  | 0.23 | .000 |
|  |  | 4 | 0.01 | .455 |  | 0.33 | .000 |  | 0.06 | .017 |  | 0.11 | .403 |  | 0.33 | .000 |
|  | Alcohol consumption | 1 | 0.00 | .903 |  | 0.12 | .038 |  | 0.50 | .000 |  | 0.13 | .352 |  | 0.12 | .043 |
|  |  | 2 | 0.00 | .805 |  | 0.24 | .001 |  | -0.08 | .570 |  | 0.01 | .577 |  | 0.24 | .001 |
|  |  | 3 | 0.00 | .477 |  | 0.23 | .000 |  | 0.16 | .209 |  | -0.02 | .288 |  | 0.23 | .000 |
|  |  | 4 | 0.00 | .810 |  | 0.33 | .000 |  | 0.05 | .693 |  | -0.02 | .385 |  | 0.33 | .000 |
|  | Exercise (units per week) | 1 | 0.00 | .785 |  | 0.12 | .047 |  | 0.03 | .715 |  | -0.04 | .202 |  | 0.12 | .043 |
|  |  | 2 | 0.00 | .971 |  | 0.24 | .001 |  | -0.06 | .454 |  | 0.00 | .951 |  | 0.24 | .001 |
|  |  | 3 | 0.00 | .968 |  | 0.23 | .000 |  | -0.03 | .587 |  | 0.00 | .934 |  | 0.23 | .000 |
|  |  | 4 | 0.00 | .606 |  | 0.33 | .000 |  | -0.05 | .434 |  | 0.05 | .279 |  | 0.33 | .000 |
|  | Sleep (hours per night | 1 | 0.01 | .257 |  | 0.13 | .020 |  | -0.16 | .000 |  | -0.09 | .217 |  | 0.12 | .043 |
|  |  | 2 | 0.02 | .131 |  | 0.26 | .000 |  | -0.11 | .000 |  | -0.23 | .115 |  | 0.24 | .001 |
|  |  | 3 | 0.01 | .223 |  | 0.25 | .000 |  | -0.13 | .001 |  | -0.10 | .196 |  | 0.23 | .000 |
|  |  | 4 | -0.01 | .736 |  | 0.32 | .000 |  | -0.10 | .000 |  | 0.05 | .717 |  | 0.33 | .000 |
| Anxiety | Excessive Media use | 1 | 0.00 | .340 |  | 0.26 | .000 |  | 0.05 | .062 |  | 0.06 | .226 |  | 0.25 | .000 |
|  |  | 2 | 0.00 | .286 |  | 0.27 | .000 |  | 0.04 | .122 |  | 0.09 | .116 |  | 0.27 | .000 |
|  |  | 3 | 0.01 | .021 |  | 0.28 | .000 |  | 0.11 | .000 |  | 0.13 | .012 |  | 0.26 | .000 |
|  |  | 4 | 0.00 | .192 |  | 0.29 | .000 |  | 0.05 | .056 |  | 0.09 | .058 |  | 0.29 | .000 |
|  | Drug consumption | 1 | 0.00 | .918 |  | 0.25 | .000 |  | 0.07 | .001 |  | -0.01 | .915 |  | 0.25 | .000 |
|  |  | 2 | 0.01 | .404 |  | 0.27 | .000 |  | 0.05 | .077 |  | 0.09 | .247 |  | 0.27 | .000 |
|  |  | 3 | 0.01 | .307 |  | 0.27 | .000 |  | 0.10 | .000 |  | 0.08 | .307 |  | 0.26 | .000 |
|  |  | 4 | 0.00 | .795 |  | 0.29 | .000 |  | 0.06 | .017 |  | -0,02 | .777 |  | 0.29 | .000 |
|  | Alcohol consumption | 1 | 0.00 | .513 |  | 0.25 | .000 |  | 0.50 | .000 |  | -0.01 | .487 |  | 0.25 | .000 |
|  |  | 2 | 0.00 | .936 |  | 0.27 | .000 |  | -0.08 | .570 |  | 0.00 | .861 |  | 0.27 | .000 |
|  |  | 3 | 0.00 | .526 |  | 0.26 | .000 |  | 0.16 | .209 |  | -0.01 | .324 |  | 0.26 | .000 |
|  |  | 4 | 0.00 | .842 |  | 0.29 | .000 |  | 0.05 | .693 |  | 0.01 | .511 |  | 0.29 | .000 |
|  | Exercise (units per week) | 1 | 0.00 | .982 |  | 0.25 | .000 |  | 0.03 | .715 |  | 0.00 | .951 |  | 0.25 | .000 |
|  |  | 2 | 0.00 | .872 |  | 0.27 | .000 |  | -0.06 | .454 |  | -0.01 | .784 |  | 0.27 | .000 |
|  |  | 3 | 0.00 | .988 |  | 0.26 | .000 |  | -0.03 | .587 |  | 0.00 | .976 |  | 0.26 | .000 |
|  |  | 4 | 0.00 | .485 |  | 0.28 | .000 |  | -0.05 | .434 |  | 0.05 | .011 |  | 0.29 | .000 |
|  | Sleep (hours per night | 1 | 0.00 | .539 |  | 0.26 | .000 |  | -0.16 | .000 |  | -0.02 | .524 |  | 0.25 | .000 |
|  |  | 2 | 0.00 | .847 |  | 0.27 | .000 |  | -0.11 | .000 |  | -0.01 | .843 |  | 0.27 | .000 |
|  |  | 3 | 0.00 | .627 |  | 0.26 | .000 |  | -0.13 | .001 |  | 0.02 | .620 |  | 0.26 | .000 |
|  |  | 4 | 0.00 | .623 |  | 0.28 | .000 |  | -0.10 | .000 |  | 0.03 | .597 |  | 0.29 | .000 |
| Depression | Excessive Media use | 1 | 0.00 | .489 |  | 0.48 | .000 |  | 0.05 | .062 |  | 0.04 | .430 |  | 0.47 | .000 |
|  |  | 2 | 0.00 | .464 |  | 0.47 | .000 |  | 0.04 | .122 |  | 0.06 | .353 |  | 0.46 | .000 |
|  |  | 3 | 0.01 | .024 |  | 0.51 | .000 |  | 0.11 | .000 |  | 0.13 | .019 |  | 0.49 | .000 |
|  |  | 4 | 0.00 | .393 |  | 0.56 | .000 |  | 0.05 | .056 |  | 0.06 | .333 |  | 0.55 | .000 |
|  | Drug consumption | 1 | 0.00 | .730 |  | 0.47 | .000 |  | 0.07 | .001 |  | -0.03 | .708 |  | 0.47 | .000 |
|  |  | 2 | 0.01 | .383 |  | 0.47 | .000 |  | 0.05 | .077 |  | -0.10 | .279 |  | 0.46 | .000 |
|  |  | 3 | 0.00 | .662 |  | 0.50 | .000 |  | 0.10 | .000 |  | 0.04 | .664 |  | 0.49 | .000 |
|  |  | 4 | 0.01 | .362 |  | 0.56 | .000 |  | 0.06 | .017 |  | 0.09 | .287 |  | 0.55 | .000 |
|  | Alcohol consumption | 1 | 0.00 | .667 |  | 0.48 | .000 |  | 0.50 | .000 |  | 0.01 | .663 |  | 0.47 | .000 |
|  |  | 2 | 0.00 | .670 |  | 0.46 | .000 |  | -0.08 | .570 |  | 0.02 | .102 |  | 0.46 | .000 |
|  |  | 3 | 0.00 | .373 |  | 0.50 | .000 |  | 0.16 | .209 |  | 0.02 | .202 |  | 0.49 | .000 |
|  |  | 4 | 0.00 | .747 |  | 0.55 | .000 |  | 0.05 | .693 |  | -0.02 | .149 |  | 0.55 | .000 |
|  | Exercise (units per week) | 1 | 0.00 | .761 |  | 0.47 | .000 |  | 0.03 | .715 |  | -0.03 | .113 |  | 0.47 | .000 |
|  |  | 2 | 0.00 | .571 |  | 0.47 | .000 |  | -0.06 | .454 |  | -0.03 | .212 |  | 0.46 | .000 |
|  |  | 3 | 0.00 | .621 |  | 0.50 | .000 |  | -0.03 | .587 |  | -0.04 | .079 |  | 0.49 | .000 |
|  |  | 4 | 0.00 | .876 |  | 0.55 | .000 |  | -0.05 | .434 |  | -0.01 | .795 |  | 0.55 | .000 |
|  | Sleep  (hours per  Night) | 1 | 0.01 | .384 |  | 0.48 | .000 |  | -0.16 | .000 |  | -0.03 | .371 |  | 0.47 | .000 |
|  |  | 2 | -0.01 | .463 |  | 0.46 | .000 |  | -0.11 | .000 |  | 0.06 | .431 |  | 0.46 | .000 |
|  |  | 3 | 0.00 | .814 |  | 0.50 | .000 |  | -0.13 | .001 |  | -0.01 | .807 |  | 0.49 | .000 |
|  |  | 4 | 0.00 | .868 |  | 0.55 | .000 |  | -0.10 | .000 |  | 0.03 | .597 |  | 0.55 | .000 |
| a: pathway estimate between exogeneous latent variable and mediator, b: pathway estimate between mediator and endogenous variable, c: pathway estimate between exogeneous latent variable and endogenous variable | | | | | | | | | | | | | | | | |

### 6 Complete outcome of structural equation models

#### 6.1 COVID-19 related life concerns’ model – first to forth timepoint

| 6.1.1 First Timepoint |
| --- |
| Estimator ML |
| Optimization method NLMINB |
| Number of free parameters 108 |
| Number of observations 480 |
| Model Test User Model: |
| Test statistic 480.156 |
| Degrees of freedom 135 |
| P-value (Chi-square) 0.000 |
| Model Test Baseline Model: |
| Test statistic 2326.466 |
| Degrees of freedom 225 |
| P-value 0.000 |
| User Model versus Baseline Model: |
| Comparative Fit Index (CFI) 0.836 |
| Tucker-Lewis Index (TLI) 0.726 |
| Loglikelihood and Information Criteria: |
| Loglikelihood user model (H0) -10237.799 |
| Loglikelihood unrestricted model (H1) -9997.722 |
| Akaike (AIC) 20691.599 |
| Bayesian (BIC) 21142.368 |
| Sample-size adjusted Bayesian (BIC) 20799.587 |
| Root Mean Square Error of Approximation: |
| RMSEA 0.073 |
| 90 Percent confidence interval - lower 0.066 |
| 90 Percent confidence interval - upper 0.080 |
| P-value RMSEA <= 0.05 0.000 |
| Standardized Root Mean Square Residual: |
| SRMR 0.067 |
| Parameter Estimates: |
| Standard errors Bootstrap |
| Number of requested bootstrap draws 1000 |
| Number of successful bootstrap draws 1000 |
| Latent Variables: |
| Estimate Std.Err z-value P(>\|z\|) Std.lv Std.all |
| livCond =~ |
| Nmbr_rms_hs_fl 1.000 2.201 0.789 |
| Nmbr_ppl_pr_hs 0.232 0.023 10.299 0.000 0.510 0.642 |
| LivingArea 0.324 0.030 10.924 0.000 0.713 0.528 |
| Garden_yard2 0.132 0.014 9.264 0.000 0.290 0.630 |
| healthBeforeCo19 =~ |
| Rglr_trtmnt_PI 1.000 0.163 0.477 |
| MntlHlthS_BC19 5.523 0.981 5.628 0.000 0.898 0.815 |
| PhysclHlt_BC19 3.489 0.628 5.554 0.000 0.567 0.574 |
| Co19_stress =~ |
| C19_fnncl_mpct 0.593 0.124 4.798 0.000 0.593 0.498 |
| Cncrnd_LfStblt 1.005 0.199 5.052 0.000 1.005 0.786 |
| Rstrctns_strss 0.557 0.159 3.512 0.000 0.557 0.467 |
| Regressions: |
| Estimate Std.Err z-value P(>\|z\|) Std.lv Std.all |
| depression ~ |
| hlthBC19 2.306 0.348 6.626 0.000 0.375 0.533 |
| Cntry_rs -0.284 0.067 -4.210 0.000 -0.284 -0.182 |
| Age -0.057 0.030 -1.854 0.064 -0.057 -0.080 |
| Gender 0.061 0.060 1.015 0.310 0.061 0.038 |
| Hghst___ -0.031 0.022 -1.447 0.148 -0.031 -0.054 |
| livCond 0.009 0.018 0.483 0.629 0.019 0.027 |
| anxiety ~ |
| hlthBC19 1.632 0.294 5.553 0.000 0.265 0.458 |
| Cntry_rs -0.240 0.061 -3.969 0.000 -0.240 -0.187 |
| Age -0.085 0.024 -3.542 0.000 -0.085 -0.146 |
| Gender 0.032 0.049 0.656 0.512 0.032 0.025 |
| Hghst___ 0.006 0.019 0.290 0.772 0.006 0.011 |
| livCond 0.006 0.015 0.376 0.707 0.012 0.022 |
| SPQ_total ~ |
| hlthBC19 2.671 0.488 5.469 0.000 0.434 0.447 |
| Cntry_rs 0.028 0.112 0.247 0.805 0.028 0.013 |
| Age -0.087 0.048 -1.813 0.070 -0.087 -0.089 |
| Gender -0.141 0.088 -1.604 0.109 -0.141 -0.064 |
| Hghst___ -0.063 0.040 -1.586 0.113 -0.063 -0.078 |
| livCond -0.020 0.024 -0.857 0.392 -0.045 -0.046 |
| Mscore.g ~ |
| hlthBC19 0.105 0.181 0.581 0.561 0.017 0.037 |
| Cntry_rs -0.174 0.050 -3.492 0.000 -0.174 -0.169 |
| Age -0.070 0.021 -3.309 0.001 -0.070 -0.151 |
| Gender -0.009 0.047 -0.193 0.847 -0.009 -0.009 |
| Hghst___ -0.046 0.017 -2.642 0.008 -0.046 -0.119 |
| livCond -0.018 0.013 -1.397 0.162 -0.039 -0.084 |
| drug_score.g ~ |
| hlthBC19 0.556 0.147 3.779 0.000 0.090 0.272 |
| Cntry_rs 0.011 0.036 0.300 0.764 0.011 0.015 |
| Age 0.025 0.016 1.562 0.118 0.025 0.075 |
| Gender -0.007 0.035 -0.209 0.835 -0.007 -0.010 |
| Hghst___ -0.005 0.012 -0.371 0.711 -0.005 -0.016 |
| livCond -0.004 0.008 -0.436 0.663 -0.008 -0.024 |
| Alcohol_DuringCo19 ~ |
| hlthBC19 -1.439 0.880 -1.635 0.102 -0.234 -0.104 |
| Cntry_rs -0.504 0.232 -2.169 0.030 -0.504 -0.101 |
| Age 0.456 0.102 4.466 0.000 0.456 0.202 |
| Gender -0.490 0.228 -2.144 0.032 -0.490 -0.096 |
| Hghst___ 0.063 0.092 0.686 0.493 0.063 0.034 |
| livCond 0.051 0.059 0.860 0.390 0.112 0.050 |
| Exercise_DuringCo19 ~ |
| hlthBC19 -1.090 0.523 -2.084 0.037 -0.177 -0.142 |
| Cntry_rs -0.508 0.138 -3.667 0.000 -0.508 -0.184 |
| Age 0.174 0.060 2.895 0.004 0.174 0.139 |
| Gender -0.047 0.127 -0.369 0.712 -0.047 -0.017 |
| Hghst___ 0.036 0.045 0.812 0.417 0.036 0.035 |
| livCond -0.013 0.032 -0.411 0.681 -0.029 -0.023 |
| Sleep_week_DuringCo19 ~ |
| hlthBC19 -0.095 0.256 -0.370 0.711 -0.015 -0.024 |
| Cntry_rs -0.087 0.067 -1.301 0.193 -0.087 -0.062 |
| Age -0.075 0.029 -2.575 0.010 -0.075 -0.118 |
| Gender 0.102 0.067 1.526 0.127 0.102 0.071 |
| Hghst___ -0.044 0.024 -1.799 0.072 -0.044 -0.083 |
| livCond -0.019 0.017 -1.118 0.264 -0.042 -0.066 |
| healthBeforeCo19 ~ |
| Cntry_rs -0.060 0.021 -2.812 0.005 -0.370 -0.167 |
| Age -0.025 0.010 -2.589 0.010 -0.154 -0.153 |
| Gender 0.045 0.020 2.281 0.023 0.278 0.123 |
| Hghst___ -0.020 0.008 -2.608 0.009 -0.124 -0.149 |
| livCond -0.008 0.006 -1.424 0.154 -0.107 -0.107 |
| depression ~ |
| C19_strs (c1) 0.217 0.076 2.851 0.004 0.217 0.309 |
| anxiety ~ |
| C19_strs (c2) 0.069 0.049 1.405 0.160 0.069 0.119 |
| SPQ_total ~ |
| C19_strs (c3) -0.004 0.065 -0.063 0.950 -0.004 -0.004 |
| Mscore.g ~ |
| C19_strs (a1) 0.048 0.027 1.762 0.078 0.048 0.103 |
| drug_score.g ~ |
| C19_strs (a2) 0.030 0.027 1.112 0.266 0.030 0.089 |
| Alcohol_DuringCo19 ~ |
| C19_strs (a3) 0.199 0.180 1.102 0.271 0.199 0.088 |
| Exercise_DuringCo19 ~ |
| C19_strs (a4) -0.079 0.082 -0.955 0.340 -0.079 -0.063 |
| Sleep_week_DuringCo19 ~ |
| C19_strs (a5) -0.080 0.050 -1.600 0.110 -0.080 -0.125 |
| depression ~ |
| Mscore.g (b1) 0.084 0.061 1.372 0.170 0.084 0.056 |
| drg_scr. (b2) 0.076 0.098 0.775 0.439 0.076 0.036 |
| Alc_DC19 (b3) 0.036 0.014 2.588 0.010 0.036 0.114 |
| Exr_DC19 (b4) -0.008 0.022 -0.343 0.731 -0.008 -0.014 |
| Sl__DC19 (b5) -0.152 0.054 -2.797 0.005 -0.152 -0.137 |
| anxiety ~ |
| Mscore.g (b6) 0.095 0.057 1.674 0.094 0.095 0.076 |
| drg_scr. (b7) 0.052 0.097 0.538 0.591 0.052 0.030 |
| Alc_DC19 (b8) 0.011 0.011 0.948 0.343 0.011 0.041 |
| Exr_DC19 (b9) 0.011 0.021 0.514 0.607 0.011 0.023 |
| Sl__DC19 (b10) -0.094 0.046 -2.023 0.043 -0.094 -0.103 |
| SPQ_total ~ |
| Mscore.g (b11) 0.231 0.100 2.323 0.020 0.231 0.111 |
| drg_scr. (b12) 0.147 0.148 0.998 0.318 0.147 0.050 |
| Alc_DC19 (b13) 0.010 0.021 0.489 0.625 0.010 0.024 |
| Exr_DC19 (b14) -0.041 0.033 -1.233 0.218 -0.041 -0.053 |
| Sl__DC19 (b15) -0.118 0.066 -1.791 0.073 -0.118 -0.077 |
| Covariances: |
| Estimate Std.Err z-value P(>\|z\|) Std.lv Std.all |
| .anxiety ~~ |
| .SPQ_total 0.135 0.025 5.446 0.000 0.135 0.363 |
| .depression ~~ |
| .SPQ_total 0.058 0.026 2.222 0.026 0.058 0.151 |
| .anxiety 0.099 0.019 5.086 0.000 0.099 0.458 |
| livCond ~~ |
| Co19_stress -0.443 0.150 -2.958 0.003 -0.201 -0.201 |
| Variances: |
| Estimate Std.Err z-value P(>\|z\|) Std.lv Std.all |
| Co19_stress 1.000 1.000 1.000 |
| .Nmbr_rms_hs_fl 2.931 0.457 6.410 0.000 2.931 0.377 |
| .Nmbr_ppl_pr_hs 0.371 0.028 13.260 0.000 0.371 0.588 |
| .LivingArea 1.318 0.098 13.503 0.000 1.318 0.722 |
| .Garden_yard2 0.128 0.010 13.073 0.000 0.128 0.603 |
| .Rglr_trtmnt_PI 0.089 0.009 10.132 0.000 0.089 0.772 |
| .MntlHlthS_BC19 0.409 0.098 4.164 0.000 0.409 0.336 |
| .PhysclHlt_BC19 0.655 0.050 13.064 0.000 0.655 0.671 |
| .C19_fnncl_mpct 1.064 0.111 9.620 0.000 1.064 0.752 |
| .Cncrnd_LfStblt 0.624 0.387 1.613 0.107 0.624 0.381 |
| .Rstrctns_strss 1.114 0.195 5.717 0.000 1.114 0.782 |
| .depression 0.224 0.033 6.752 0.000 0.224 0.453 |
| .anxiety 0.207 0.022 9.239 0.000 0.207 0.618 |
| .SPQ_total 0.664 0.061 10.812 0.000 0.664 0.705 |
| .Mscore.g 0.193 0.009 21.552 0.000 0.193 0.896 |
| .drug_score.g 0.101 0.010 9.649 0.000 0.101 0.917 |
| .Alcohl_DrngC19 4.606 0.199 23.100 0.000 4.606 0.913 |
| .Exercs_DrngC19 1.438 0.076 18.908 0.000 1.438 0.929 |
| .Slp_wk_DrngC19 0.383 0.027 14.418 0.000 0.383 0.951 |
| livCond 4.846 0.569 8.518 0.000 1.000 1.000 |
| .healthBeforC19 0.023 0.006 3.620 0.000 0.886 0.886 |
| Defined Parameters: |
| Estimate Std.Err z-value P(>\|z\|) Std.lv Std.all |
| indirect1 0.004 0.004 1.075 0.282 0.004 0.006 |
| indirect2 0.002 0.004 0.520 0.603 0.002 0.003 |
| indirect3 0.007 0.006 1.250 0.211 0.007 0.010 |
| indirect4 0.001 0.002 0.256 0.798 0.001 0.001 |
| indirect5 0.012 0.007 1.721 0.085 0.012 0.017 |
| indirect6 0.005 0.004 1.174 0.240 0.005 0.008 |
| indirect7 0.002 0.004 0.351 0.726 0.002 0.003 |
| indirect8 0.002 0.003 0.620 0.535 0.002 0.004 |
| indirect9 -0.001 0.002 -0.390 0.696 -0.001 -0.001 |
| indirect10 0.007 0.006 1.293 0.196 0.007 0.013 |
| indirect11 0.011 0.008 1.337 0.181 0.011 0.011 |
| indirect12 0.004 0.008 0.581 0.561 0.004 0.005 |
| indirect13 0.002 0.007 0.304 0.761 0.002 0.002 |
| indirect14 0.003 0.005 0.666 0.506 0.003 0.003 |
| indirect15 0.009 0.009 1.021 0.307 0.009 0.010 |
| total1 0.221 0.075 2.934 0.003 0.221 0.315 |
| total2 0.220 0.076 2.897 0.004 0.220 0.312 |
| total3 0.225 0.077 2.899 0.004 0.225 0.319 |
| total4 0.218 0.076 2.868 0.004 0.218 0.310 |
| total5 0.229 0.078 2.933 0.003 0.229 0.326 |
| total6 0.073 0.049 1.509 0.131 0.073 0.127 |
| total7 0.070 0.049 1.449 0.147 0.070 0.122 |
| total8 0.071 0.049 1.449 0.147 0.071 0.123 |
| total9 0.068 0.049 1.383 0.167 0.068 0.117 |
| total10 0.076 0.050 1.523 0.128 0.076 0.132 |
| total11 0.007 0.066 0.107 0.915 0.007 0.007 |
| total12 0.000 0.064 0.004 0.997 0.000 0.000 |
| total13 -0.002 0.064 -0.033 0.974 -0.002 -0.002 |
| total14 -0.001 0.066 -0.014 0.989 -0.001 -0.001 |
| total15 0.005 0.063 0.083 0.934 0.005 0.005 |

| 6.1.2 Second Timepoint |
| --- |
| Estimator ML |
| Optimization method NLMINB |
| Number of free parameters 108 |
| Used Total |
| Number of observations 431 464 |
| Model Test User Model: |
| Test statistic 440.014 |
| Degrees of freedom 135 |
| P-value (Chi-square) 0.000 |
| Model Test Baseline Model: |
| Test statistic 2430.458 |
| Degrees of freedom 225 |
| P-value 0.000 |
| User Model versus Baseline Model: |
| Comparative Fit Index (CFI) 0.862 |
| Tucker-Lewis Index (TLI) 0.770 |
| Loglikelihood and Information Criteria: |
| Loglikelihood user model (H0) -8403.637 |
| Loglikelihood unrestricted model (H1) -8183.630 |
| Akaike (AIC) 17023.274 |
| Bayesian (BIC) 17462.414 |
| Sample-size adjusted Bayesian (BIC) 17119.684 |
| Root Mean Square Error of Approximation: |
| RMSEA 0.072 |
| 90 Percent confidence interval - lower 0.065 |
| 90 Percent confidence interval - upper 0.080 |
| P-value RMSEA <= 0.05 0.000 |
| Standardized Root Mean Square Residual: |
| SRMR 0.063 |
| Parameter Estimates: |
| Standard errors Bootstrap |
| Number of requested bootstrap draws 1000 |
| Number of successful bootstrap draws 1000 |
| Latent Variables: |
| Estimate Std.Err z-value P(>\|z\|) Std.lv Std.all |
| livCond =~ |
| Nmbr_rms___FU1 1.000 1.988 0.905 |
| Nmbr_ppl___FU1 0.412 0.035 11.667 0.000 0.819 0.647 |
| Living_Are_FU1 0.344 0.041 8.287 0.000 0.683 0.498 |
| Grdn_yrd_FU1_2 0.134 0.017 8.018 0.000 0.267 0.569 |
| healthBeforeCo19 =~ |
| Rglr_tr_PI_FU1 1.000 0.159 0.496 |
| MntHS_BC19_FU1 -5.172 1.110 -4.661 0.000 -0.824 -0.803 |
| PhysH_BC19_FU1 -3.316 0.679 -4.880 0.000 -0.528 -0.573 |
| Co19_stress =~ |
| C19_fnncl__FU1 0.602 0.065 9.220 0.000 0.602 0.628 |
| Cncrnd_LfS_FU1 0.912 0.078 11.727 0.000 0.912 0.813 |
| Rstrctns_s_FU1 0.453 0.065 6.973 0.000 0.453 0.437 |
| Regressions: |
| Estimate Std.Err z-value P(>\|z\|) Std.lv Std.all |
| depression ~ |
| hlthBC19 -1.724 0.358 -4.810 0.000 -0.275 -0.412 |
| Cnt__FU1 -0.277 0.068 -4.061 0.000 -0.277 -0.174 |
| age_FU1 -0.006 0.002 -3.793 0.000 -0.006 -0.144 |
| Gender -0.009 0.061 -0.146 0.884 -0.009 -0.006 |
| Hghst___ 0.012 0.021 0.553 0.580 0.012 0.022 |
| livCond -0.003 0.015 -0.216 0.829 -0.007 -0.010 |
| anxiety ~ |
| hlthBC19 -1.390 0.254 -5.476 0.000 -0.221 -0.406 |
| Cnt__FU1 -0.163 0.065 -2.496 0.013 -0.163 -0.125 |
| age_FU1 -0.008 0.001 -5.806 0.000 -0.008 -0.220 |
| Gender 0.019 0.048 0.384 0.701 0.019 0.015 |
| Hghst___ -0.040 0.020 -2.033 0.042 -0.040 -0.090 |
| livCond 0.012 0.014 0.829 0.407 0.023 0.043 |
| SPQ_total ~ |
| hlthBC19 -2.861 0.534 -5.353 0.000 -0.456 -0.466 |
| Cnt__FU1 -0.034 0.118 -0.286 0.775 -0.034 -0.014 |
| age_FU1 -0.006 0.002 -2.606 0.009 -0.006 -0.100 |
| Gender -0.213 0.093 -2.284 0.022 -0.213 -0.096 |
| Hghst___ -0.119 0.034 -3.482 0.000 -0.119 -0.150 |
| livCond 0.020 0.026 0.788 0.430 0.040 0.041 |
| Mscore.g ~ |
| hlthBC19 -0.272 0.161 -1.695 0.090 -0.043 -0.105 |
| Cnt__FU1 -0.122 0.047 -2.605 0.009 -0.122 -0.123 |
| age_FU1 -0.004 0.001 -2.911 0.004 -0.004 -0.154 |
| Gender -0.039 0.043 -0.912 0.362 -0.039 -0.042 |
| Hghst___ -0.080 0.015 -5.174 0.000 -0.080 -0.239 |
| livCond -0.007 0.011 -0.645 0.519 -0.014 -0.035 |
| drug_score.g ~ |
| hlthBC19 -0.256 0.152 -1.679 0.093 -0.041 -0.119 |
| Cnt__FU1 -0.103 0.047 -2.172 0.030 -0.103 -0.126 |
| age_FU1 -0.000 0.001 -0.366 0.714 -0.000 -0.016 |
| Gender 0.029 0.035 0.850 0.395 0.029 0.038 |
| Hghst___ -0.032 0.015 -2.045 0.041 -0.032 -0.114 |
| livCond -0.021 0.008 -2.465 0.014 -0.041 -0.120 |
| Alcohol_FU1 ~ |
| hlthBC19 1.471 0.915 1.608 0.108 0.234 0.109 |
| Cnt__FU1 -0.506 0.247 -2.046 0.041 -0.506 -0.099 |
| age_FU1 0.018 0.007 2.665 0.008 0.018 0.134 |
| Gender -0.462 0.236 -1.955 0.051 -0.462 -0.095 |
| Hghst___ 0.166 0.093 1.772 0.076 0.166 0.095 |
| livCond 0.042 0.058 0.720 0.471 0.083 0.039 |
| exerciseMin_FU1 ~ |
| hlthBC19 1.044 0.443 2.355 0.019 0.166 0.164 |
| Cnt__FU1 -0.186 0.126 -1.472 0.141 -0.186 -0.077 |
| age_FU1 0.000 0.003 0.042 0.967 0.000 0.002 |
| Gender -0.165 0.115 -1.438 0.150 -0.165 -0.072 |
| Hghst___ 0.055 0.042 1.308 0.191 0.055 0.067 |
| livCond -0.010 0.027 -0.369 0.712 -0.020 -0.019 |
| Sleep_week_FU1 ~ |
| hlthBC19 0.397 0.153 2.588 0.010 0.063 0.183 |
| Cnt__FU1 0.070 0.042 1.664 0.096 0.070 0.085 |
| age_FU1 -0.001 0.001 -1.356 0.175 -0.001 -0.061 |
| Gender 0.061 0.041 1.472 0.141 0.061 0.078 |
| Hghst___ 0.020 0.016 1.289 0.197 0.020 0.072 |
| livCond -0.003 0.010 -0.360 0.719 -0.007 -0.020 |
| healthBeforeCo19 ~ |
| Cnt__FU1 0.085 0.028 3.052 0.002 0.534 0.224 |
| age_FU1 0.001 0.001 1.598 0.110 0.006 0.089 |
| Gender 0.013 0.019 0.654 0.513 0.079 0.035 |
| Hghst___ 0.015 0.010 1.572 0.116 0.095 0.117 |
| livCond 0.017 0.007 2.573 0.010 0.217 0.217 |
| depression ~ |
| C19_strs (c1) 0.278 0.044 6.352 0.000 0.278 0.417 |
| anxiety ~ |
| C19_strs (c2) 0.183 0.047 3.899 0.000 0.183 0.336 |
| SPQ_total ~ |
| C19_strs (c3) 0.225 0.066 3.400 0.001 0.225 0.230 |
| Mscore.g ~ |
| C19_strs (a1) 0.005 0.024 0.217 0.828 0.005 0.012 |
| drug_score.g ~ |
| C19_strs (a2) 0.038 0.023 1.650 0.099 0.038 0.110 |
| Alcohol_FU1 ~ |
| C19_strs (a3) -0.041 0.137 -0.300 0.764 -0.041 -0.019 |
| exerciseMin_FU1 ~ |
| C19_strs (a4) -0.099 0.066 -1.503 0.133 -0.099 -0.097 |
| Sleep_week_FU1 ~ |
| C19_strs (a5) -0.022 0.023 -0.969 0.332 -0.022 -0.063 |
| depression ~ |
| Mscore.g (b1) 0.099 0.065 1.523 0.128 0.099 0.061 |
| drg_scr. (b2) 0.140 0.115 1.219 0.223 0.140 0.072 |
| Alch_FU1 (b3) 0.020 0.013 1.489 0.137 0.020 0.063 |
| exrM_FU1 (b4) -0.015 0.025 -0.606 0.544 -0.015 -0.023 |
| Slp__FU1 (b5) -0.189 0.083 -2.272 0.023 -0.189 -0.098 |
| anxiety ~ |
| Mscore.g (b6) 0.105 0.054 1.942 0.052 0.105 0.080 |
| drg_scr. (b7) 0.110 0.089 1.233 0.218 0.110 0.069 |
| Alch_FU1 (b8) 0.002 0.011 0.181 0.857 0.002 0.008 |
| exrM_FU1 (b9) 0.018 0.023 0.792 0.428 0.018 0.034 |
| Slp__FU1 (b10) -0.130 0.072 -1.792 0.073 -0.130 -0.082 |
| SPQ_total ~ |
| Mscore.g (b11) 0.255 0.101 2.532 0.011 0.255 0.108 |
| drg_scr. (b12) 0.138 0.132 1.048 0.295 0.138 0.048 |
| Alch_FU1 (b13) 0.013 0.019 0.706 0.480 0.013 0.030 |
| exrM_FU1 (b14) 0.031 0.044 0.708 0.479 0.031 0.032 |
| Slp__FU1 (b15) -0.300 0.124 -2.428 0.015 -0.300 -0.106 |
| Covariances: |
| Estimate Std.Err z-value P(>\|z\|) Std.lv Std.all |
| .anxiety ~~ |
| .SPQ_total 0.121 0.024 5.041 0.000 0.121 0.416 |
| .depression ~~ |
| .SPQ_total 0.076 0.027 2.791 0.005 0.076 0.225 |
| .anxiety 0.088 0.017 5.181 0.000 0.088 0.495 |
| livCond ~~ |
| Co19_stress -0.176 0.123 -1.422 0.155 -0.088 -0.088 |
| Variances: |
| Estimate Std.Err z-value P(>\|z\|) Std.lv Std.all |
| Co19_stress 1.000 1.000 1.000 |
| .Nmbr_rms___FU1 0.870 0.309 2.819 0.005 0.870 0.180 |
| .Nmbr_ppl___FU1 0.928 0.076 12.292 0.000 0.928 0.581 |
| .Living_Are_FU1 1.412 0.092 15.393 0.000 1.412 0.752 |
| .Grdn_yrd_FU1_2 0.149 0.010 14.352 0.000 0.149 0.677 |
| .Rglr_tr_PI_FU1 0.078 0.008 9.167 0.000 0.078 0.754 |
| .MntHS_BC19_FU1 0.374 0.092 4.068 0.000 0.374 0.356 |
| .PhysH_BC19_FU1 0.571 0.057 10.019 0.000 0.571 0.672 |
| .C19_fnncl__FU1 0.557 0.063 8.874 0.000 0.557 0.606 |
| .Cncrnd_LfS_FU1 0.426 0.109 3.915 0.000 0.426 0.339 |
| .Rstrctns_s_FU1 0.868 0.067 12.934 0.000 0.868 0.809 |
| .depression 0.206 0.028 7.301 0.000 0.206 0.464 |
| .anxiety 0.155 0.020 7.892 0.000 0.155 0.521 |
| .SPQ_total 0.550 0.047 11.603 0.000 0.550 0.576 |
| .Mscore.g 0.149 0.010 182 0.000 0.149 0.873 |
| .drug_score.g 0.106 0.011 9.588 0.000 0.106 0.910 |
| .Alcohol_FU1 4.300 0.169 25.434 0.000 4.300 0.936 |
| .exerciseMn_FU1 0.981 0.072 13.698 0.000 0.981 0.951 |
| .Sleep_week_FU1 0.112 0.011 10.198 0.000 0.112 0.935 |
| livCond 3.953 0.475 8.319 0.000 1.000 1.000 |
| .healthBeforC19 0.022 0.006 3.490 0.000 0.879 0.879 |
| Defined Parameters: |
| Estimate Std.Err z-value P(>\|z\|) Std.lv Std.all |
| indirect1 0.001 0.003 0.188 0.851 0.001 0.001 |
| indirect2 0.005 0.006 0.877 0.381 0.005 0.008 |
| indirect3 -0.001 0.003 -0.243 0.808 -0.001 -0.001 |
| indirect4 0.001 0.003 0.506 0.613 0.001 0.002 |
| indirect5 0.004 0.005 0.855 0.392 0.004 0.006 |
| indirect6 0.001 0.003 0.197 0.844 0.001 0.001 |
| indirect7 0.004 0.005 0.870 0.384 0.004 0.008 |
| indirect8 -0.000 0.002 -0.049 0.961 -0.000 -0.000 |
| indirect9 -0.002 0.003 -0.613 0.540 -0.002 -0.003 |
| indirect10 0.003 0.004 0.798 0.425 0.003 0.005 |
| indirect11 0.001 0.007 0.201 0.840 0.001 0.001 |
| indirect12 0.005 0.007 0.776 0.437 0.005 0.005 |
| indirect13 -0.001 0.003 -0.161 0.872 -0.001 -0.001 |
| indirect14 -0.003 0.006 -0.540 0.589 -0.003 -0.003 |
| indirect15 0.007 0.007 0.898 0.369 0.007 0.007 |
| total1 0.278 0.044 6.350 0.000 0.278 0.418 |
| total2 0.283 0.045 6.347 0.000 0.283 0.425 |
| total3 0.277 0.044 6.227 0.000 0.277 0.416 |
| total4 0.279 0.043 6.432 0.000 0.279 0.419 |
| total5 0.282 0.044 6.388 0.000 0.282 0.423 |
| total6 0.183 0.047 3.924 0.000 0.183 0.337 |
| total7 0.187 0.047 3.957 0.000 0.187 0.343 |
| total8 0.183 0.047 3.906 0.000 0.183 0.335 |
| total9 0.181 0.046 3.899 0.000 0.181 0.332 |
| total10 0.186 0.047 3.914 0.000 0.186 0.341 |
| total11 0.226 0.066 3.408 0.001 0.226 0.231 |
| total12 0.230 0.066 3.480 0.001 0.230 0.235 |
| total13 0.224 0.066 3.397 0.001 0.224 0.229 |
| total14 0.222 0.066 3.384 0.001 0.222 0.227 |
| total15 0.231 0.066 3.492 0.000 0.231 0.237 |

| 6.1.3 Third Timepoint |
| --- |
| Estimator ML |
| Optimization method NLMINB |
| Number of free parameters 108 |
| Used Total |
| Number of observations 496 532 |
| Model Test User Model: |
| Test statistic 426.425 |
| Degrees of freedom 135 |
| P-value (Chi-square) 0.000 |
| Model Test Baseline Model: |
| Test statistic 2725.462 |
| Degrees of freedom 225 |
| P-value 0.000 |
| User Model versus Baseline Model: |
| Comparative Fit Index (CFI) 0.883 |
| Tucker-Lewis Index (TLI) 0.806 |
| Loglikelihood and Information Criteria: |
| Loglikelihood user model (H0) -10554.934 |
| Loglikelihood unrestricted model (H1) -10341.722 |
| Akaike (AIC) 21325.868 |
| Bayesian (BIC) 21780.178 |
| Sample-size adjusted Bayesian (BIC) 21437.383 |
| Root Mean Square Error of Approximation: |
| RMSEA 0.066 |
| 90 Percent confidence interval - lower 0.059 |
| 90 Percent confidence interval - upper 0.073 |
| P-value RMSEA <= 0.05 0.000 |
| Standardized Root Mean Square Residual: |
| SRMR 0.056 |
| Parameter Estimates: |
| Standard errors Bootstrap |
| Number of requested bootstrap draws 1000 |
| Number of successful bootstrap draws 1000 |
| Latent Variables: |
| Estimate Std.Err z-value P(>\|z\|) Std.lv Std.all |
| livCond =~ |
| Nmbr_rms___FU2 1.000 1.744 0.804 |
| Nmbr_ppl___FU2 0.402 0.041 9.927 0.000 0.702 0.531 |
| Living_Are_FU2 0.424 0.062 6.815 0.000 0.739 0.535 |
| Grdn_yrd_FU2_2 0.159 0.023 6.783 0.000 0.278 0.605 |
| healthBeforeCo19 =~ |
| Rglr_tr_PI_FU2 1.000 0.205 0.566 |
| MntHS_BC19_FU2 -4.338 0.614 -7.069 0.000 -0.889 -0.867 |
| PhysH_BC19_FU2 -2.995 0.416 -7.207 0.000 -0.614 -0.619 |
| Co19_stress =~ |
| C19_fnncl__FU2 0.672 0.066 10.154 0.000 0.672 0.627 |
| Cncrnd_LIS_FU2 0.997 0.062 16.127 0.000 0.997 0.772 |
| Rstrctns_s_FU2 0.780 0.064 12.109 0.000 0.780 0.660 |
| Regressions: |
| Estimate Std.Err z-value P(>\|z\|) Std.lv Std.all |
| depression ~ |
| hlthBC19 -1.575 0.228 -6.893 0.000 -0.323 -0.434 |
| Cnt__FU2 -0.189 0.069 -2.755 0.006 -0.189 -0.117 |
| age_FU2 -0.005 0.002 -2.952 0.003 -0.005 -0.107 |
| Gender 0.027 0.062 0.434 0.664 0.027 0.016 |
| Hghst___ 0.020 0.019 1.038 0.299 0.020 0.039 |
| livCond 0.019 0.020 0.973 0.331 0.034 0.045 |
| anxiety ~ |
| hlthBC19 -1.120 0.206 -5.447 0.000 -0.229 -0.400 |
| Cnt__FU2 -0.140 0.063 -2.226 0.026 -0.140 -0.112 |
| age_FU2 -0.007 0.001 -4.627 0.000 -0.007 -0.172 |
| Gender 0.125 0.043 2.911 0.004 0.125 0.098 |
| Hghst___ 0.011 0.017 0.678 0.498 0.011 0.030 |
| livCond 0.021 0.016 1.317 0.188 0.037 0.065 |
| SPQ_total ~ |
| hlthBC19 -1.836 0.300 -6.112 0.000 -0.376 -0.373 |
| Cnt__FU2 -0.083 0.109 -0.761 0.446 -0.083 -0.038 |
| age_FU2 -0.011 0.003 -4.336 0.000 -0.011 -0.167 |
| Gender 0.039 0.095 0.411 0.681 0.039 0.017 |
| Hghst___ -0.065 0.028 -2.286 0.022 -0.065 -0.096 |
| livCond -0.023 0.029 -0.787 0.431 -0.039 -0.039 |
| Mscore.g ~ |
| hlthBC19 -0.130 0.123 -1.055 0.292 -0.027 -0.055 |
| Cnt__FU2 -0.059 0.044 -1.331 0.183 -0.059 -0.056 |
| age_FU2 -0.001 0.001 -0.836 0.403 -0.001 -0.037 |
| Gender -0.140 0.048 -2.907 0.004 -0.140 -0.130 |
| Hghst___ -0.071 0.014 -5.258 0.000 -0.071 -0.218 |
| livCond -0.031 0.015 -2.033 0.042 -0.054 -0.110 |
| drug_score.g ~ |
| hlthBC19 -0.387 0.101 -3.818 0.000 -0.079 -0.214 |
| Cnt__FU2 0.054 0.038 1.393 0.164 0.054 0.067 |
| age_FU2 -0.001 0.001 -1.364 0.173 -0.001 -0.058 |
| Gender 0.008 0.034 0.247 0.805 0.008 0.010 |
| Hghst___ -0.004 0.011 -0.346 0.729 -0.004 -0.016 |
| livCond -0.014 0.011 -1.258 0.208 -0.024 -0.065 |
| Alcohol_FU2 ~ |
| hlthBC19 1.292 0.567 2.279 0.023 0.265 0.119 |
| Cnt__FU2 -0.233 0.209 -1.114 0.265 -0.233 -0.048 |
| age_FU2 0.022 0.007 3.212 0.001 0.022 0.147 |
| Gender -0.237 0.232 -1.022 0.307 -0.237 -0.048 |
| Hghst___ 0.233 0.070 3.342 0.001 0.233 0.156 |
| livCond 0.012 0.063 0.183 0.855 0.020 0.009 |
| exerciseMin_FU2 ~ |
| hlthBC19 -0.130 0.291 -0.448 0.654 -0.027 -0.023 |
| Cnt__FU2 -0.201 0.116 -1.737 0.082 -0.201 -0.080 |
| age_FU2 0.004 0.004 1.056 0.291 0.004 0.050 |
| Gender 0.110 0.113 0.971 0.331 0.110 0.043 |
| Hghst___ 0.142 0.035 4.023 0.000 0.142 0.183 |
| livCond 0.070 0.036 1.914 0.056 0.121 0.105 |
| Sleep_week_FU2 ~ |
| hlthBC19 -0.065 0.189 -0.341 0.733 -0.013 -0.021 |
| Cnt__FU2 0.143 0.071 2.026 0.043 0.143 0.104 |
| age_FU2 -0.004 0.002 -2.324 0.020 -0.004 -0.105 |
| Gender 0.264 0.058 4.532 0.000 0.264 0.188 |
| Hghst___ 0.008 0.018 0.468 0.640 0.008 0.019 |
| livCond -0.016 0.020 -0.832 0.405 -0.028 -0.045 |
| healthBeforeCo19 ~ |
| Cnt__FU2 0.050 0.026 1.929 0.054 0.245 0.113 |
| age_FU2 0.001 0.001 2.203 0.028 0.007 0.107 |
| Gender -0.022 0.022 -1.016 0.310 -0.109 -0.049 |
| Hghst___ 0.002 0.007 0.235 0.814 0.008 0.012 |
| livCond 0.011 0.009 1.260 0.208 0.092 0.092 |
| depression ~ |
| C19_strs (c1) 0.305 0.048 6.387 0.000 0.305 0.409 |
| anxiety ~ |
| C19_strs (c2) 0.144 0.036 3.987 0.000 0.144 0.252 |
| SPQ_total ~ |
| C19_strs (c3) 0.090 0.060 1.510 0.131 0.090 0.089 |
| Mscore.g ~ |
| C19_strs (a1) 0.099 0.026 3.850 0.000 0.099 0.203 |
| drug_score.g ~ |
| C19_strs (a2) 0.091 0.025 3.693 0.000 0.091 0.246 |
| Alcohol_FU2 ~ |
| C19_strs (a3) 0.075 0.120 0.621 0.535 0.075 0.034 |
| exerciseMin_FU2 ~ |
| C19_strs (a4) -0.107 0.057 -1.887 0.059 -0.107 -0.093 |
| Sleep_week_FU2 ~ |
| C19_strs (a5) -0.168 0.033 -5.099 0.000 -0.168 -0.264 |
| depression ~ |
| Mscore.g (b1) 0.189 0.062 3.054 0.002 0.189 0.124 |
| drg_scr. (b2) 0.140 0.101 1.386 0.166 0.140 0.070 |
| Alch_FU2 (b3) 0.020 0.012 1.653 0.098 0.020 0.061 |
| exrM_FU2 (b4) -0.025 0.024 -1.049 0.294 -0.025 -0.039 |
| Slp__FU2 (b5) -0.046 0.050 -0.934 0.350 -0.046 -0.040 |
| anxiety ~ |
| Mscore.g (b6) 0.174 0.051 3.406 0.001 0.174 0.148 |
| drg_scr. (b7) 0.143 0.082 1.731 0.083 0.143 0.092 |
| Alch_FU2 (b8) -0.009 0.011 -0.767 0.443 -0.009 -0.034 |
| exrM_FU2 (b9) 0.009 0.019 0.503 0.615 0.009 0.019 |
| Slp__FU2 (b10) -0.005 0.049 -0.107 0.915 -0.005 -0.006 |
| SPQ_total ~ |
| Mscore.g (b11) 0.306 0.089 3.444 0.001 0.306 0.148 |
| drg_scr. (b12) 0.114 0.136 0.842 0.400 0.114 0.042 |
| Alch_FU2 (b13) -0.020 0.020 -0.995 0.320 -0.020 -0.043 |
| exrM_FU2 (b14) 0.007 0.035 0.187 0.852 0.007 0.007 |
| Slp__FU2 (b15) -0.145 0.076 -1.906 0.057 -0.145 -0.091 |
| Covariances: |
| Estimate Std.Err z-value P(>\|z\|) Std.lv Std.all |
| .anxiety ~~ |
| .SPQ_total 0.209 0.028 7.382 0.000 0.209 0.550 |
| .depression ~~ |
| .SPQ_total 0.178 0.029 6.099 0.000 0.178 0.394 |
| .anxiety 0.131 0.020 6.710 0.000 0.131 0.553 |
| livCond ~~ |
| Co19_stress -0.094 0.116 -0.811 0.418 -0.054 -0.054 |
| Variances: |
| Estimate Std.Err z-value P(>\|z\|) Std.lv Std.all |
| Co19_stress 1.000 1.000 1.000 |
| .Nmbr_rms___FU2 1.667 0.364 4.574 0.000 1.667 0.354 |
| .Nmbr_ppl___FU2 1.253 0.150 8.375 0.000 1.253 0.718 |
| .Living_Are_FU2 1.362 0.112 12.187 0.000 1.362 0.714 |
| .Grdn_yrd_FU2_2 0.134 0.012 11.147 0.000 0.134 0.634 |
| .Rglr_tr_PI_FU2 0.089 0.009 9.835 0.000 0.089 0.680 |
| .MntHS_BC19_FU2 0.260 0.072 3.615 0.000 0.260 0.248 |
| .PhysH_BC19_FU2 0.607 0.051 12.012 0.000 0.607 0.617 |
| .C19_fnncl__FU2 0.698 0.068 10.314 0.000 0.698 0.607 |
| .Cncrnd_LIS_FU2 0.673 0.103 6.518 0.000 0.673 0.404 |
| .Rstrctns_s_FU2 0.787 0.091 8.609 0.000 0.787 0.564 |
| .depression 0.282 0.028 10.157 0.000 0.282 0.510 |
| .anxiety 0.200 0.023 8.669 0.000 0.200 0.605 |
| .SPQ_total 0.724 0.059 12.251 0.000 0.724 0.711 |
| .Mscore.g 0.208 0.008 26.634 0.000 0.208 0.878 |
| .drug_score.g 0.121 0.009 12.953 0.000 0.121 0.877 |
| .Alcohol_FU2 4.586 0.161 28.528 0.000 4.586 0.926 |
| .exerciseMn_FU2 1.238 0.077 16.139 0.000 1.238 0.933 |
| .Sleep_week_FU2 0.352 0.026 13.710 0.000 0.352 0.876 |
| livCond 3.042 0.457 6.651 0.000 1.000 1.000 |
| .healthBeforC19 0.040 0.009 4.546 0.000 0.963 0.963 |
| Defined Parameters: |
| Estimate Std.Err z-value P(>\|z\|) Std.lv Std.all |
| indirect1 0.019 0.007 2.561 0.010 0.019 0.025 |
| indirect2 0.013 0.009 1.353 0.176 0.013 0.017 |
| indirect3 0.002 0.003 0.531 0.596 0.002 0.002 |
| indirect4 0.003 0.003 0.881 0.378 0.003 0.004 |
| indirect5 0.008 0.009 0.894 0.371 0.008 0.010 |
| indirect6 0.017 0.007 2.643 0.008 0.017 0.030 |
| indirect7 0.013 0.008 1.550 0.121 0.013 0.023 |
| indirect8 -0.001 0.002 -0.337 0.736 -0.001 -0.001 |
| indirect9 -0.001 0.002 -0.419 0.675 -0.001 -0.002 |
| indirect10 0.001 0.008 0.106 0.916 0.001 0.002 |
| indirect11 0.030 0.012 2.433 0.015 0.030 0.030 |
| indirect12 0.010 0.013 0.789 0.430 0.010 0.010 |
| indirect13 -0.001 0.004 -0.384 0.701 -0.001 -0.001 |
| indirect14 -0.001 0.004 -0.162 0.871 -0.001 -0.001 |
| indirect15 0.024 0.014 1.746 0.081 0.024 0.024 |
| total1 0.323 0.047 6.852 0.000 0.323 0.434 |
| total2 0.317 0.045 7.065 0.000 0.317 0.426 |
| total3 0.306 0.048 6.362 0.000 0.306 0.411 |
| total4 0.307 0.047 6.490 0.000 0.307 0.413 |
| total5 0.312 0.045 6.993 0.000 0.312 0.420 |
| total6 0.162 0.036 4.459 0.000 0.162 0.282 |
| total7 0.157 0.036 4.386 0.000 0.157 0.274 |
| total8 0.144 0.036 3.975 0.000 0.144 0.251 |
| total9 0.143 0.036 3.975 0.000 0.143 0.250 |
| total10 0.145 0.033 4.351 0.000 0.145 0.253 |
| total11 0.120 0.059 2.050 0.040 0.120 0.119 |
| total12 0.101 0.057 1.767 0.077 0.101 0.100 |
| total13 0.089 0.060 1.488 0.137 0.089 0.088 |
| total14 0.089 0.059 1.503 0.133 0.089 0.089 |
| total15 0.114 0.056 2.036 0.042 0.114 0.113 |

| 6.1.4 Fourth Timepoint |
| --- |
| Estimator ML |
| Optimization method NLMINB |
| Number of free parameters 108 |
| Used Total |
| Number of observations 452 478 |
| Model Test User Model: |
| Test statistic 413.528 |
| Degrees of freedom 135 |
| P-value (Chi-square) 0.000 |
| Model Test Baseline Model: |
| Test statistic 2315.281 |
| Degrees of freedom 225 |
| P-value 0.000 |
| User Model versus Baseline Model: |
| Comparative Fit Index (CFI) 0.867 |
| Tucker-Lewis Index (TLI) 0.778 |
| Loglikelihood and Information Criteria: |
| Loglikelihood user model (H0) -9247.599 |
| Loglikelihood unrestricted model (H1) -9040.835 |
| Akaike (AIC) 18711.199 |
| Bayesian (BIC) 19155.476 |
| Sample-size adjusted Bayesian (BIC) 18812.723 |
| Root Mean Square Error of Approximation: |
| RMSEA 0.068 |
| 90 Percent confidence interval - lower 0.060 |
| 90 Percent confidence interval - upper 0.075 |
| P-value RMSEA <= 0.05 0.000 |
| Standardized Root Mean Square Residual: |
| SRMR 0.058 |
| Parameter Estimates: |
| Standard errors Bootstrap |
| Number of requested bootstrap draws 1000 |
| Number of successful bootstrap draws 1000 |
| Latent Variables: |
| Estimate Std.Err z-value P(>\|z\|) Std.lv Std.all |
| livCond =~ |
| Nmbr_rms___FU3 1.000 1.773 0.846 |
| Nmbr_ppl___FU3 0.399 0.034 11.721 0.000 0.707 0.605 |
| Living_Are_FU3 0.346 0.051 6.763 0.000 0.613 0.456 |
| Grdn_yrd_FU3_2 0.164 0.019 8.692 0.000 0.291 0.616 |
| healthBeforeCo19 =~ |
| Rglr_tr_PI_FU3 1.000 0.145 0.434 |
| MntHS_BC19_FU3 -5.618 1.279 -4.392 0.000 -0.816 -0.813 |
| PhysH_BC19_FU3 -3.995 0.877 -4.553 0.000 -0.580 -0.633 |
| Co19_stress =~ |
| C19_fnncl__FU3 0.577 0.072 7.957 0.000 0.577 0.583 |
| Cncrnd_LIS_FU3 0.893 0.062 14.522 0.000 0.893 0.759 |
| Rstrctns_s_FU3 0.740 0.066 11.254 0.000 0.740 0.601 |
| Regressions: |
| Estimate Std.Err z-value P(>\|z\|) Std.lv Std.all |
| depression ~ |
| hlthBC19 -2.027 0.461 -4.401 0.000 -0.294 -0.394 |
| Cnt__FU3 -0.052 0.082 -0.637 0.524 -0.052 -0.029 |
| age_FU3 -0.004 0.002 -2.154 0.031 -0.004 -0.082 |
| Gender 0.004 0.075 0.058 0.954 0.004 0.002 |
| Hgh__FU3 -0.052 0.024 -2.186 0.029 -0.052 -0.095 |
| livCond -0.010 0.020 -0.476 0.634 -0.017 -0.023 |
| anxiety ~ |
| hlthBC19 -1.550 0.328 -4.720 0.000 -0.225 -0.416 |
| Cnt__FU3 -0.154 0.064 -2.429 0.015 -0.154 -0.117 |
| age_FU3 -0.006 0.001 -4.535 0.000 -0.006 -0.175 |
| Gender 0.009 0.059 0.154 0.878 0.009 0.007 |
| Hgh__FU3 -0.056 0.017 -3.247 0.001 -0.056 -0.143 |
| livCond -0.011 0.015 -0.697 0.486 -0.019 -0.035 |
| SPQ_total ~ |
| hlthBC19 -2.660 0.643 -4.134 0.000 -0.386 -0.398 |
| Cnt__FU3 -0.058 0.130 -0.445 0.656 -0.058 -0.024 |
| age_FU3 -0.007 0.003 -2.659 0.008 -0.007 -0.109 |
| Gender -0.202 0.104 -1.949 0.051 -0.202 -0.089 |
| Hgh__FU3 -0.123 0.033 -3.766 0.000 -0.123 -0.174 |
| livCond -0.044 0.027 -1.642 0.101 -0.078 -0.081 |
| media_score_FU3.g ~ |
| hlthBC19 -0.334 0.187 -1.790 0.074 -0.048 -0.105 |
| Cnt__FU3 -0.053 0.052 -1.026 0.305 -0.053 -0.048 |
| age_FU3 -0.001 0.002 -0.469 0.639 -0.001 -0.023 |
| Gender -0.171 0.052 -3.291 0.001 -0.171 -0.159 |
| Hgh__FU3 -0.039 0.016 -2.447 0.014 -0.039 -0.115 |
| livCond -0.021 0.014 -1.423 0.155 -0.036 -0.079 |
| drug_score_during_FU3.g ~ |
| hlthBC19 -0.590 0.153 -3.865 0.000 -0.086 -0.228 |
| Cnt__FU3 -0.003 0.044 -0.067 0.947 -0.003 -0.003 |
| age_FU3 0.001 0.001 1.162 0.245 0.001 0.050 |
| Gender 0.015 0.042 0.370 0.712 0.015 0.018 |
| Hgh__FU3 -0.038 0.014 -2.664 0.008 -0.038 -0.139 |
| livCond -0.023 0.011 -2.130 0.033 -0.040 -0.107 |
| Alcohol_FU3 ~ |
| hlthBC19 2.950 1.115 2.647 0.008 0.428 0.189 |
| Cnt__FU3 0.050 0.251 0.198 0.843 0.050 0.009 |
| age_FU3 0.027 0.007 3.697 0.000 0.027 0.174 |
| Gender -0.381 0.254 -1.497 0.135 -0.381 -0.072 |
| Hgh__FU3 0.258 0.082 3.151 0.002 0.258 0.157 |
| livCond 0.092 0.063 1.457 0.145 0.162 0.072 |
| exerciseMin_FU3 ~ |
| hlthBC19 0.210 0.487 0.430 0.667 0.030 0.027 |
| Cnt__FU3 -0.062 0.144 -0.431 0.666 -0.062 -0.023 |
| age_FU3 0.001 0.004 0.318 0.751 0.001 0.017 |
| Gender -0.049 0.131 -0.375 0.707 -0.049 -0.019 |
| Hgh__FU3 0.138 0.036 3.783 0.000 0.138 0.171 |
| livCond 0.024 0.041 0.591 0.554 0.043 0.039 |
| Sleep_week_FU3 ~ |
| hlthBC19 0.310 0.165 1.884 0.059 0.045 0.113 |
| Cnt__FU3 0.034 0.049 0.689 0.491 0.034 0.035 |
| age_FU3 0.000 0.001 0.069 0.945 0.000 0.003 |
| Gender 0.045 0.047 0.950 0.342 0.045 0.048 |
| Hgh__FU3 0.037 0.015 2.524 0.012 0.037 0.128 |
| livCond 0.002 0.012 0.150 0.881 0.003 0.008 |
| healthBeforeCo19 ~ |
| Cnt__FU3 0.044 0.021 2.097 0.036 0.305 0.126 |
| age_FU3 0.001 0.001 1.730 0.084 0.007 0.099 |
| Gender 0.008 0.019 0.457 0.648 0.058 0.025 |
| Hgh__FU3 -0.008 0.006 -1.333 0.182 -0.055 -0.075 |
| livCond -0.003 0.006 -0.521 0.602 -0.035 -0.035 |
| depression ~ |
| C19_strs (c1) 0.351 0.050 7.084 0.000 0.351 0.470 |
| anxiety ~ |
| C19_strs (c2) 0.198 0.035 5.634 0.000 0.198 0.366 |
| SPQ_total ~ |
| C19_strs (c3) 0.202 0.059 3.439 0.001 0.202 0.208 |
| media_score_FU3.g ~ |
| C19_strs (a1) 0.037 0.029 1.292 0.196 0.037 0.081 |
| drug_score_during_FU3.g ~ |
| C19_strs (a2) 0.064 0.027 2.365 0.018 0.064 0.170 |
| Alcohol_FU3 ~ |
| C19_strs (a3) 0.331 0.139 2.387 0.017 0.331 0.146 |
| exerciseMin_FU3 ~ |
| C19_strs (a4) -0.095 0.060 -1.579 0.114 -0.095 -0.086 |
| Sleep_week_FU3 ~ |
| C19_strs (a5) -0.086 0.028 -3.036 0.002 -0.086 -0.214 |
| depression ~ |
| md__FU3. (b1) 0.106 0.072 1.479 0.139 0.106 0.066 |
| d___FU3. (b2) 0.118 0.106 1.114 0.265 0.118 0.059 |
| Alch_FU3 (b3) 0.005 0.015 0.361 0.718 0.005 0.016 |
| exrM_FU3 (b4) 0.006 0.029 0.212 0.832 0.006 0.009 |
| Slp__FU3 (b5) -0.095 0.087 -1.086 0.278 -0.095 -0.051 |
| anxiety ~ |
| md__FU3. (b6) 0.116 0.050 2.304 0.021 0.116 0.099 |
| d___FU3. (b7) -0.005 0.079 -0.069 0.945 -0.005 -0.004 |
| Alch_FU3 (b8) -0.003 0.011 -0.250 0.802 -0.003 -0.011 |
| exrM_FU3 (b9) 0.057 0.023 2.444 0.015 0.057 0.117 |
| Slp__FU3 (b10) -0.017 0.067 -0.261 0.794 -0.017 -0.013 |
| SPQ_total ~ |
| md__FU3. (b11) 0.023 0.092 0.255 0.799 0.023 0.011 |
| d___FU3. (b12) 0.149 0.137 1.088 0.277 0.149 0.058 |
| Alch_FU3 (b13) -0.030 0.020 -1.489 0.136 -0.030 -0.070 |
| exrM_FU3 (b14) 0.035 0.043 0.805 0.421 0.035 0.040 |
| Slp__FU3 (b15) -0.015 0.123 -0.119 0.906 -0.015 -0.006 |
| Covariances: |
| Estimate Std.Err z-value P(>\|z\|) Std.lv Std.all |
| .anxiety ~~ |
| .SPQ_total 0.168 0.027 6.150 0.000 0.168 0.505 |
| .depression ~~ |
| .SPQ_total 0.147 0.031 4.803 0.000 0.147 0.330 |
| .anxiety 0.107 0.020 5.278 0.000 0.107 0.473 |
| livCond ~~ |
| Co19_stress -0.265 0.115 -2.316 0.021 -0.150 -0.150 |
| Variances: |
| Estimate Std.Err z-value P(>\|z\|) Std.lv Std.all |
| Co19_stress 1.000 1.000 1.000 |
| .Nmbr_rms___FU3 1.247 0.319 3.911 0.000 1.247 0.284 |
| .Nmbr_ppl___FU3 0.865 0.061 14.072 0.000 0.865 0.634 |
| .Living_Are_FU3 1.427 0.102 13.939 0.000 1.427 0.792 |
| .Grdn_yrd_FU3_2 0.139 0.012 12.040 0.000 0.139 0.621 |
| .Rglr_tr_PI_FU3 0.091 0.010 9.293 0.000 0.091 0.811 |
| .MntHS_BC19_FU3 0.341 0.086 3.966 0.000 0.341 0.339 |
| .PhysH_BC19_FU3 0.502 0.053 9.530 0.000 0.502 0.599 |
| .C19_fnncl__FU3 0.645 0.071 9.032 0.000 0.645 0.660 |
| .Cncrnd_LIS_FU3 0.586 0.088 6.691 0.000 0.586 0.424 |
| .Rstrctns_s_FU3 0.968 0.091 10.643 0.000 0.968 0.639 |
| .depression 0.303 0.032 9.540 0.000 0.303 0.542 |
| .anxiety 0.168 0.020 8.198 0.000 0.168 0.572 |
| .SPQ_total 0.657 0.059 11.038 0.000 0.657 0.698 |
| .medi_scr_FU3.g 0.200 0.010 20.910 0.000 0.200 0.936 |
| .drg_scr_d_FU3. 0.126 0.010 12.212 0.000 0.126 0.890 |
| .Alcohol_FU3 4.492 0.201 22.352 0.000 4.492 0.874 |
| .exerciseMn_FU3 1.183 0.079 14.905 0.000 1.183 0.959 |
| .Sleep_week_FU3 0.148 0.012 12.820 0.000 0.148 0.924 |
| livCond 3.143 0.402 7.825 0.000 1.000 1.000 |
| .healthBeforC19 0.020 0.007 2.853 0.004 0.965 0.965 |
| Defined Parameters: |
| Estimate Std.Err z-value P(>\|z\|) Std.lv Std.all |
| indirect1 0.004 0.004 0.950 0.342 0.004 0.005 |
| indirect2 0.008 0.008 0.988 0.323 0.008 0.010 |
| indirect3 0.002 0.005 0.339 0.735 0.002 0.002 |
| indirect4 -0.001 0.004 -0.169 0.866 -0.001 -0.001 |
| indirect5 0.008 0.008 1.031 0.303 0.008 0.011 |
| indirect6 0.004 0.004 1.111 0.266 0.004 0.008 |
| indirect7 -0.000 0.006 -0.062 0.951 -0.000 -0.001 |
| indirect8 -0.001 0.004 -0.230 0.818 -0.001 -0.002 |
| indirect9 -0.005 0.005 -1.174 0.240 -0.005 -0.010 |
| indirect10 0.001 0.006 0.246 0.805 0.001 0.003 |
| indirect11 0.001 0.004 0.204 0.839 0.001 0.001 |
| indirect12 0.010 0.010 0.967 0.334 0.010 0.010 |
| indirect13 -0.010 0.008 -1.210 0.226 -0.010 -0.010 |
| indirect14 -0.003 0.005 -0.627 0.531 -0.003 -0.003 |
| indirect15 0.001 0.011 0.115 0.909 0.001 0.001 |
| total1 0.355 0.050 7.120 0.000 0.355 0.475 |
| total2 0.359 0.049 7.276 0.000 0.359 0.480 |
| total3 0.353 0.048 7.290 0.000 0.353 0.472 |
| total4 0.351 0.049 7.150 0.000 0.351 0.469 |
| total5 0.359 0.047 7.589 0.000 0.359 0.481 |
| total6 0.203 0.036 5.665 0.000 0.203 0.374 |
| total7 0.198 0.034 5.759 0.000 0.198 0.366 |
| total8 0.198 0.035 5.716 0.000 0.198 0.365 |
| total9 0.193 0.035 5.537 0.000 0.193 0.356 |
| total10 0.200 0.033 5.989 0.000 0.200 0.369 |
| total11 0.203 0.059 3.453 0.001 0.203 0.209 |
| total12 0.211 0.058 3.655 0.000 0.211 0.218 |
| total13 0.192 0.058 3.301 0.001 0.192 0.198 |
| total14 0.198 0.058 3.412 0.001 0.198 0.205 |
| total15 0.203 0.056 3.611 0.000 0.203 0.209 |

#### 6.2 ‘Social adversity’ Model – first to forth timepoint

| 6.2.1 First TImepoint |
| --- |
| Estimator ML |
| Optimization method NLMINB |
| Number of free parameters 108 |
| Number of observations 480 |
| Model Test User Model: |
| Test statistic 492.394 |
| Degrees of freedom 135 |
| P-value (Chi-square) 0.000 |
| Model Test Baseline Model: |
| Test statistic 2653.514 |
| Degrees of freedom 225 |
| P-value 0.000 |
| User Model versus Baseline Model: |
| Comparative Fit Index (CFI) 0.853 |
| Tucker-Lewis Index (TLI) 0.755 |
| Loglikelihood and Information Criteria: |
| Loglikelihood user model (H0) -10089.290 |
| Loglikelihood unrestricted model (H1) -9843.093 |
| Akaike (AIC) 20394.580 |
| Bayesian (BIC) 20845.349 |
| Sample-size adjusted Bayesian (BIC) 20502.568 |
| Root Mean Square Error of Approximation: |
| RMSEA 0.074 |
| 90 Percent confidence interval - lower 0.067 |
| 90 Percent confidence interval - upper 0.081 |
| P-value RMSEA <= 0.05 0.000 |
| Standardized Root Mean Square Residual: |
| SRMR 0.082 |
| Parameter Estimates: |
| Standard errors Bootstrap |
| Number of requested bootstrap draws 1000 |
| Number of successful bootstrap draws 1000 |
| Latent Variables: |
| Estimate Std.Err z-value P(>\|z\|) Std.lv Std.all |
| livCond =~ |
| Nmbr_rms_hs_fl 1.000 2.182 0.783 |
| Nmbr_ppl_pr_hs 0.236 0.023 10.085 0.000 0.516 0.649 |
| LivingArea 0.323 0.030 10.671 0.000 0.704 0.521 |
| Garden_yard2 0.134 0.014 9.268 0.000 0.293 0.635 |
| healthBeforeCo19 =~ |
| Rglr_trtmnt_PI 1.000 0.155 0.457 |
| MntlHlthS_BC19 5.886 1.056 5.574 0.000 0.915 0.830 |
| PhysclHlt_BC19 3.673 0.695 5.286 0.000 0.571 0.578 |
| lonely =~ |
| Lonely_DrngC19 0.860 0.061 14.086 0.000 0.860 0.678 |
| Ngtv_thgh_DC19 0.813 0.046 17.639 0.000 0.813 0.754 |
| ChngSclRltn_st 0.765 0.061 12.575 0.000 0.765 0.566 |
| Regressions: |
| Estimate Std.Err z-value P(>\|z\|) Std.lv Std.all |
| depression ~ |
| hlthBC19 1.580 0.297 5.315 0.000 0.246 0.368 |
| Cntry_rs -0.304 0.057 -5.366 0.000 -0.304 -0.205 |
| Age 0.020 0.023 0.887 0.375 0.020 0.030 |
| Gender 0.063 0.048 1.298 0.194 0.063 0.041 |
| Hghst___ -0.051 0.018 -2.777 0.005 -0.051 -0.091 |
| livCond 0.023 0.013 1.687 0.092 0.049 0.074 |
| anxiety ~ |
| hlthBC19 1.198 0.252 4.753 0.000 0.186 0.336 |
| Cntry_rs -0.260 0.059 -4.417 0.000 -0.260 -0.211 |
| Age -0.045 0.021 -2.089 0.037 -0.045 -0.081 |
| Gender 0.024 0.044 0.537 0.591 0.024 0.019 |
| Hghst___ -0.005 0.019 -0.269 0.788 -0.005 -0.011 |
| livCond 0.017 0.014 1.159 0.246 0.036 0.065 |
| SPQ_total ~ |
| hlthBC19 2.472 0.506 4.889 0.000 0.384 0.403 |
| Cntry_rs 0.012 0.109 0.112 0.911 0.012 0.006 |
| Age -0.070 0.047 -1.503 0.133 -0.070 -0.074 |
| Gender -0.149 0.090 -1.658 0.097 -0.149 -0.069 |
| Hghst___ -0.069 0.039 -1.774 0.076 -0.069 -0.087 |
| livCond -0.014 0.025 -0.545 0.585 -0.030 -0.031 |
| Mscore.g ~ |
| hlthBC19 0.043 0.196 0.219 0.827 0.007 0.014 |
| Cntry_rs -0.171 0.050 -3.389 0.001 -0.171 -0.166 |
| Age -0.065 0.021 -3.028 0.002 -0.065 -0.139 |
| Gender -0.001 0.046 -0.015 0.988 -0.001 -0.001 |
| Hghst___ -0.049 0.017 -2.872 0.004 -0.049 -0.127 |
| livCond -0.019 0.012 -1.558 0.119 -0.041 -0.088 |
| drug_score.g ~ |
| hlthBC19 0.454 0.153 2.958 0.003 0.071 0.213 |
| Cntry_rs 0.010 0.036 0.287 0.774 0.010 0.014 |
| Age 0.032 0.016 2.029 0.042 0.032 0.095 |
| Gender -0.003 0.033 -0.088 0.930 -0.003 -0.004 |
| Hghst___ -0.008 0.013 -0.644 0.519 -0.008 -0.029 |
| livCond -0.002 0.008 -0.238 0.812 -0.004 -0.012 |
| Alcohol_DuringCo19 ~ |
| hlthBC19 -2.456 0.953 -2.578 0.010 -0.382 -0.167 |
| Cntry_rs -0.512 0.229 -2.238 0.025 -0.512 -0.101 |
| Age 0.507 0.102 4.991 0.000 0.507 0.221 |
| Gender -0.470 0.229 -2.047 0.041 -0.470 -0.091 |
| Hghst___ 0.038 0.088 0.432 0.666 0.038 0.020 |
| livCond 0.068 0.056 1.200 0.230 0.148 0.065 |
| Exercise_DuringCo19 ~ |
| hlthBC19 -1.301 0.571 -2.277 0.023 -0.202 -0.162 |
| Cntry_rs -0.523 0.138 -3.793 0.000 -0.523 -0.189 |
| Age 0.175 0.059 2.988 0.003 0.175 0.140 |
| Gender -0.063 0.129 -0.490 0.624 -0.063 -0.022 |
| Hghst___ 0.035 0.046 0.774 0.439 0.035 0.034 |
| livCond -0.006 0.030 -0.191 0.849 -0.013 -0.010 |
| Sleep_week_DuringCo19 ~ |
| hlthBC19 0.153 0.254 0.600 0.548 0.024 0.037 |
| Cntry_rs -0.089 0.070 -1.283 0.199 -0.089 -0.063 |
| Age -0.093 0.031 -3.025 0.002 -0.093 -0.144 |
| Gender 0.094 0.060 1.550 0.121 0.094 0.065 |
| Hghst___ -0.036 0.024 -1.508 0.132 -0.036 -0.068 |
| livCond -0.023 0.015 -1.513 0.130 -0.051 -0.080 |
| healthBeforeCo19 ~ |
| Cntry_rs -0.058 0.021 -2.732 0.006 -0.374 -0.169 |
| Age -0.024 0.010 -2.485 0.013 -0.154 -0.154 |
| Gender 0.043 0.020 2.161 0.031 0.275 0.122 |
| Hghst___ -0.019 0.008 -2.500 0.012 -0.124 -0.148 |
| livCond -0.008 0.005 -1.453 0.146 -0.110 -0.110 |
| depression ~ |
| lonely (c1) 0.474 0.035 13.362 0.000 0.474 0.710 |
| anxiety ~ |
| lonely (c2) 0.253 0.033 7.582 0.000 0.253 0.456 |
| SPQ_total ~ |
| lonely (c3) 0.118 0.058 2.021 0.043 0.118 0.123 |
| Mscore.g ~ |
| lonely (a1) 0.050 0.027 1.866 0.062 0.050 0.108 |
| drug_score.g ~ |
| lonely (a2) 0.065 0.020 3.223 0.001 0.065 0.196 |
| Alcohol_DuringCo19 ~ |
| lonely (a3) 0.497 0.127 3.905 0.000 0.497 0.218 |
| Exercise_DuringCo19 ~ |
| lonely (a4) 0.029 0.078 0.366 0.715 0.029 0.023 |
| Sleep_week_DuringCo19 ~ |
| lonely (a5) -0.158 0.038 -4.196 0.000 -0.158 -0.246 |
| depression ~ |
| Mscore.g (b1) 0.040 0.050 0.789 0.430 0.040 0.028 |
| drg_scr. (b2) -0.031 0.083 -0.375 0.708 -0.031 -0.015 |
| Alc_DC19 (b3) 0.005 0.011 0.436 0.663 0.005 0.016 |
| Exr_DC19 (b4) -0.029 0.018 -1.586 0.113 -0.029 -0.055 |
| Sl__DC19 (b5) -0.034 0.038 -0.894 0.371 -0.034 -0.033 |
| anxiety ~ |
| Mscore.g (b6) 0.063 0.052 1.211 0.226 0.063 0.053 |
| drg_scr. (b7) -0.009 0.085 -0.106 0.915 -0.009 -0.005 |
| Alc_DC19 (b8) -0.007 0.010 -0.694 0.487 -0.007 -0.030 |
| Exr_DC19 (b9) 0.001 0.020 0.061 0.951 0.001 0.003 |
| Sl__DC19 (b10) -0.024 0.038 -0.637 0.524 -0.024 -0.028 |
| SPQ_total ~ |
| Mscore.g (b11) 0.213 0.093 2.294 0.022 0.213 0.104 |
| drg_scr. (b12) 0.133 0.142 0.931 0.352 0.133 0.046 |
| Alc_DC19 (b13) 0.002 0.020 0.125 0.901 0.002 0.006 |
| Exr_DC19 (b14) -0.044 0.034 -1.277 0.202 -0.044 -0.058 |
| Sl__DC19 (b15) -0.085 0.069 -1.235 0.217 -0.085 -0.057 |
| Covariances: |
| Estimate Std.Err z-value P(>\|z\|) Std.lv Std.all |
| .anxiety ~~ |
| .SPQ_total 0.125 0.023 5.436 0.000 0.125 0.366 |
| .depression ~~ |
| .SPQ_total 0.034 0.021 1.592 0.111 0.034 0.121 |
| .anxiety 0.036 0.012 3.070 0.002 0.036 0.254 |
| livCond ~~ |
| lonely -0.355 0.146 -2.427 0.015 -0.162 -0.162 |
| Variances: |
| Estimate Std.Err z-value P(>\|z\|) Std.lv Std.all |
| lonely 1.000 1.000 1.000 |
| .Nmbr_rms_hs_fl 3.014 0.473 6.376 0.000 3.014 0.388 |
| .Nmbr_ppl_pr_hs 0.365 0.028 13.020 0.000 0.365 0.579 |
| .LivingArea 1.330 0.097 13.752 0.000 1.330 0.728 |
| .Garden_yard2 0.127 0.010 12.671 0.000 0.127 0.596 |
| .Rglr_trtmnt_PI 0.092 0.009 10.016 0.000 0.092 0.792 |
| .MntlHlthS_BC19 0.378 0.103 3.657 0.000 0.378 0.311 |
| .PhysclHlt_BC19 0.651 0.053 12.182 0.000 0.651 0.666 |
| .Lonely_DrngC19 0.871 0.076 11.395 0.000 0.871 0.541 |
| .Ngtv_thgh_DC19 0.502 0.057 8.801 0.000 0.502 0.432 |
| .ChngSclRltn_st 1.242 0.090 13.764 0.000 1.242 0.680 |
| .depression 0.118 0.015 7.746 0.000 0.118 0.264 |
| .anxiety 0.173 0.018 9.867 0.000 0.173 0.566 |
| .SPQ_total 0.667 0.060 11.204 0.000 0.667 0.733 |
| .Mscore.g 0.194 0.010 20.310 0.000 0.194 0.903 |
| .drug_score.g 0.099 0.010 10.313 0.000 0.099 0.911 |
| .Alcohl_DrngC19 4.405 0.205 21.526 0.000 4.405 0.846 |
| .Exercs_DrngC19 1.441 0.074 19.587 0.000 1.441 0.924 |
| .Slp_wk_DrngC19 0.367 0.026 14.023 0.000 0.367 0.894 |
| livCond 4.763 0.578 8.235 0.000 1.000 1.000 |
| .healthBeforC19 0.021 0.006 3.460 0.001 0.885 0.885 |
| Defined Parameters: |
| Estimate Std.Err z-value P(>\|z\|) Std.lv Std.all |
| indirect1 0.002 0.003 0.692 0.489 0.002 0.003 |
| indirect2 -0.002 0.006 -0.345 0.730 -0.002 -0.003 |
| indirect3 0.002 0.006 0.430 0.667 0.002 0.004 |
| indirect4 -0.001 0.003 -0.304 0.761 -0.001 -0.001 |
| indirect5 0.005 0.006 0.870 0.384 0.005 0.008 |
| indirect6 0.003 0.003 0.954 0.340 0.003 0.006 |
| indirect7 -0.001 0.006 -0.103 0.918 -0.001 -0.001 |
| indirect8 -0.004 0.005 -0.655 0.513 -0.004 -0.006 |
| indirect9 0.000 0.002 0.022 0.982 0.000 0.000 |
| indirect10 0.004 0.006 0.614 0.539 0.004 0.007 |
| indirect11 0.011 0.008 1.418 0.156 0.011 0.011 |
| indirect12 0.009 0.010 0.873 0.383 0.009 0.009 |
| indirect13 0.001 0.010 0.122 0.903 0.001 0.001 |
| indirect14 -0.001 0.005 -0.273 0.785 -0.001 -0.001 |
| indirect15 0.013 0.012 1.133 0.257 0.013 0.014 |
| total1 0.476 0.035 13.436 0.000 0.476 0.713 |
| total2 0.472 0.036 13.291 0.000 0.472 0.707 |
| total3 0.477 0.034 13.963 0.000 0.477 0.714 |
| total4 0.473 0.036 13.290 0.000 0.473 0.709 |
| total5 0.480 0.036 13.433 0.000 0.480 0.718 |
| total6 0.256 0.034 7.632 0.000 0.256 0.462 |
| total7 0.252 0.033 7.642 0.000 0.252 0.455 |
| total8 0.249 0.033 7.517 0.000 0.249 0.450 |
| total9 0.253 0.033 7.587 0.000 0.253 0.456 |
| total10 0.257 0.034 7.568 0.000 0.257 0.463 |
| total11 0.128 0.058 2.207 0.027 0.128 0.134 |
| total12 0.126 0.058 2.188 0.029 0.126 0.132 |
| total13 0.119 0.057 2.076 0.038 0.119 0.125 |
| total14 0.116 0.059 1.986 0.047 0.116 0.122 |
| total15 0.131 0.056 2.331 0.020 0.131 0.137 |

| 6.2.2 Second Timepoint |
| --- |
| Estimator ML |
| Optimization method NLMINB |
| Number of free parameters 108 |
| Used Total |
| Number of observations 423 464 |
| Model Test User Model: |
| Test statistic 481.871 |
| Degrees of freedom 135 |
| P-value (Chi-square) 0.000 |
| Model Test Baseline Model: |
| Test statistic 2645.369 |
| Degrees of freedom 225 |
| P-value 0.000 |
| User Model versus Baseline Model: |
| Comparative Fit Index (CFI) 0.857 |
| Tucker-Lewis Index (TLI) 0.761 |
| Loglikelihood and Information Criteria: |
| Loglikelihood user model (H0) -8297.995 |
| Loglikelihood unrestricted model (H1) -8057.059 |
| Akaike (AIC) 16811.989 |
| Bayesian (BIC) 17249.105 |
| Sample-size adjusted Bayesian (BIC) 16906.385 |
| Root Mean Square Error of Approximation: |
| RMSEA 0.078 |
| 90 Percent confidence interval - lower 0.070 |
| 90 Percent confidence interval - upper 0.086 |
| P-value RMSEA <= 0.05 0.000 |
| Standardized Root Mean Square Residual: |
| SRMR 0.084 |
| Parameter Estimates: |
| Standard errors Bootstrap |
| Number of requested bootstrap draws 1000 |
| Number of successful bootstrap draws 1000 |
| Latent Variables: |
| Estimate Std.Err z-value P(>\|z\|) Std.lv Std.all |
| livCond =~ |
| Nmbr_rms___FU1 1.000 2.028 0.920 |
| Nmbr_ppl___FU1 0.400 0.035 11.569 0.000 0.812 0.642 |
| Living_Are_FU1 0.336 0.041 8.242 0.000 0.682 0.499 |
| Grdn_yrd_FU1_2 0.132 0.017 7.974 0.000 0.267 0.568 |
| healthBeforeCo19 =~ |
| Rglr_tr_PI_FU1 1.000 0.163 0.504 |
| MntHS_BC19_FU1 -5.060 1.315 -3.849 0.000 -0.825 -0.812 |
| PhysH_BC19_FU1 -3.036 0.675 -4.501 0.000 -0.495 -0.541 |
| lonely =~ |
| Lonely_FU1 0.758 0.065 11.705 0.000 0.758 0.636 |
| Ngtv_thgth_FU1 0.826 0.049 16.965 0.000 0.826 0.778 |
| ChngSclRl__FU1 0.742 0.069 10.697 0.000 0.742 0.616 |
| Regressions: |
| Estimate Std.Err z-value P(>\|z\|) Std.lv Std.all |
| depression ~ |
| hlthBC19 -0.944 0.284 -3.324 0.001 -0.154 -0.234 |
| Cnt__FU1 -0.357 0.070 -5.092 0.000 -0.357 -0.229 |
| age_FU1 -0.002 0.001 -1.114 0.265 -0.002 -0.039 |
| Gender -0.025 0.053 -0.466 0.641 -0.025 -0.017 |
| Hghst___ -0.010 0.019 -0.519 0.604 -0.010 -0.019 |
| livCond -0.024 0.014 -1.654 0.098 -0.048 -0.074 |
| anxiety ~ |
| hlthBC19 -0.932 0.227 -4.111 0.000 -0.152 -0.285 |
| Cnt__FU1 -0.209 0.060 -3.474 0.001 -0.209 -0.165 |
| age_FU1 -0.006 0.001 -4.865 0.000 -0.006 -0.165 |
| Gender 0.012 0.048 0.248 0.804 0.012 0.010 |
| Hghst___ -0.052 0.019 -2.767 0.006 -0.052 -0.121 |
| livCond -0.001 0.013 -0.074 0.941 -0.002 -0.004 |
| SPQ_total ~ |
| hlthBC19 -2.295 0.473 -4.851 0.000 -0.374 -0.395 |
| Cnt__FU1 -0.064 0.118 -0.546 0.585 -0.064 -0.029 |
| age_FU1 -0.005 0.002 -2.066 0.039 -0.005 -0.083 |
| Gender -0.226 0.100 -2.268 0.023 -0.226 -0.106 |
| Hghst___ -0.129 0.034 -3.758 0.000 -0.129 -0.168 |
| livCond 0.006 0.023 0.268 0.789 0.013 0.013 |
| Mscore.g ~ |
| hlthBC19 -0.181 0.175 -1.035 0.301 -0.030 -0.072 |
| Cnt__FU1 -0.131 0.047 -2.764 0.006 -0.131 -0.133 |
| age_FU1 -0.003 0.001 -2.249 0.025 -0.003 -0.117 |
| Gender -0.036 0.043 -0.832 0.405 -0.036 -0.039 |
| Hghst___ -0.080 0.016 -5.008 0.000 -0.080 -0.238 |
| livCond -0.008 0.011 -0.756 0.450 -0.017 -0.041 |
| drug_score.g ~ |
| hlthBC19 -0.164 0.186 -0.885 0.376 -0.027 -0.079 |
| Cnt__FU1 -0.098 0.045 -2.164 0.031 -0.098 -0.122 |
| age_FU1 -0.000 0.001 -0.224 0.823 -0.000 -0.010 |
| Gender 0.026 0.035 0.750 0.453 0.026 0.035 |
| Hghst___ -0.035 0.015 -2.384 0.017 -0.035 -0.129 |
| livCond -0.021 0.008 -2.630 0.009 -0.044 -0.129 |
| Alcohol_FU1 ~ |
| hlthBC19 1.037 1.001 1.036 0.300 0.169 0.079 |
| Cnt__FU1 -0.408 0.245 -1.668 0.095 -0.408 -0.080 |
| age_FU1 0.021 0.007 2.995 0.003 0.021 0.152 |
| Gender -0.448 0.246 -1.817 0.069 -0.448 -0.092 |
| Hghst___ 0.174 0.088 1.979 0.048 0.174 0.100 |
| livCond 0.064 0.056 1.144 0.253 0.130 0.061 |
| exerciseMin_FU1 ~ |
| hlthBC19 0.851 0.518 1.642 0.101 0.139 0.134 |
| Cnt__FU1 -0.254 0.135 -1.879 0.060 -0.254 -0.103 |
| age_FU1 -0.001 0.003 -0.188 0.851 -0.001 -0.010 |
| Gender -0.174 0.125 -1.392 0.164 -0.174 -0.074 |
| Hghst___ 0.059 0.041 1.449 0.147 0.059 0.070 |
| livCond -0.004 0.026 -0.169 0.866 -0.009 -0.009 |
| Sleep_week_FU1 ~ |
| hlthBC19 0.216 0.151 1.425 0.154 0.035 0.102 |
| Cnt__FU1 0.071 0.042 1.697 0.090 0.071 0.086 |
| age_FU1 -0.002 0.001 -2.154 0.031 -0.002 -0.100 |
| Gender 0.056 0.037 1.507 0.132 0.056 0.072 |
| Hghst___ 0.024 0.016 1.505 0.132 0.024 0.087 |
| livCond -0.000 0.009 -0.031 0.975 -0.001 -0.002 |
| healthBeforeCo19 ~ |
| Cnt__FU1 0.087 0.030 2.873 0.004 0.532 0.224 |
| age_FU1 0.001 0.001 1.402 0.161 0.005 0.084 |
| Gender 0.020 0.020 1.025 0.305 0.123 0.054 |
| Hghst___ 0.015 0.010 1.409 0.159 0.089 0.110 |
| livCond 0.017 0.007 2.436 0.015 0.210 0.210 |
| depression ~ |
| lonely (c1) 0.463 0.040 11.699 0.000 0.463 0.705 |
| anxiety ~ |
| lonely (c2) 0.268 0.041 6.567 0.000 0.268 0.503 |
| SPQ_total ~ |
| lonely (c3) 0.237 0.068 3.476 0.001 0.237 0.250 |
| Mscore.g ~ |
| lonely (a1) 0.042 0.027 1.546 0.122 0.042 0.101 |
| drug_score.g ~ |
| lonely (a2) 0.049 0.028 1.770 0.077 0.049 0.144 |
| Alcohol_FU1 ~ |
| lonely (a3) -0.084 0.148 -0.568 0.570 -0.084 -0.039 |
| exerciseMin_FU1 ~ |
| lonely (a4) -0.055 0.073 -0.749 0.454 -0.055 -0.053 |
| Sleep_week_FU1 ~ |
| lonely (a5) -0.106 0.023 -4.590 0.000 -0.106 -0.307 |
| depression ~ |
| Mscore.g (b1) 0.057 0.062 0.929 0.353 0.057 0.036 |
| drg_scr. (b2) 0.102 0.095 1.082 0.279 0.102 0.053 |
| Alch_FU1 (b3) 0.019 0.011 1.637 0.102 0.019 0.061 |
| exrM_FU1 (b4) -0.030 0.024 -1.249 0.212 -0.030 -0.048 |
| Slp__FU1 (b5) 0.062 0.079 0.788 0.431 0.062 0.032 |
| anxiety ~ |
| Mscore.g (b6) 0.087 0.056 1.573 0.116 0.087 0.068 |
| drg_scr. (b7) 0.093 0.080 1.158 0.247 0.093 0.059 |
| Alch_FU1 (b8) 0.002 0.011 0.175 0.861 0.002 0.007 |
| exrM_FU1 (b9) -0.006 0.021 -0.275 0.784 -0.006 -0.011 |
| Slp__FU1 (b10) -0.014 0.073 -0.198 0.843 -0.014 -0.009 |
| SPQ_total ~ |
| Mscore.g (b11) 0.247 0.102 2.423 0.015 0.247 0.107 |
| drg_scr. (b12) 0.178 0.124 1.431 0.152 0.178 0.063 |
| Alch_FU1 (b13) 0.011 0.019 0.558 0.577 0.011 0.024 |
| exrM_FU1 (b14) 0.003 0.041 0.061 0.951 0.003 0.003 |
| Slp__FU1 (b15) -0.225 0.143 -1.576 0.115 -0.225 -0.082 |
| Covariances: |
| Estimate Std.Err z-value P(>\|z\|) Std.lv Std.all |
| .anxiety ~~ |
| .SPQ_total 0.121 0.022 5.449 0.000 0.121 0.433 |
| .depression ~~ |
| .SPQ_total 0.069 0.023 2.933 0.003 0.069 0.246 |
| .anxiety 0.056 0.014 3.870 0.000 0.056 0.397 |
| livCond ~~ |
| lonely -0.054 0.128 -0.424 0.671 -0.027 -0.027 |
| Variances: |
| Estimate Std.Err z-value P(>\|z\|) Std.lv Std.all |
| lonely 1.000 1.000 1.000 |
| .Nmbr_rms___FU1 0.749 0.317 2.366 0.018 0.749 0.154 |
| .Nmbr_ppl___FU1 0.939 0.075 12.491 0.000 0.939 0.587 |
| .Living_Are_FU1 1.407 0.095 14.871 0.000 1.407 0.751 |
| .Grdn_yrd_FU1_2 0.150 0.011 13.839 0.000 0.150 0.678 |
| .Rglr_tr_PI_FU1 0.078 0.009 8.586 0.000 0.078 0.746 |
| .MntHS_BC19_FU1 0.352 0.134 2.627 0.009 0.352 0.341 |
| .PhysH_BC19_FU1 0.592 0.065 9.121 0.000 0.592 0.707 |
| .Lonely_FU1 0.847 0.088 9.638 0.000 0.847 0.596 |
| .Ngtv_thgth_FU1 0.445 0.055 8.154 0.000 0.445 0.395 |
| .ChngSclRl__FU1 0.898 0.075 11.918 0.000 0.898 0.620 |
| .depression 0.140 0.019 7.449 0.000 0.140 0.325 |
| .anxiety 0.141 0.019 7.359 0.000 0.141 0.498 |
| .SPQ_total 0.558 0.048 11.590 0.000 0.558 0.621 |
| .Mscore.g 0.150 0.011 13.202 0.000 0.150 0.883 |
| .drug_score.g 0.104 0.011 9.929 0.000 0.104 0.912 |
| .Alcohol_FU1 4.302 0.185 23.263 0.000 4.302 0.935 |
| .exerciseMn_FU1 1.028 0.073 14.169 0.000 1.028 0.962 |
| .Sleep_week_FU1 0.102 0.010 9.704 0.000 0.102 0.862 |
| livCond 4.114 0.490 8.396 0.000 1.000 1.000 |
| .healthBeforC19 0.023 0.007 3.245 0.001 0.883 0.883 |
| Defined Parameters: |
| Estimate Std.Err z-value P(>\|z\|) Std.lv Std.all |
| indirect1 0.002 0.003 0.733 0.464 0.002 0.004 |
| indirect2 0.005 0.006 0.872 0.383 0.005 0.008 |
| indirect3 -0.002 0.004 -0.426 0.670 -0.002 -0.002 |
| indirect4 0.002 0.003 0.566 0.571 0.002 0.003 |
| indirect5 -0.007 0.009 -0.734 0.463 -0.007 -0.010 |
| indirect6 0.004 0.003 1.068 0.286 0.004 0.007 |
| indirect7 0.005 0.005 0.835 0.404 0.005 0.009 |
| indirect8 -0.000 0.002 -0.080 0.936 -0.000 -0.000 |
| indirect9 0.000 0.002 0.161 0.872 0.000 0.001 |
| indirect10 0.002 0.008 0.193 0.847 0.002 0.003 |
| indirect11 0.010 0.008 1.228 0.219 0.010 0.011 |
| indirect12 0.009 0.009 1.008 0.313 0.009 0.009 |
| indirect13 -0.001 0.004 -0.247 0.805 -0.001 -0.001 |
| indirect14 -0.000 0.004 -0.036 0.971 -0.000 -0.000 |
| indirect15 0.024 0.016 1.510 0.131 0.024 0.025 |
| total1 0.466 0.040 11.763 0.000 0.466 0.709 |
| total2 0.468 0.042 11.280 0.000 0.468 0.713 |
| total3 0.462 0.040 11.591 0.000 0.462 0.703 |
| total4 0.465 0.039 11.804 0.000 0.465 0.708 |
| total5 0.457 0.038 12.181 0.000 0.457 0.696 |
| total6 0.272 0.040 6.787 0.000 0.272 0.510 |
| total7 0.273 0.042 6.480 0.000 0.273 0.512 |
| total8 0.268 0.041 6.576 0.000 0.268 0.503 |
| total9 0.268 0.041 6.607 0.000 0.268 0.504 |
| total10 0.270 0.040 6.804 0.000 0.270 0.506 |
| total11 0.247 0.067 3.666 0.000 0.247 0.260 |
| total12 0.245 0.069 3.574 0.000 0.245 0.259 |
| total13 0.236 0.068 3.452 0.001 0.236 0.249 |
| total14 0.236 0.068 3.477 0.001 0.236 0.249 |
| total15 0.260 0.064 4.099 0.000 0.260 0.275 |

| 6.2.3 Third Timepoint |
| --- |
| Estimator ML |
| Optimization method NLMINB |
| Number of free parameters 108 |
| Used Total |
| Number of observations 488 532 |
| Model Test User Model: |
| Test statistic 498.065 |
| Degrees of freedom 135 |
| P-value (Chi-square) 0.000 |
| Model Test Baseline Model: |
| Test statistic 2937.944 |
| Degrees of freedom 225 |
| P-value 0.000 |
| User Model versus Baseline Model: |
| Comparative Fit Index (CFI) 0.866 |
| Tucker-Lewis Index (TLI) 0.777 |
| Loglikelihood and Information Criteria: |
| Loglikelihood user model (H0) -10386.424 |
| Loglikelihood unrestricted model (H1) -10137.391 |
| Akaike (AIC) 20988.847 |
| Bayesian (BIC) 21441.401 |
| Sample-size adjusted Bayesian (BIC) 21098.613 |
| Root Mean Square Error of Approximation: |
| RMSEA 0.074 |
| 90 Percent confidence interval - lower 0.067 |
| 90 Percent confidence interval - upper 0.081 |
| P-value RMSEA <= 0.05 0.000 |
| Standardized Root Mean Square Residual: |
| SRMR 0.072 |
| Parameter Estimates: |
| Standard errors Bootstrap |
| Number of requested bootstrap draws 1000 |
| Number of successful bootstrap draws 1000 |
| Latent Variables: |
| Estimate Std.Err z-value P(>\|z\|) Std.lv Std.all |
| livCond =~ |
| Nmbr_rms___FU2 1.000 1.706 0.798 |
| Nmbr_ppl___FU2 0.408 0.041 9.999 0.000 0.695 0.527 |
| Living_Are_FU2 0.413 0.061 6.757 0.000 0.704 0.509 |
| Grdn_yrd_FU2_2 0.161 0.022 7.197 0.000 0.275 0.599 |
| healthBeforeCo19 =~ |
| Rglr_tr_PI_FU2 1.000 0.199 0.549 |
| MntHS_BC19_FU2 -4.547 0.644 -7.058 0.000 -0.905 -0.882 |
| PhysH_BC19_FU2 -3.135 0.434 -7.228 0.000 -0.624 -0.630 |
| lonely =~ |
| Lonely_FU2 0.891 0.056 15.872 0.000 0.891 0.698 |
| Ngtv_thght_FU2 0.890 0.046 19.160 0.000 0.890 0.764 |
| ChngSclRl__FU2 0.879 0.061 14.331 0.000 0.879 0.650 |
| Regressions: |
| Estimate Std.Err z-value P(>\|z\|) Std.lv Std.all |
| depression ~ |
| hlthBC19 -1.156 0.193 -5.986 0.000 -0.230 -0.323 |
| Cnt__FU2 -0.186 0.062 -3.020 0.003 -0.186 -0.120 |
| age_FU2 -0.003 0.002 -1.807 0.071 -0.003 -0.062 |
| Gender 0.020 0.052 0.376 0.707 0.020 0.013 |
| Hghst___ 0.012 0.015 0.756 0.449 0.012 0.024 |
| livCond 0.043 0.018 2.343 0.019 0.073 0.102 |
| anxiety ~ |
| hlthBC19 -0.895 0.197 -4.542 0.000 -0.178 -0.322 |
| Cnt__FU2 -0.144 0.061 -2.347 0.019 -0.144 -0.119 |
| age_FU2 -0.005 0.001 -3.539 0.000 -0.005 -0.132 |
| Gender 0.114 0.041 2.767 0.006 0.114 0.094 |
| Hghst___ 0.012 0.015 0.806 0.420 0.012 0.034 |
| livCond 0.031 0.016 1.926 0.054 0.052 0.094 |
| SPQ_total ~ |
| hlthBC19 -1.639 0.300 -5.461 0.000 -0.326 -0.331 |
| Cnt__FU2 -0.089 0.105 -0.851 0.395 -0.089 -0.042 |
| age_FU2 -0.010 0.003 -3.468 0.001 -0.010 -0.146 |
| Gender 0.010 0.087 0.120 0.904 0.010 0.005 |
| Hghst___ -0.057 0.028 -2.056 0.040 -0.057 -0.086 |
| livCond -0.010 0.029 -0.334 0.738 -0.016 -0.017 |
| Mscore.g ~ |
| hlthBC19 -0.017 0.126 -0.138 0.890 -0.003 -0.007 |
| Cnt__FU2 -0.077 0.046 -1.678 0.093 -0.077 -0.073 |
| age_FU2 -0.001 0.001 -0.541 0.588 -0.001 -0.024 |
| Gender -0.143 0.046 -3.080 0.002 -0.143 -0.133 |
| Hghst___ -0.077 0.014 -5.533 0.000 -0.077 -0.234 |
| livCond -0.041 0.015 -2.692 0.007 -0.070 -0.143 |
| drug_score.g ~ |
| hlthBC19 -0.285 0.110 -2.587 0.010 -0.057 -0.154 |
| Cnt__FU2 0.051 0.039 1.305 0.192 0.051 0.063 |
| age_FU2 -0.001 0.001 -1.241 0.215 -0.001 -0.054 |
| Gender 0.009 0.033 0.281 0.779 0.009 0.012 |
| Hghst___ -0.007 0.011 -0.641 0.522 -0.007 -0.028 |
| livCond -0.014 0.011 -1.254 0.210 -0.024 -0.065 |
| Alcohol_FU2 ~ |
| hlthBC19 1.553 0.593 2.617 0.009 0.309 0.138 |
| Cnt__FU2 -0.226 0.214 -1.059 0.290 -0.226 -0.047 |
| age_FU2 0.025 0.007 3.594 0.000 0.025 0.170 |
| Gender -0.189 0.226 -0.836 0.403 -0.189 -0.039 |
| Hghst___ 0.225 0.069 3.271 0.001 0.225 0.151 |
| livCond -0.016 0.065 -0.253 0.800 -0.028 -0.013 |
| exerciseMin_FU2 ~ |
| hlthBC19 -0.137 0.317 -0.432 0.666 -0.027 -0.023 |
| Cnt__FU2 -0.261 0.117 -2.221 0.026 -0.261 -0.104 |
| age_FU2 0.005 0.004 1.252 0.210 0.005 0.061 |
| Gender 0.125 0.111 1.121 0.262 0.125 0.049 |
| Hghst___ 0.155 0.035 4.466 0.000 0.155 0.199 |
| livCond 0.057 0.038 1.478 0.139 0.097 0.083 |
| Sleep_week_FU2 ~ |
| hlthBC19 -0.167 0.199 -0.837 0.403 -0.033 -0.052 |
| Cnt__FU2 0.124 0.071 1.742 0.082 0.124 0.090 |
| age_FU2 -0.004 0.002 -1.922 0.055 -0.004 -0.095 |
| Gender 0.253 0.059 4.292 0.000 0.253 0.182 |
| Hghst___ 0.025 0.018 1.380 0.168 0.025 0.059 |
| livCond -0.022 0.021 -1.087 0.277 -0.038 -0.060 |
| healthBeforeCo19 ~ |
| Cnt__FU2 0.051 0.027 1.930 0.054 0.258 0.119 |
| age_FU2 0.001 0.001 1.978 0.048 0.006 0.097 |
| Gender -0.022 0.021 -1.028 0.304 -0.111 -0.051 |
| Hghst___ 0.002 0.007 0.317 0.751 0.010 0.016 |
| livCond 0.013 0.009 1.457 0.145 0.111 0.111 |
| depression ~ |
| lonely (c1) 0.494 0.036 13.744 0.000 0.494 0.693 |
| anxiety ~ |
| lonely (c2) 0.262 0.034 7.783 0.000 0.262 0.473 |
| SPQ_total ~ |
| lonely (c3) 0.234 0.060 3.886 0.000 0.234 0.237 |
| Mscore.g ~ |
| lonely (a1) 0.108 0.025 4.279 0.000 0.108 0.222 |
| drug_score.g ~ |
| lonely (a2) 0.103 0.022 4.651 0.000 0.103 0.280 |
| Alcohol_FU2 ~ |
| lonely (a3) 0.161 0.128 1.255 0.209 0.161 0.072 |
| exerciseMin_FU2 ~ |
| lonely (a4) -0.033 0.062 -0.542 0.587 -0.033 -0.029 |
| Sleep_week_FU2 ~ |
| lonely (a5) -0.129 0.038 -3.426 0.001 -0.129 -0.202 |
| depression ~ |
| Mscore.g (b1) 0.128 0.054 2.348 0.019 0.128 0.087 |
| drg_scr. (b2) 0.035 0.081 0.435 0.664 0.035 0.018 |
| Alch_FU2 (b3) 0.015 0.012 1.276 0.202 0.015 0.047 |
| exrM_FU2 (b4) -0.037 0.021 -1.757 0.079 -0.037 -0.060 |
| Slp__FU2 (b5) -0.010 0.040 -0.245 0.807 -0.010 -0.009 |
| anxiety ~ |
| Mscore.g (b6) 0.128 0.051 2.522 0.012 0.128 0.113 |
| drg_scr. (b7) 0.080 0.078 1.021 0.307 0.080 0.053 |
| Alch_FU2 (b8) -0.011 0.011 -0.986 0.324 -0.011 -0.045 |
| exrM_FU2 (b9) 0.000 0.017 0.030 0.976 0.000 0.001 |
| Slp__FU2 (b10) 0.022 0.045 0.496 0.620 0.022 0.025 |
| SPQ_total ~ |
| Mscore.g (b11) 0.250 0.086 2.918 0.004 0.250 0.124 |
| drg_scr. (b12) 0.054 0.140 0.387 0.699 0.054 0.020 |
| Alch_FU2 (b13) -0.022 0.021 -1.064 0.288 -0.022 -0.050 |
| exrM_FU2 (b14) -0.003 0.035 -0.083 0.934 -0.003 -0.003 |
| Slp__FU2 (b15) -0.102 0.079 -1.293 0.196 -0.102 -0.066 |
| Covariances: |
| Estimate Std.Err z-value P(>\|z\|) Std.lv Std.all |
| livCond ~~ |
| lonely -0.207 0.107 -1.928 0.054 -0.121 -0.121 |
| .depression ~~ |
| .anxiety 0.073 0.016 4.593 0.000 0.073 0.431 |
| .SPQ_total 0.125 0.027 4.681 0.000 0.125 0.361 |
| .anxiety ~~ |
| .SPQ_total 0.180 0.029 6.323 0.000 0.180 0.526 |
| Variances: |
| Estimate Std.Err z-value P(>\|z\|) Std.lv Std.all |
| lonely 1.000 1.000 1.000 |
| .Nmbr_rms___FU2 1.657 0.355 4.673 0.000 1.657 0.363 |
| .Nmbr_ppl___FU2 1.260 0.160 7.894 0.000 1.260 0.723 |
| .Living_Are_FU2 1.416 0.111 12.777 0.000 1.416 0.741 |
| .Grdn_yrd_FU2_2 0.135 0.012 11.172 0.000 0.135 0.641 |
| .Rglr_tr_PI_FU2 0.092 0.009 9.805 0.000 0.092 0.699 |
| .MntHS_BC19_FU2 0.234 0.075 3.114 0.002 0.234 0.223 |
| .PhysH_BC19_FU2 0.592 0.052 11.425 0.000 0.592 0.603 |
| .Lonely_FU2 0.836 0.071 11.783 0.000 0.836 0.513 |
| .Ngtv_thght_FU2 0.566 0.060 9.358 0.000 0.566 0.417 |
| .ChngSclRl__FU2 1.058 0.096 11.073 0.000 1.058 0.578 |
| .depression 0.172 0.020 8.646 0.000 0.172 0.337 |
| .anxiety 0.169 0.022 7.605 0.000 0.169 0.550 |
| .SPQ_total 0.698 0.058 11.971 0.000 0.698 0.719 |
| .Mscore.g 0.203 0.009 23.234 0.000 0.203 0.853 |
| .drug_score.g 0.119 0.009 12.698 0.000 0.119 0.878 |
| .Alcohol_FU2 4.548 0.170 26.709 0.000 4.548 0.911 |
| .exerciseMn_FU2 1.255 0.078 16.076 0.000 1.255 0.932 |
| .Sleep_week_FU2 0.367 0.027 13.836 0.000 0.367 0.907 |
| livCond 2.910 0.431 6.757 0.000 1.000 1.000 |
| .healthBeforC19 0.038 0.009 4.262 0.000 0.960 0.960 |
| Defined Parameters: |
| Estimate Std.Err z-value P(>\|z\|) Std.lv Std.all |
| indirect1 0.014 0.006 2.260 0.024 0.014 0.019 |
| indirect2 0.004 0.008 0.437 0.662 0.004 0.005 |
| indirect3 0.002 0.003 0.891 0.373 0.002 0.003 |
| indirect4 0.001 0.003 0.494 0.621 0.001 0.002 |
| indirect5 0.001 0.005 0.235 0.814 0.001 0.002 |
| indirect6 0.014 0.006 2.313 0.021 0.014 0.025 |
| indirect7 0.008 0.008 1.022 0.307 0.008 0.015 |
| indirect8 -0.002 0.003 -0.634 0.526 -0.002 -0.003 |
| indirect9 -0.000 0.001 -0.015 0.988 -0.000 -0.000 |
| indirect10 -0.003 0.006 -0.486 0.627 -0.003 -0.005 |
| indirect11 0.027 0.011 2.396 0.017 0.027 0.028 |
| indirect12 0.006 0.014 0.385 0.700 0.006 0.006 |
| indirect13 -0.004 0.005 -0.711 0.477 -0.004 -0.004 |
| indirect14 0.000 0.002 0.041 0.968 0.000 0.000 |
| indirect15 0.013 0.011 1.219 0.223 0.013 0.013 |
| total1 0.508 0.036 14.194 0.000 0.508 0.713 |
| total2 0.498 0.035 14.254 0.000 0.498 0.698 |
| total3 0.497 0.036 13.776 0.000 0.497 0.697 |
| total4 0.496 0.036 13.738 0.000 0.496 0.695 |
| total5 0.496 0.035 14.081 0.000 0.496 0.695 |
| total6 0.276 0.033 8.301 0.000 0.276 0.498 |
| total7 0.270 0.035 7.790 0.000 0.270 0.487 |
| total8 0.260 0.033 7.781 0.000 0.260 0.469 |
| total9 0.262 0.034 7.793 0.000 0.262 0.473 |
| total10 0.259 0.032 8.022 0.000 0.259 0.467 |
| total11 0.261 0.059 4.404 0.000 0.261 0.265 |
| total12 0.239 0.058 4.151 0.000 0.239 0.243 |
| total13 0.230 0.060 3.845 0.000 0.230 0.234 |
| total14 0.234 0.060 3.893 0.000 0.234 0.237 |
| total15 0.247 0.058 4.252 0.000 0.247 0.250 |

| 6.2.4 Fourth Timepoint |
| --- |
| Estimator ML |
| Optimization method NLMINB |
| Number of free parameters 108 |
| Used Total |
| Number of observations 440 478 |
| Model Test User Model: |
| Test statistic 448.081 |
| Degrees of freedom 135 |
| P-value (Chi-square) 0.000 |
| Model Test Baseline Model: |
| Test statistic 2567.853 |
| Degrees of freedom 225 |
| P-value 0.000 |
| User Model versus Baseline Model: |
| Comparative Fit Index (CFI) 0.866 |
| Tucker-Lewis Index (TLI) 0.777 |
| Loglikelihood and Information Criteria: |
| Loglikelihood user model (H0) -9024.252 |
| Loglikelihood unrestricted model (H1) -8800.211 |
| Akaike (AIC) 18264.504 |
| Bayesian (BIC) 18705.875 |
| Sample-size adjusted Bayesian (BIC) 18363.135 |
| Root Mean Square Error of Approximation: |
| RMSEA 0.073 |
| 90 Percent confidence interval - lower 0.065 |
| 90 Percent confidence interval - upper 0.080 |
| P-value RMSEA <= 0.05 0.000 |
| Standardized Root Mean Square Residual: |
| SRMR 0.070 |
| Parameter Estimates: |
| Standard errors Bootstrap |
| Number of requested bootstrap draws 1000 |
| Number of successful bootstrap draws 1000 |
| Latent Variables: |
| Estimate Std.Err z-value P(>\|z\|) Std.lv Std.all |
| livCond =~ |
| Nmbr_rms___FU3 1.000 1.786 0.853 |
| Nmbr_ppl___FU3 0.389 0.035 11.025 0.000 0.694 0.592 |
| Living_Are_FU3 0.345 0.052 6.693 0.000 0.616 0.459 |
| Grdn_yrd_FU3_2 0.164 0.020 8.248 0.000 0.293 0.618 |
| healthBeforeCo19 =~ |
| Rglr_tr_PI_FU3 1.000 0.148 0.442 |
| MntHS_BC19_FU3 -5.515 1.265 -4.359 0.000 -0.819 -0.808 |
| PhysH_BC19_FU3 -4.094 0.923 -4.435 0.000 -0.608 -0.656 |
| lonely =~ |
| Lonely_FU3 0.888 0.059 14.941 0.000 0.888 0.682 |
| Ngtv_thght_FU3 0.943 0.044 21.353 0.000 0.943 0.826 |
| ChngSclRl__FU3 0.744 0.063 11.904 0.000 0.744 0.588 |
| Regressions: |
| Estimate Std.Err z-value P(>\|z\|) Std.lv Std.all |
| depression ~ |
| hlthBC19 -1.419 0.333 -4.267 0.000 -0.211 -0.285 |
| Cnt__FU3 -0.098 0.067 -1.472 0.141 -0.098 -0.055 |
| age_FU3 -0.002 0.002 -1.139 0.255 -0.002 -0.035 |
| Gender -0.103 0.059 -1.760 0.078 -0.103 -0.060 |
| Hgh__FU3 -0.060 0.020 -3.006 0.003 -0.060 -0.113 |
| livCond 0.003 0.019 0.160 0.873 0.005 0.007 |
| anxiety ~ |
| hlthBC19 -1.245 0.263 -4.741 0.000 -0.185 -0.345 |
| Cnt__FU3 -0.179 0.064 -2.798 0.005 -0.179 -0.139 |
| age_FU3 -0.005 0.001 -3.849 0.000 -0.005 -0.148 |
| Gender -0.049 0.054 -0.907 0.364 -0.049 -0.039 |
| Hgh__FU3 -0.062 0.016 -3.790 0.000 -0.062 -0.162 |
| livCond -0.007 0.015 -0.481 0.630 -0.013 -0.024 |
| SPQ_total ~ |
| hlthBC19 -2.342 0.554 -4.225 0.000 -0.348 -0.358 |
| Cnt__FU3 -0.139 0.131 -1.062 0.288 -0.139 -0.060 |
| age_FU3 -0.006 0.003 -2.025 0.043 -0.006 -0.089 |
| Gender -0.261 0.101 -2.590 0.010 -0.261 -0.115 |
| Hgh__FU3 -0.135 0.033 -4.115 0.000 -0.135 -0.194 |
| livCond -0.039 0.026 -1.481 0.139 -0.070 -0.072 |
| media_score_FU3.g ~ |
| hlthBC19 -0.303 0.192 -1.583 0.113 -0.045 -0.097 |
| Cnt__FU3 -0.068 0.052 -1.309 0.191 -0.068 -0.061 |
| age_FU3 -0.000 0.002 -0.135 0.893 -0.000 -0.007 |
| Gender -0.174 0.051 -3.382 0.001 -0.174 -0.160 |
| Hgh__FU3 -0.039 0.016 -2.369 0.018 -0.039 -0.116 |
| livCond -0.024 0.014 -1.639 0.101 -0.042 -0.091 |
| drug_score_during_FU3.g ~ |
| hlthBC19 -0.489 0.157 -3.120 0.002 -0.073 -0.194 |
| Cnt__FU3 -0.008 0.045 -0.179 0.858 -0.008 -0.009 |
| age_FU3 0.002 0.001 1.468 0.142 0.002 0.067 |
| Gender -0.015 0.042 -0.353 0.724 -0.015 -0.017 |
| Hgh__FU3 -0.040 0.013 -3.036 0.002 -0.040 -0.150 |
| livCond -0.024 0.011 -2.207 0.027 -0.042 -0.113 |
| Alcohol_FU3 ~ |
| hlthBC19 2.759 1.138 2.425 0.015 0.410 0.182 |
| Cnt__FU3 0.056 0.266 0.210 0.834 0.056 0.010 |
| age_FU3 0.023 0.008 2.958 0.003 0.023 0.149 |
| Gender -0.458 0.246 -1.864 0.062 -0.458 -0.087 |
| Hgh__FU3 0.218 0.080 2.728 0.006 0.218 0.135 |
| livCond 0.077 0.066 1.168 0.243 0.138 0.061 |
| exerciseMin_FU3 ~ |
| hlthBC19 0.182 0.481 0.379 0.705 0.027 0.024 |
| Cnt__FU3 -0.053 0.145 -0.363 0.716 -0.053 -0.020 |
| age_FU3 0.002 0.004 0.486 0.627 0.002 0.026 |
| Gender -0.050 0.134 -0.374 0.708 -0.050 -0.019 |
| Hgh__FU3 0.140 0.036 3.887 0.000 0.140 0.175 |
| livCond 0.030 0.039 0.766 0.444 0.054 0.049 |
| Sleep_week_FU3 ~ |
| hlthBC19 0.190 0.172 1.107 0.268 0.028 0.070 |
| Cnt__FU3 0.053 0.051 1.037 0.300 0.053 0.055 |
| age_FU3 -0.001 0.001 -0.600 0.549 -0.001 -0.026 |
| Gender 0.066 0.046 1.441 0.150 0.066 0.071 |
| Hgh__FU3 0.038 0.014 2.687 0.007 0.038 0.131 |
| livCond 0.004 0.012 0.357 0.721 0.007 0.018 |
| healthBeforeCo19 ~ |
| Cnt__FU3 0.046 0.022 2.062 0.039 0.308 0.128 |
| age_FU3 0.001 0.001 1.487 0.137 0.006 0.091 |
| Gender 0.008 0.019 0.429 0.668 0.054 0.023 |
| Hgh__FU3 -0.009 0.006 -1.455 0.146 -0.059 -0.082 |
| livCond -0.001 0.006 -0.195 0.846 -0.014 -0.014 |
| depression ~ |
| lonely (c1) 0.552 0.035 15.699 0.000 0.552 0.747 |
| anxiety ~ |
| lonely (c2) 0.287 0.030 9.465 0.000 0.287 0.535 |
| SPQ_total ~ |
| lonely (c3) 0.328 0.062 5.315 0.000 0.328 0.337 |
| media_score_FU3.g ~ |
| lonely (a1) 0.051 0.027 1.915 0.056 0.051 0.111 |
| drug_score_during_FU3.g ~ |
| lonely (a2) 0.058 0.024 2.386 0.017 0.058 0.154 |
| Alcohol_FU3 ~ |
| lonely (a3) 0.053 0.135 0.395 0.693 0.053 0.024 |
| exerciseMin_FU3 ~ |
| lonely (a4) -0.048 0.061 -0.782 0.434 -0.048 -0.043 |
| Sleep_week_FU3 ~ |
| lonely (a5) -0.095 0.024 -4.048 0.000 -0.095 -0.238 |
| depression ~ |
| md__FU3. (b1) 0.058 0.060 0.969 0.333 0.058 0.037 |
| d___FU3. (b2) 0.088 0.083 1.064 0.287 0.088 0.045 |
| Alch_FU3 (b3) 0.019 0.013 1.443 0.149 0.019 0.058 |
| exrM_FU3 (b4) -0.006 0.024 -0.260 0.795 -0.006 -0.009 |
| Slp__FU3 (b5) 0.013 0.075 0.180 0.857 0.013 0.007 |
| anxiety ~ |
| md__FU3. (b6) 0.087 0.046 1.894 0.058 0.087 0.076 |
| d___FU3. (b7) -0.021 0.075 -0.283 0.777 -0.021 -0.015 |
| Alch_FU3 (b8) 0.007 0.010 0.658 0.511 0.007 0.028 |
| exrM_FU3 (b9) 0.049 0.019 2.542 0.011 0.049 0.102 |
| Slp__FU3 (b10) 0.033 0.062 0.529 0.597 0.033 0.025 |
| SPQ_total ~ |
| md__FU3. (b11) -0.001 0.092 -0.016 0.987 -0.001 -0.001 |
| d___FU3. (b12) 0.113 0.135 0.836 0.403 0.113 0.044 |
| Alch_FU3 (b13) -0.017 0.020 -0.869 0.385 -0.017 -0.040 |
| exrM_FU3 (b14) 0.045 0.042 1.083 0.279 0.045 0.052 |
| Slp__FU3 (b15) 0.051 0.140 0.363 0.717 0.051 0.021 |
| Covariances: |
| Estimate Std.Err z-value P(>\|z\|) Std.lv Std.all |
| .anxiety ~~ |
| .SPQ_total 0.132 0.024 5.514 0.000 0.132 0.438 |
| .depression ~~ |
| .SPQ_total 0.073 0.024 2.967 0.003 0.073 0.220 |
| .anxiety 0.048 0.014 3.354 0.001 0.048 0.302 |
| livCond ~~ |
| lonely -0.251 0.119 -2.109 0.035 -0.141 -0.141 |
| Variances: |
| Estimate Std.Err z-value P(>\|z\|) Std.lv Std.all |
| lonely 1.000 1.000 1.000 |
| .Nmbr_rms___FU3 1.196 0.312 3.833 0.000 1.196 0.273 |
| .Nmbr_ppl___FU3 0.894 0.063 14.231 0.000 0.894 0.650 |
| .Living_Are_FU3 1.426 0.102 13.971 0.000 1.426 0.790 |
| .Grdn_yrd_FU3_2 0.139 0.012 11.508 0.000 0.139 0.618 |
| .Rglr_tr_PI_FU3 0.091 0.009 9.569 0.000 0.091 0.805 |
| .MntHS_BC19_FU3 0.357 0.098 3.624 0.000 0.357 0.347 |
| .PhysH_BC19_FU3 0.489 0.060 8.097 0.000 0.489 0.570 |
| .Lonely_FU3 0.908 0.088 10.365 0.000 0.908 0.535 |
| .Ngtv_thght_FU3 0.415 0.054 7.623 0.000 0.415 0.318 |
| .ChngSclRl__FU3 1.047 0.085 12.263 0.000 1.047 0.654 |
| .depression 0.173 0.022 7.971 0.000 0.173 0.317 |
| .anxiety 0.144 0.017 8.434 0.000 0.144 0.503 |
| .SPQ_total 0.632 0.062 10.163 0.000 0.632 0.670 |
| .medi_scr_FU3.g 0.199 0.009 21.337 0.000 0.199 0.927 |
| .drg_scr_d_FU3. 0.126 0.011 11.602 0.000 0.126 0.903 |
| .Alcohol_FU3 4.635 0.202 22.916 0.000 4.635 0.911 |
| .exerciseMn_FU3 1.185 0.084 14.115 0.000 1.185 0.962 |
| .Sleep_week_FU3 0.147 0.010 14.163 0.000 0.147 0.915 |
| livCond 3.189 0.415 7.683 0.000 1.000 1.000 |
| .healthBeforC19 0.021 0.008 2.745 0.006 0.966 0.966 |
| Defined Parameters: |
| Estimate Std.Err z-value P(>\|z\|) Std.lv Std.all |
| indirect1 0.003 0.003 0.855 0.393 0.003 0.004 |
| indirect2 0.005 0.006 0.911 0.362 0.005 0.007 |
| indirect3 0.001 0.003 0.322 0.747 0.001 0.001 |
| indirect4 0.000 0.002 0.156 0.876 0.000 0.000 |
| indirect5 -0.001 0.008 -0.166 0.868 -0.001 -0.002 |
| indirect6 0.004 0.003 1.305 0.192 0.004 0.008 |
| indirect7 -0.001 0.005 -0.260 0.795 -0.001 -0.002 |
| indirect8 0.000 0.002 0.200 0.842 0.000 0.001 |
| indirect9 -0.002 0.003 -0.699 0.485 -0.002 -0.004 |
| indirect10 -0.003 0.006 -0.492 0.623 -0.003 -0.006 |
| indirect11 -0.000 0.006 -0.013 0.989 -0.000 -0.000 |
| indirect12 0.007 0.009 0.746 0.455 0.007 0.007 |
| indirect13 -0.001 0.004 -0.240 0.810 -0.001 -0.001 |
| indirect14 -0.002 0.004 -0.515 0.606 -0.002 -0.002 |
| indirect15 -0.005 0.014 -0.337 0.736 -0.005 -0.005 |
| total1 0.555 0.035 15.789 0.000 0.555 0.751 |
| total2 0.557 0.035 15.756 0.000 0.557 0.754 |
| total3 0.553 0.035 15.671 0.000 0.553 0.748 |
| total4 0.552 0.035 15.732 0.000 0.552 0.747 |
| total5 0.550 0.034 15.997 0.000 0.550 0.745 |
| total6 0.291 0.030 9.581 0.000 0.291 0.544 |
| total7 0.285 0.030 9.452 0.000 0.285 0.533 |
| total8 0.287 0.030 9.515 0.000 0.287 0.536 |
| total9 0.284 0.030 9.353 0.000 0.284 0.531 |
| total10 0.283 0.029 9.849 0.000 0.283 0.530 |
| total11 0.328 0.061 5.335 0.000 0.328 0.337 |
| total12 0.334 0.062 5.422 0.000 0.334 0.344 |
| total13 0.327 0.061 5.334 0.000 0.327 0.336 |
| total14 0.325 0.062 5.277 0.000 0.325 0.335 |
| total15 0.323 0.056 5.801 0.000 0.323 0.332 |

### 7 Model with reduced in complexity, with one predictor and one outcome without control variables

| Table 2. Model fit for model with reduced complexity, one predictor, one outcome, five mediators, example for timepoint 1 | | | | | | |
| --- | --- | --- | --- | --- | --- | --- |
|  | Fit index: | Teststatistic | DF | *X^2^* | CFI | RMSEA |
| Predictor | Outcome |  |  |  |  |  |
| COVID-19 related life concerns | SPQ | 40.31 | 22 | .010 | 0.922 | 0.042 |
|  | Anxiety | 61.28 | 22 | .000 | 0.843 | 0.061 |
|  | Depression | 83.31 | 22 | .000 | 0.817 | 0.076 |
| Social adversity | SPQ | 46.51 | 22 | .002 | 0.938 | 0.048 |
|  | Anxiety | 43.13 | 22 | .005 | 0.955 | 0.045 |
|  | Depression | 46.71 | 22 | .002 | 0.962 | 0.048 |

### 8 Alternative models

#### 8.1 COVID-19 related life concerns’ model – alternative models

The alternative models for the different samples at the four different time points evaluating the effect of ‘COVID-19 related life concerns’ -> schizotypy/anxiety/depression -> alcohol/media/drugs/sleep/exercise revealed worse fit than our first proposed model of of ‘COVID-19 related life concerns’ -> alcohol/media/drugs/sleep/exercise -> schizotyoy/anxiety/depression. See **Table X** for AIC/BIC in comparison.

#### 8.2 ‘Social adversity’ Model – alternative models

The alternative models evaluating the effect of ‘social adversity -> schizotypy/anxiety/depression -> alcohol/media/drugs/sleep/exercise revealed worse fit than our first proposed model of of ‘social adversity’ -> alcohol/media/drugs/sleep/exercise -> schizotypy/anxiety/depression. See **Table x** for AIC/BIC in comparison.

| **Table 3.** Model comparison for original and alternative model. | | | | | | | | |
| --- | --- | --- | --- | --- | --- | --- | --- | --- |
|  | Co19 life concerns | | alternative model | | Social adversity | | alternative model | |
| Timepoint | AIC | BIC | AIC | BIC | AIC | BIC | AIC | BIC |
| 1 | 20691.60 | 21142.37 | 20753.43 | 21233.42 | 20394.58 | 20845.35 | 20443.93 | 20923.92 |
| 2 | 17023.27 | 17462.41 | 17082.42 | 17550.02 | 16811.99 | 17249.11 | 16865.25 | 17330.70 |
| 3 | 21325.87 | 21780.18 | 21464.28 | 21948.03 | 20988.85 | 21441.40 | 21109.03 | 21590.92 |
| 4 | 18711.20 | 19155.48 | 18791.80 | 19264.87 | 18264.50 | 18705.88 | 18326.27 | 18796.25 |
| AIC: Akaike information criterion, BIC: Bayesian information criterion | | | | | | | | |

### 9 **Exploratory model COVID-stress -> Anxiety/Depression -> SPQ**

| **Table 4. Overview of the model fit indices separated by exogeneous latent variable and time point** | | | | | | | | | |
| --- | --- | --- | --- | --- | --- | --- | --- | --- | --- |
|  |  |  |  | exact modelfit |  | relativ modelfit | |  | absolute modelfit |
|  |  | Teststatistic | DF | *X^2^* |  | CFI | TLI |  | RMSEA |
| Predictor | Timepoint |  |  |  |  |  |  |  |  |
| COVID-19 related life concerns | 1 | 396.76 | 90 | .000 |  | 0.838 | 0.765 |  | 0.084 |
|  | 2 | 359.18 | 90 | .000 |  | 0.866 | 0.807 |  | 0.083 |
|  | 3 | 359.08 | 90 | .000 |  | 0.880 | 0.827 |  | 0.077 |
|  | 4 | 332.55 | 90 | .000 |  | 0.871 | 0.814 |  | 0.077 |
| Social adversity | 1 | 416.14 | 90 | .000 |  | 0.853 | 0.787 |  | 0.087 |
|  | 2 | 413.23 | 90 | .000 |  | 0.854 | 0.790 |  | 0.092 |
|  | 3 | 411.72 | 90 | .000 |  | 0.868 | 0.809 |  | 0.085 |
|  | 4 | 347.08 | 90 | .000 |  | 0.880 | 0.826 |  | 0.081 |
| DF: degree of freedom, *X^2^*: Chi squared test, CFI: comparative fit index, TLI: Tucker-Lewis index, RMSEA: root mean square error of approximation | | | | | | | | | |
